# Epigenetic age acceleration is related to cognition and cognitive decline in the Elderly: Results of the Austrian Stroke Prevention Study

**DOI:** 10.1101/2023.11.21.23298753

**Authors:** Piyush Gampawar, Sai Pavan Kumar Veeranki, Katja-Elisabeth Petrovic, Reinhold Schmidt, Helena Schmidt

## Abstract

Epigenetic clocks, an estimate of biological age based on DNA methylation (DNAmAge) are gaining prominence as potential markers of brain ageing. However, consensus is lacking as the repertoire of DNAmAges expands, particularly concerning their ability to predict age-related cognitive changes. In our cohort of 785 elderly, we examined 11 DNAmAges, evaluating their associations with brain ageing in cross-sectional and longitudinal settings. Our results highlighted DNAmAges as strong predictors of cognitive change compared to baseline cognition, albeit varying performance across cognitive domains. DunedinPACE excelled in predicting baseline cognition, while Zhang’s clocks and principal component-based PhenoAge (PCPheno) performed best in predicting cognitive decline. DNAmAges elucidated substantial cognitive variability, matching or surpassing the predictive power of vascular risk factors and ApoE4 genotypes. Notably, in ApoE4 carriers, Zhang’s clock and PCPheno exhibited significantly larger effects, explaining over five times the variability in memory decline compared to non-carriers.

## Background

Brain ageing is a multifaceted process marked by complex physiological, cellular, and molecular changes leading to cognitive decline and age-related neuropathological conditions. The quest to understand these changes and their implications for health outcomes has led to the emergence of biomarkers^1,2^.

Epigenetic clocks are molecular markers based on DNA methylation (DNAm) that estimate an individual’s biological age based on DNAm changes (DNAmAge)^2,3^. The difference between a person’s predicted biological and chronological age is called age acceleration (AA) and indicates the speed of biological ageing relative to chronological ageing. The initial DNAmAges classified as first-generation clocks, namely the Horvath^3^ and the Hannum^4^ clocks were primarily designed to estimate an individual’s chronological age^2,3^. Zhang and colleagues later developed near-perfect age predictors using Elastic Net (Zhang-ENP) and Best Linear Unbiased Prediction (Zhang-BLUP) algorithms, trained on 13,000+ samples^5^. The second-generation DNAmAges, PhenoAge^6^ and GrimAge^7^, incorporated both indicators of physiological state and chronological age to identify DNAm patterns. DunedinPACE (Pace of Aging Calculated from the Epigenome) is a third-generation clock that quantifies changes in the pace of biological ageing over 20 years offering a unique longitudinal perspective^8^. Recent enhancements to Horvath, Hannum, Pheno, and Grim DNAmAges utilised CpG principal components (PC). These refined versions, PCHorvath, PCHannum, PCPheno, and PCGrim, increased the reliability of the existing clocks^9^.

Recently epigenetic clocks were explored in brain ageing, predicting age-related cognitive differences and decline. Despite linking DNAmAges to cognition and neurodegenerative diseases, consensus is lacking across DNAmAges and cognitive domains^10–16^. Studies simultaneously examining cross-sectional and longitudinal relationships within a cohort are scarce, hindering temporal insights^15,17^. Few have investigated the link between DNAmAges and MRI markers like brain volume and white matter hyperintensities (WMH)^16^. Lastly, the potential of enhanced PC-based DNAmAges in brain ageing and the modulatory effects of vascular risk and genetic factors on the DNAmAge-cognition connection remain underexplored.

To address these knowledge gaps, we conducted an exploratory analysis using both cross-sectional and longitudinal data from the Austrian Stroke Prevention Study (ASPS)^18,19^. Our study had several key objectives: 1) assessing the reproducibility of DNAmAges, including the impact of different DNAm processing pipelines. 2) evaluating three generations of DNAmAges, and their PC-based counterparts with brain ageing phenotypes. This encompassed cognitive tests spanning various domains such as executive function (Trail making test-part B (TMT-B) and Wisconsin Card Sorting Test (WCST)), manual dexterity (Purdue Pegboard Test (PPT)), visual and verbal memory, and processing speed as well as dementia screening tools including the mini-mental state examination (MMSE) and the Mattis Dementia Rating Scale (MDRS). Additionally, we also explored the associations with structural brain ageing markers, specifically brain volume and WMH. 3) estimating DNAmAges’ role in explaining unique variability in cognition and cognitive decline, comparing them with established risk factors. 4) testing the modulatory effects of risk factors through subgroup and interaction analyses on the DNAmAge-cognition association.

## Results

### Study design

The present analysis included 796 ASPS participants with baseline DNAm, cognitive tests and brain MRI data. Among them, 485 had a first follow-up after 3 years, and 330 had a second follow-up after 6 years. The mean age was 65.8 years (SD:7.9), with 58% females, 69% hypertensives, and 19% *ApoE4* carriers (Table 1) (Figure 1).

**Figure 1:**
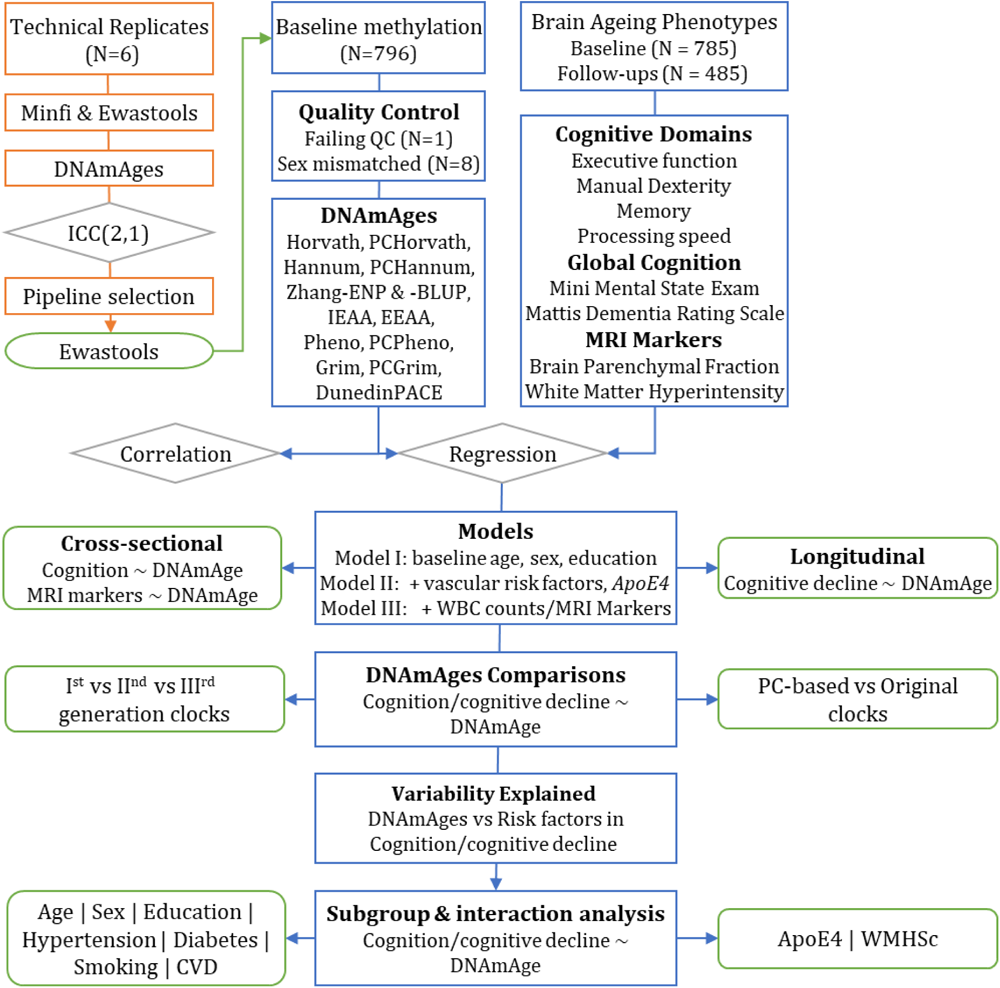
Study design and workflow.

**Table 1:** Study characteristics.

|  | N (%) | Mean (SD) | IQR | Minimum | Maximum |
| --- | --- | --- | --- | --- | --- |
| **Demographics** |  |  |  |  |  |
| Age at baseline (Years) | 785 | 65.8 (7.9) | (60.7, 71.6) | 46.1 | 89.9 |
| Female | 456 (58%) |  |  |  |  |
| **Risk Factors** |  |  |  |  |  |
| Hypertension | 544 (69%) |  |  |  |  |
| Diabetes | 91 (12%) |  |  |  |  |
| CVD | 319 (41%) |  |  |  |  |
| Education |  |  |  |  |  |
| ≤ 9 Years | 223 (28%) |  |  |  |  |
| 10 Years | 318 (41%) |  |  |  |  |
| 13 Years | 173 (22%) |  |  |  |  |
| 18 Years | 71 (9%) |  |  |  |  |
| Smoking |  |  |  |  |  |
| Current | 83 (11%) |  |  |  |  |
| Former | 223 (29%) |  |  |  |  |
| Never | 476 (61%) |  |  |  |  |
| Apoe4 Present | 141 (19%) |  |  |  |  |
| Cholesterol (mg/dl) | 784 | 228.2 (41.6) | (200.8, 253) | 116 | 403 |
| BMI_BL ((kg/m2) | 784 | 26.8 (4) | (24.0, 29.1) | 17.1 | 46.7 |
| **Epigenetic clocks** |  |  |  |  |  |
| Horvath Age | 785 | 63.5 (7.2) | (58.6, 67.8) | 43.8 | 104.7 |
| PCHorvath Age | 785 | 65.2 (6.3) | (61.3, 69.0) | 45.1 | 105.5 |
| Hannum Age | 785 | 57.0 (6.8) | (52.3, 61.4) | 36.7 | 83.2 |
| PCHannum Age | 785 | 69.8 (6.5) | (65.5, 73.8) | 48.5 | 103.7 |
| Zhang-ENP Age | 785 | 66.1 (7.1) | (60.9, 71.2) | 41 | 91.5 |
| Zhang-BLUP Age | 785 | 65.8 (7.1) | (60.9, 70.7) | 41.3 | 89.8 |
| Pheno Age | 785 | 57.1 (8.3) | (51.5, 62.3) | 31.6 | 91.8 |
| PCPheno Age | 785 | 68.9 (6.9) | (64.1, 72.9) | 49.5 | 92.1 |
| Grim Age | 785 | 66.3 (7.5) | (61.3, 71.1) | 39.4 | 111.6 |
| PCGrim Age | 785 | 75.8 (6.6) | (71.2, 80.3) | 59 | 99.8 |
| DunedinPACE | 785 | 1.0 (0.1) | (1.0, 1.1) | 0.7 | 1.5 |
| **Epigenetic Age Acceleration** |  |  |  |  |  |
| Horvath AA | 785 | 0.0 (5.5) | (-3.7, 3.0) | -19.2 | 37.5 |
| PCHorvath AA | 785 | 0.0 (4.9) | (-3.0, 2.5) | -19.7 | 36.8 |
| Hannum AA | 785 | 0.0 (4.7) | (-3.2, 2.5) | -19.9 | 21.8 |
| PCHannum AA | 785 | 0.0 (4.9) | (-3.2, 2.5) | -20.9 | 28.2 |
| IEAA | 785 | 0.1 (5.2) | (-3.3, 2.9) | -20.4 | 34.3 |
| EEAA | 785 | 0.3 (5.8) | (-3.4, 3.6) | -22.6 | 24.2 |
| Zhang-ENP AA | 785 | 0.0 (4.1) | (-2.8, 2.4) | -24.5 | 18.2 |
| Zhang-BLUP AA | 785 | 0.0 (3.9) | (-2.7, 2.3) | -23.9 | 16.5 |
| Pheno AA | 785 | 0.0 (6.5) | (-4.3, 3.9) | -25.1 | 34.8 |
| Grim AA | 785 | 0.0 (4.1) | (-2.8, 2.0) | -9.5 | 20.6 |
| PCPheno AA | 785 | 0.0 (5.3) | (-3.2, 3.0) | -26.4 | 43 |
| PCGrim AA | 785 | 0.0 (3.0) | (-2.2, 1.6) | -6.6 | 14.9 |
| DunedinPACE AA | 785 | 0.0 (0.1) | (-0.08, 0.07) | -0.37 | 0.48 |

We explored the cross-sectional and longitudinal relationship between brain ageing and 11 DNAmAges including first-generation (Horvath, Hannum, Zhang-ENP, Zhang-BLUP), second-generation (Pheno and Grim), and third-generation (DunedinPACE) clocks, along with their PC-based versions (PCHorvath, PCHannum, PCPheno, and PCGrim). We employed AA as a measure of biological ageing in the analyses where positive AA indicates accelerated biological ageing relative to chronological age, while negative AA implies slower biological ageing.

### Effect of pre-processing on DNAmAges

We compared minfi^20^ and ewastools^21^ pipelines by evaluating the reliability of DNAmAges using intra-class correlation (ICC) within six samples. Except for Hannum (0.64), all clocks had excellent reliability (ICC>0.75, Range:0.79 to 1). PC-based clocks notably exhibited the highest reliability (Range:0.90 to 1), surpassing their original versions. Overall, ewastools demonstrated slightly superior ICC values for most clocks compared to minfi (Supplementary Table 1).

### Epigenetic age measures

Quality control excluded nine samples: eight for sex mismatches and one for failing quality control thresholds. The final dataset included 785 participants (Figure 1). DNAmAge calculated using first-generation clocks ranged from 57.1 to 66.1 years, and 57.0 to 69,9 years using second-generation clocks. PC-based clocks had higher mean ages than their original counterparts, with Hannum showing the largest difference of 12.8 years (Table 1, Supplementary Table 2). DunedinPACE had a mean of 1±0.1 years (Range:0.7 to 1.5).

### Correlation Analysis

First-and second-generation clocks showed age correlations from 0.62 to 0.83, while DunedinPACE had a correlation of 0.18. PCHorvath and PCHannum had slightly lower correlations, whereas PCPheno and PCGrim exhibited higher correlations than their original counterparts (Figure 2; Supplementary Table 2). First-and second-generation clocks generally had strong to very strong correlations (0.74-0.94) among themselves. GrimAge, however, displayed moderate to strong correlations (0.62-0.74) with other clocks. DunedinPACE showed low correlations (0.25-0.50), highest with GrimAge (0.50). AA measures had strong to very strong correlations (0.52-0.97), except for Grim, PCGrim, and DunedinPACE AA (0.18-0.52) (Figure 2, Supplementary Table 2).

**Figure 2:**
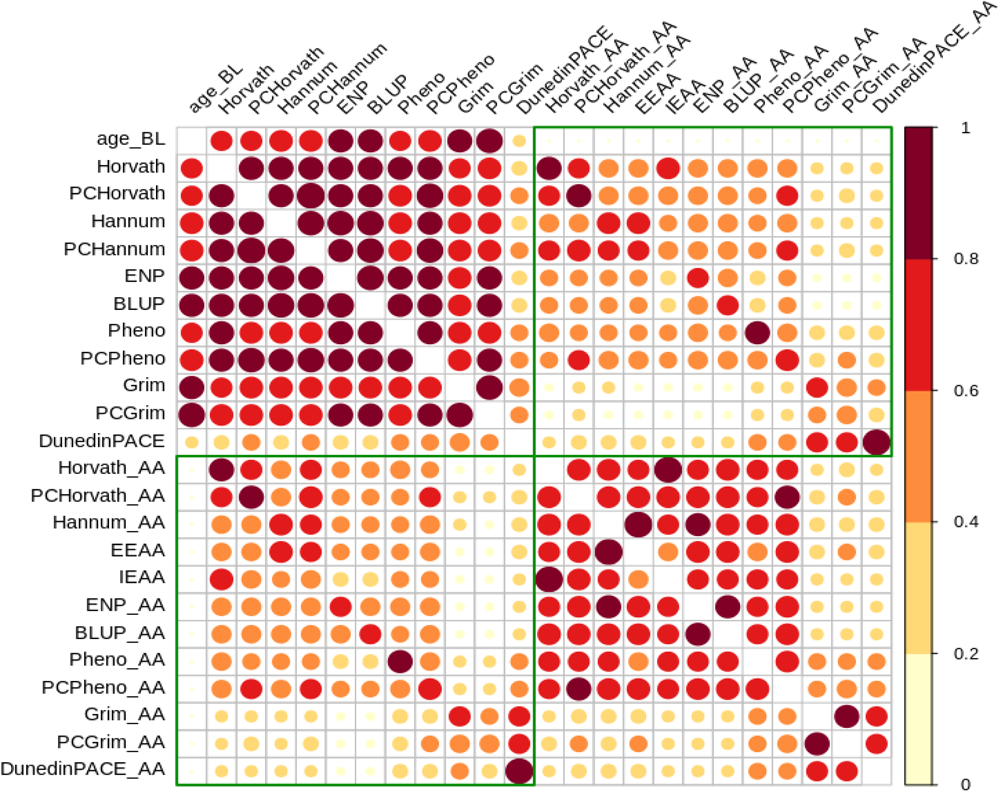
Correlations between DNAmAges and age acceleration measures. AA: Age Acceleration.

### Age acceleration and brain ageing

We hypothesised that higher AA is associated with impaired cognition, reduced BPF, and increased WMH. We employed two nested multiple linear regression models. The first included baseline age, sex, and education years as covariates, while the second included additionally hypertension, BMI, diabetes, cardiovascular diseases (CVD), smoking, total cholesterol, and *ApoE4*-carrier status. Effect size (beta) and partial R^2^ values assessed the extent of the associations.

### Age acceleration and cognition at baseline

Within executive functions, lower WCST performance was significantly associated with higher AA in PCHorvath, Hannum, PCHannum, EEAA, Zhang-ENP, Zhang-BLUP, Pheno, PCPheno, Grim, and DunedinPACE in both models (Model 1: beta =-5.97 to -0.11, partial R^2^ = 0.005 to 0.025; Model 2: beta=-6.56 to -0.12, partial R^2^=0.006 to 0.024). DunedinPACE exhibited the largest effect sizes (≈-6.0), while PCPheno had the highest partial R^2^ in both models (≈0.03). Overall, Model 2 showed higher effect sizes and explained more variability than Model 1. All associations (N=14) except for Grim and DunedinPACE remained significant after false discovery rate (FDR) correction. DunedinPACE was nominally associated with lower TMT-B performance in Model 1 (beta= -40.54; partial R^2^ = 0.01) (Table 2).

**Table 2:**
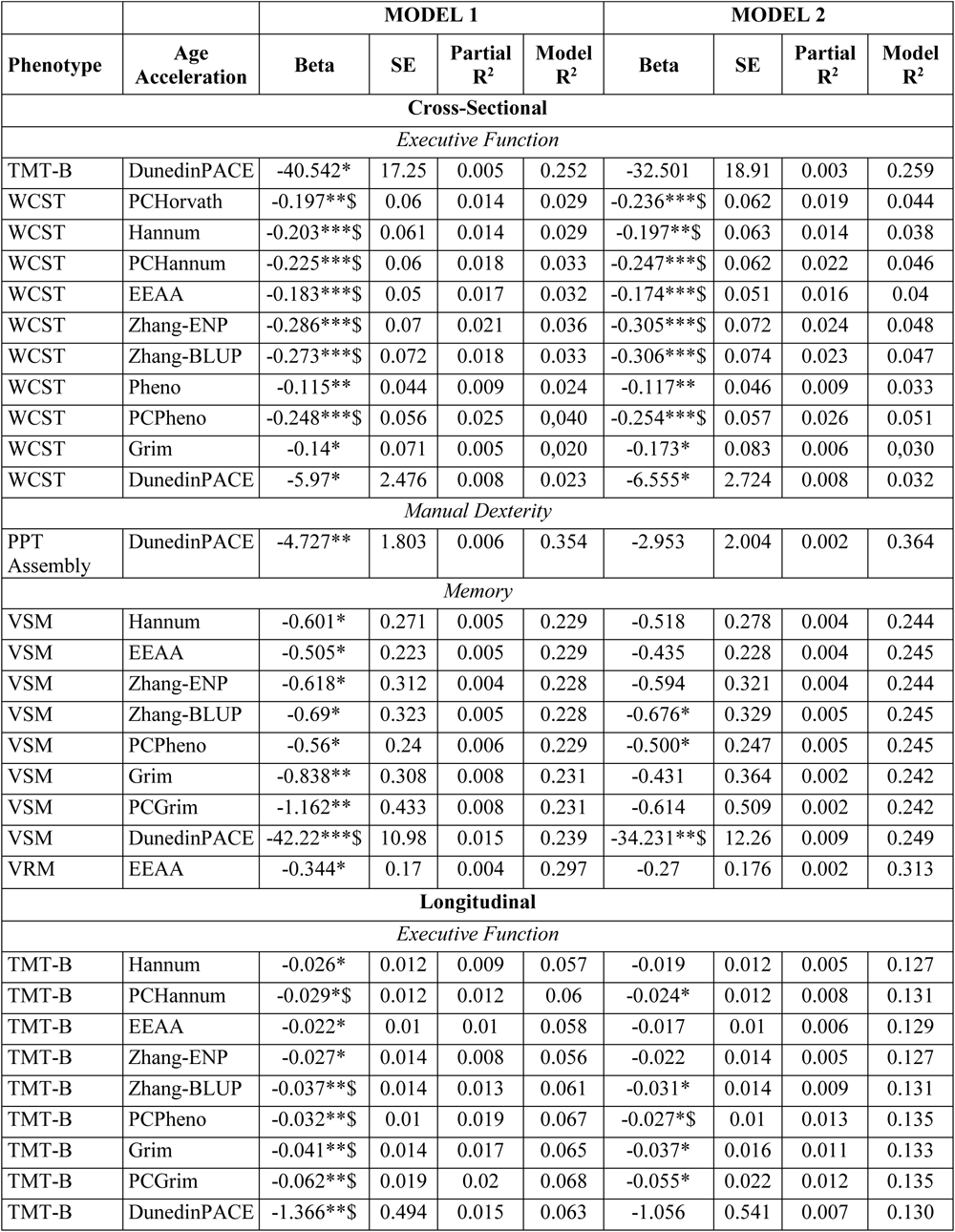
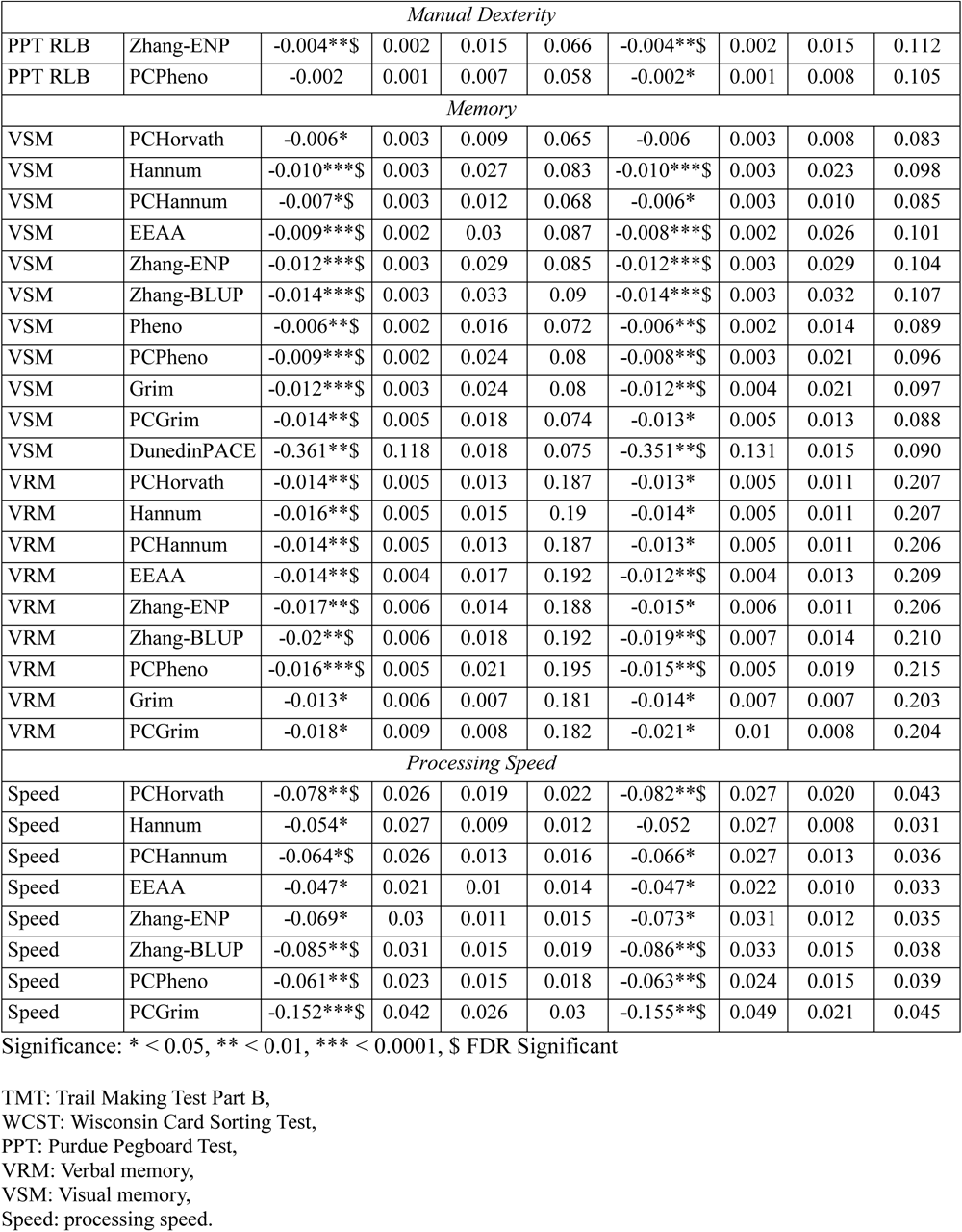
Significant associations.

In the assembly component of PPT evaluating manual dexterity, only DunedinPACE showed a nominal association in Model 1 (beta=-4.73, partial R^2^=0.01) (Table 2).

In the memory domain, lower visual memory scores were nominally associated with higher AA in Hannum, EEAA, Zhang-ENP, Zhang-BLUP, PCPheno, Grim, PCGrim and DunedinPACE (beta=-42.22 to -0.50) in Model 1, and with Zhang-BLUP, PCPheno and DunedinPACE AA in Model 2 (beta=-34.23 to -0.50). DunedinPACE had the largest effect sizes (beta=Model 1:-42.22; Model 2:-34.23), explained the highest variance (partial R^2^= Model 1:0.02, Model 2:0.01), and passed FDR correction. Verbal memory was only nominally associated with EEAA in Model 1 (beta=-0.34, partial R^2^= 0.004) (Table 2).

We also observed an unexpected FDR significant association of higher AA in certain clocks with better cognition including MMSE (Hannum, Zhang-ENP, Zhang-BLUP), PPT-RLB (Zhang-ENP) and processing speed (Zhang-ENP, Zhang-BLUP, and PCPheno) (Supplementary Table 3).

### Age acceleration and cognitive decline

We employed linear mixed models with individual-specific random effects, adjusting for baseline age and follow-up time, to estimate annual cognitive changes. We then assessed the association with DNAmAges using the two models as described in the cross-sectional analyses (Table 2).

In executive function, faster TMT-B decline in Model 1 was nominally associated with Hannum, PCHannum, EEAA, Zhang-ENP, Zhang-BLUP, PCPheno, Grim, PCGrim, and DunedinPACE (beta=-1.37 to -0.02). In Model 2, the association persisted with PCHannum, Zhang-BLUP, PCPheno, Grim, and PCGrim (beta=-0.05 to -0.02). FDR significance was observed for PCHannum, BLUP, PCPheno, Grim, PCGrim, and DunedinPACE in Model 1, as well as PCPheno in Model 2. AA explained 0.8% to 2.0% of the variance in TMT-B decline, with PCPheno explaining the highest in Model 2 (1.56%) (Table 2).

In manual dexterity, faster decline in PPT-RLB was nominally associated with higher Zhang-ENP AA in both models (beta=-0.004, partial R^2^=0.02) as well as PCPheno AA in model 2 (beta=-0.002, partial R^2^= 0.01). Only the association of Zhang-ENP remained significant after FDR correction in both models.

In the memory domain, faster visual memory decline was nominally associated with higher AA in all DNAmAges, except Horvath and IEEA, with many associations passing FDR correction. DunedinPACE exhibited the largest effect size (beta≈-0.36), surpassing other clocks (beta=-0.01) in both models. AA explained variability from 0.9% to 3.3%, with Zhang-BLUP showing the highest values in both models. Similarly, verbal memory decline showed a nominally significant association with PCHorvath, Hannum, PCHannum, EEAA, Zhang-ENP, Zhang-BLUP, PCPheno, Grim and PCGrim (beta=-0.02 to -0.01, partial R^2^=0.01 to 0.02) in both models.

Processing speed decline was associated with higher AA in PCHorvath, Hannum, PCHannum, EEAA, Zhang-ENP, Zhang-BLUP, PCPheno, and PCGrim in model 1 (beta=-0.15 to -0.05, partial R^2^=0.01 to 0.03) and with PCHorvath, PCHannum, EEAA, Zhang-ENP, Zhang-BLUP, PCPheno, and PCGrim in model 2 (beta=-0.15 to -0.05, partial R^2^=0.01 to 0.02). PCGrim AA had the highest effect size (beta =-0.15) and explained the most variability (2.1%) among all significant DNAmAges (Table 2).

Unlike cross-sectional results, most clocks unexpectedly showed FDR significant associations with annual WCST improvement, suggesting possible practice effects. To address this bias, we adjusted WCST scores by subtracting mean differences between the baseline and the first follow-up, as well as between the first and second follow-ups for those with improvements. Despite this correction, AA remained associated with WCST improvement (Supplementary Table 4).

### Age acceleration and MRI Markers

We found no significant associations between AA and decreases in BPF or increases in WMH. However, Hannum, EEAA, Zhang-ENP and Zhang-BLUP AA exhibited an unexpected FDR significant association with higher BPF (Supplementary Table 5).

### Age acceleration, cognition and MRI markers

We explored the impact of MRI markers on AA-cognition associations by adjusting for WMH in the model for executive functions and for BPF for memory.

Cross-sectionally, after WMH adjustment, DNAmAges had no association with TMT-B, but WCST associations remained unchanged. Longitudinally, WMH adjustment slightly reduced second-generation clocks’ effect sizes on TMT-B change (Supplementary Table 6).

Adjusting for BPF in memory analysis retained DunedinPACE’s nominal significance in visual memory with a minimal effect size reduction. Longitudinally, BPF adjustment slightly reduced clock effect sizes for both visual and verbal memory. (Supplementary Table 6).

### Effect of white blood cell counts

We further incorporated estimates of CD8T, CD4T, Natural killer cells, monocytes, neutrophils, and B cells derived from DNA methylation data in the model. These adjustments minimally affected the effect sizes of significant associations, both in the cross-sectional and longitudinal analyses (Supplementary Table 7).

### Comparing DNAmAges

In the subsequent analyses, we focused on phenotypes where DNAmAges showed FDR significant associations in the expected direction. These were WCST and visual memory cross-sectionally and TMT-B, visual memory, verbal memory, and processing speed longitudinally.

### PC-based clocks and cognition

PC-based clocks generally had higher effect sizes and explained more variability in baseline cognition as well as its decline except for the Hannum AA in the memory domain. PC-training had the most significant impact on PCPheno, doubling effect sizes and explained variability compared to Pheno AA in both models (Figure 3, Table 2, Supplementary Table 3 & 4).

**Figure 3:**
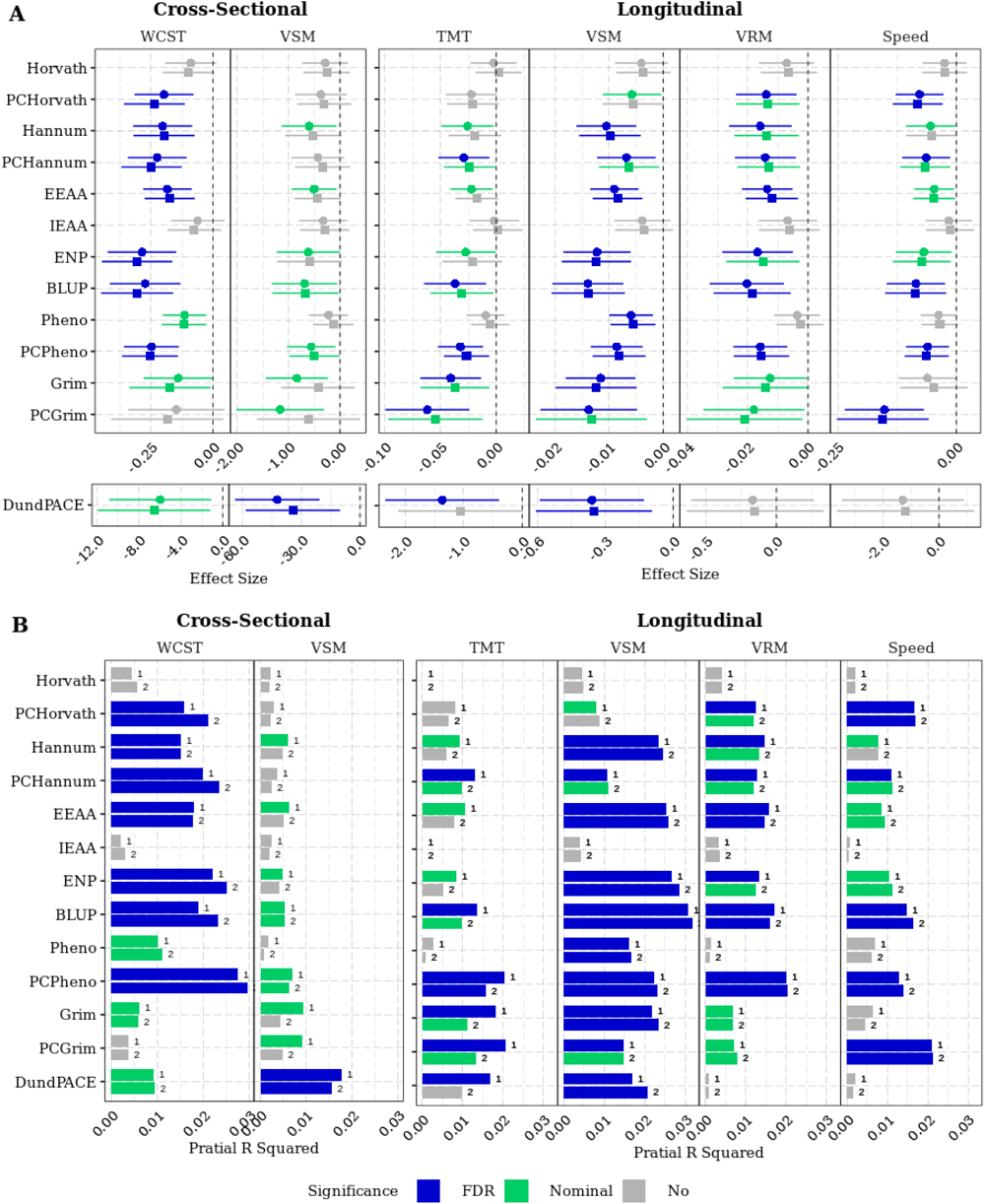
Association of DNAmAges with cognitive phenotypes. **A.** Effect size of cross-sectional and longitudinal associations between DNAmAges and cognitive phenotypes. **B**. Cognitive variability explained by various DNAmAge measures. WCST: Wisconsin Card Sorting Test, TMT: Trail Making Test Part B, VSM: Visual memory, VRM: Verbal memory, speed: Processing speed, DundPACE: DunedinPACE. Circle represent results from model 1 which was adjusted for age, sex and Education, Square represent the result from model 2 which was additionally adjusted for hypertension, BMI, diabetes, cardiovascular diseases, smoking, total cholesterol, and ApoE4-carrier status. Blue indicates FDR significant associations (FDR p<0.05, Green indicates nominally significant associations (p<0.05), Grey indicates non-significant associations

### First, second and third-generation clocks and cognition

DunedinPACE demonstrated the highest effect sizes for baseline WCST and baseline visual memory in both models. However, PCPheno accounted for the highest variability in WCST, while DunedinPACE for visual memory. Longitudinally, PCGrim AA consistently exhibited the highest effect sizes for TMT-B, verbal memory and processing speed decline. DunedinPACE displayed the highest effect size for visual memory decline. Nevertheless, PCPheno AA explained the highest variability in TMT-B and verbal memory decline, Zhang-BLUP AA in visual memory decline, and PCGrim AA in cognitive speed decline (Figure 2, Table 2, Supplementary Table 3 & 4).

### Do all clocks explain the same variability?

Given the weak correlations between DunedinPACE and Zhang-BLUP AA (r=0.18) and their significant associations with baseline visual memory and its decline, we further explored their combined predictive power for visual memory. We included both in a regression model adjusted for demographics, vascular and genetic risk factors. Cross-sectionally, visual memory was significantly associated only with DunedinPACE (beta=-31.0) but not Zhang-BLUP. However, visual memory decline was significantly associated with both Zhang-BLUP (beta=-0.01, partial R^2^=0.026), and DunedinPACE (beta=-0.27, partial R^2^=0.008) (Supplementary Table 8).

### Subgroup analysis

We stratified our cohort by age, sex, education, hypertension, BMI, CVD, smoking, diabetes, *ApoE4* carrier status, and WMH scores. We chose Zhang-BLUP, PCPheno, Grim, and DunedinPACE AA due to their outstanding performance within their respective clock generations. We found stronger Zhang-BLUP and PCPheno associations with cognition, especially in memory decline, in subgroups of young individuals, females, normotensives, lower education, non-smokers, and non-diabetic individuals. Notably, there was a significant interaction between PCPheno and hypertension in visual (beta=0.01, p=0.04) and verbal memory decline (beta=0.02, p=0.04) (Supplementary Table 9).

The *ApoE4*-carrier status showed inconsistent patterns in the AA-cognition relationship (Figure 4; Supplementary Table 10). In memory, Zhang-BLUP and PCPheno had larger effects and explained more variability in carriers that reached significance longitudinally, while DunedinPACE displayed larger effect sizes and partial R^2^ in *ApoE4* non-carriers, reaching significance in visual memory and its decline. In WCST, Zhang-BLUP and PCPheno were significantly associated in both carriers and non-carriers while DunedinPACE only in non-carriers. In processing speed decline, Zhang-BLUP was associated in *ApoE4* carriers while PCPheno and PCGrim in non-carriers. There were significant interactions with *ApoE4* carrier status for Zhang-BLUP on verbal memory decline (beta=-0.04) and processing speed decline (beta=-0.18), and for DunedinPACE on processing speed decline (beta=6.92).

**Figure 4:**
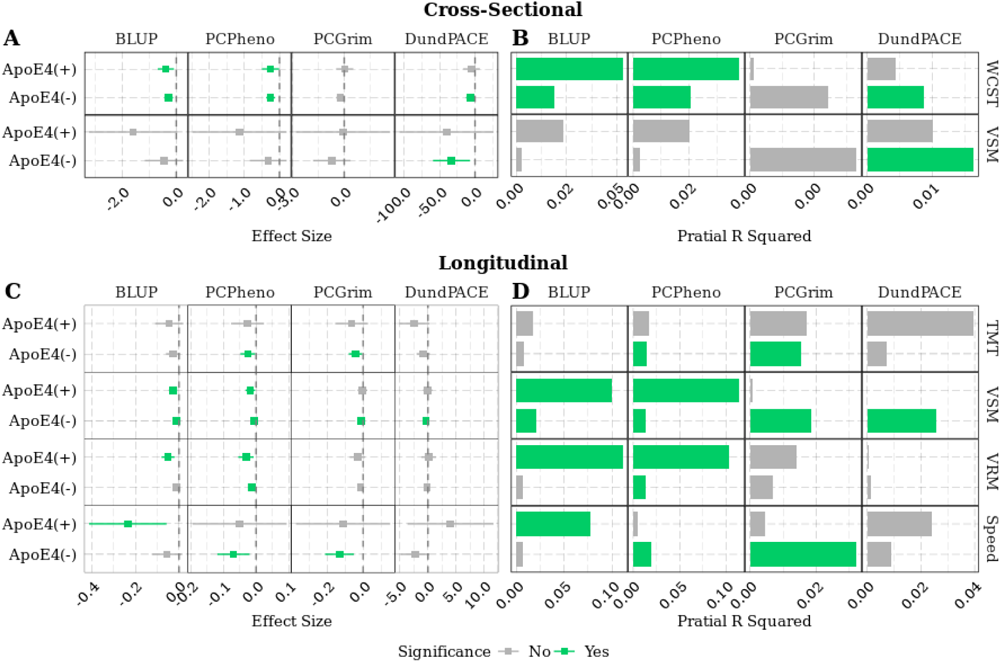
ApoE4-specific subgroup analysis between DNAmAges and cognitive phenotypes. **A**: Cross-sectional effect sizes **B**: Cross-sectional explained cognitive variability. **C:** Longitudinal effect sizes. **D:** Longitudinal explained cognitive variability. WCST: Wisconsin Card Sorting Test, TMT: Trail Making Test Part B, VSM: Visual memory, VRM: Verbal memory, speed: Processing speed, DundPACE: DunedinPACE, Green indicates nominally significant associations (p<0.05), Grey indicates non-significant associations

We did not find any evidence of the specific influence of the WMH on the AA-cognition associations.

## Discussion

Here we present the comprehensive exploration of the association between epigenetic clocks and brain ageing-related cognitive and structural phenotypes including both cross-sectional and longitudinal analyses. Our results support the importance of DNAmAges in explaining variability in cognitive performance in the elderly and especially highlight their potential for predicting longitudinal cognitive changes. Cross-sectionally, DNAmAges were significantly associated with executive function (WCST) and visual memory while longitudinally, they were linked to executive function (TMT-B), visual memory, verbal memory, and processing speed. Importantly, no single DNAmAge performed as a universal predictor of cognition or cognitive decline.

Cross-sectionally the highest effect sizes were observed using Zhang-BLUP in WCST and DunedinPACE in visual memory, and for cognitive decline using PCGrim in executive function, DunedinPACE in visual memory, Zhang-BLUP in verbal memory, and PCGrim in processing speed. DNAmAges contributed 0.5% to 3.0% to explained variability in cognition cross-sectionally, and 0.8% to 3.2% longitudinally. The difference between the first, second, or third-generation clocks was negligible. Yet, DunedinPACE performed superior in cross-sectional and PCPheno in longitudinal models. Adjusting for vascular risk factors and for *ApoE4* status did not significantly change the overall variability explained. Although stronger associations were observed in specific subgroups, such as age, sex, hypertensives and *ApoE4*-carriers, formal tests for interaction were significant only in hypertensives and *ApoE4 carriers* for PCPheno and Zhang-BLUP in memory decline. There was neither an association between AA and MRI markers nor did MRI markers modulate the effect of AA on cognition.

Prior studies have not yielded consistent evidence on the predictive capabilities of DNAmAges for cognition and MRI markers^22–29^. Here, we provide empirical evidence that DNAmAges are better predictors of cognitive decline compared to baseline cognition. Significant associations of DNAmAges with memory and its subdomains were reported mostly cross-sectionally^8,23,25,28–31^ with one study describing also longitudinal associations^17^. In our study, 10 out of 13 DNAmAges had FDR significant association with visual memory decline and seven with verbal memory decline at least in one model. At baseline, only DunedinPACE had an FDR significant association with visual memory and none with verbal memory (Figure 5).

**Figure 5:**
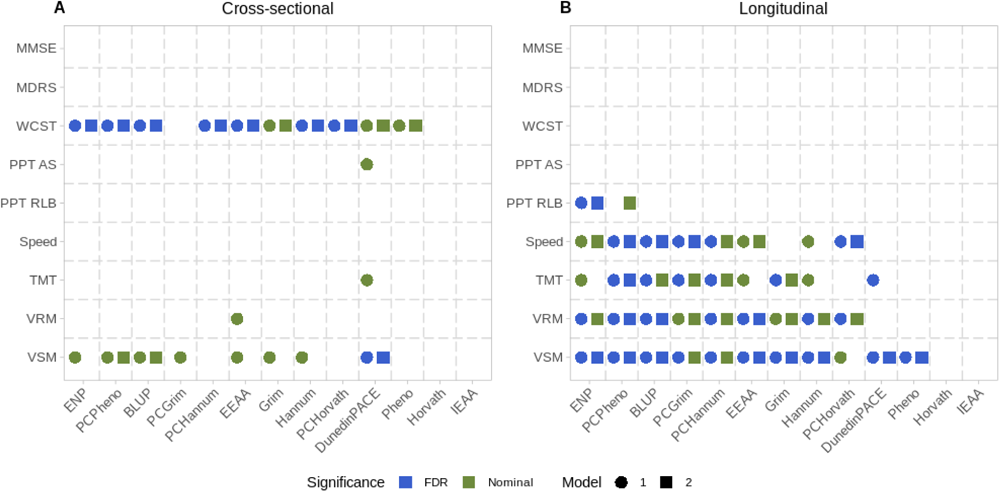
Overview of significant associations between DNAmAges and cognitive phenotypes. **A**: Cross-sectional results. B: Longitudinal Results. Circle represent results from model 1 which was adjusted for age, sex and Education, Square represent the result from model 2 which was additionally adjusted for hypertension, BMI, diabetes, cardiovascular diseases, smoking, total cholesterol, and ApoE4-carrier status. Blue indicates FDR significant associations (FDR p<0.05), Green indicates nominally significant associations (p<0.05), and Grey indicates non-significant associations. MMSE: Mini Mental State Examination, MDRS: Mattis Dementia Rating Scale, WCST: Wisconsin Card Sorting Test: PPT: Purdue Pegboard Test: AS: Assembly, RLB: Right Left Both, Speed: processing speed, TMT: Trail making test part B, VRM: Verbal memory, VSM: Visual memory

**Figure 6:**
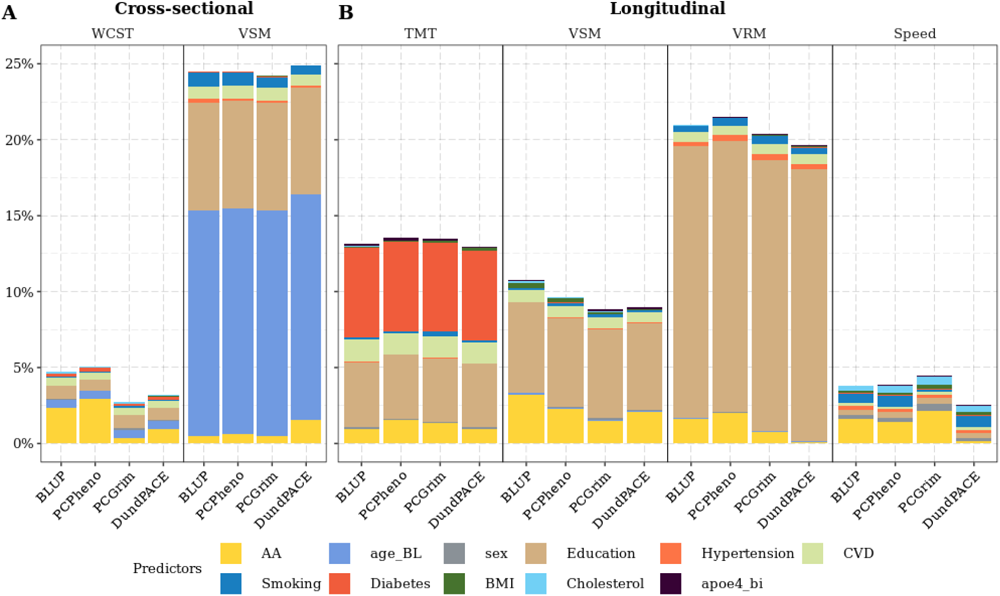
Cognitive variability explained by the complete regression model. **A:** Cross-sectional analysis. **B**: Longitudinal analysis. The X-axis represents DNAmAge used in that model. Various colours represent different covariates used in the model. WCST: Wisconsin Card Sorting Test, VRM: Verbal memory, VSM: Visual memory, Speed: processing speed.

Executive function was the second-most frequently predicted domain by DNAmAges. Its decline, assessed by TMT-B, had FDR significant associations with six DNAmAges in at least one model, but not in the expected direction when measured by WCST (Figure 5). Conversely, at baseline, seven DNAmAges were linked to executive function measured by WCST, but not TMT-B. This pattern of significant associations with TMT-B decline but not with baseline performance aligns with findings from a twin study^32^. Similar non-significant cross-sectional associations between DNAmAges and TMT-B were reported by large studies^23,25,32^. The association between higher DNAmAge and improved WCST scores longitudinally might have been influenced by practice effects^33^. Yet, after accounting for these effects, a significant relationship still persisted.

In the domain of processing speed, FDR significant associations of five DNAmAges existed exclusively with its decline (Figure 5). This contrasts with prior research that linked higher DNAmAges to lower baseline processing speed^8,28–30,34^ but not to its decline^17,28^. Cross-sectionally, higher Zhang-ENP, Zhang-BLUP, and PCPheno AA were associated with improved processing speed. Recently, higher Pheno AA was also linked to reduced variability in processing speed^30^.

Finally, in manual dexterity, a higher Zhang-ENP AA was significantly associated with a decline in PPT RLB while cross-sectionally with an unexpectedly better performance even after FDR adjustment.

Baseline BPF and WMH did not affect the association between DNAmAges and memory or executive functions suggesting that the relation between DNAmAges and cognition is not influenced by structural age-related brain changes.

Visual memory was the only phenotype with significant associations both cross-sectionally and longitudinally with remarkably larger variability explained by the clocks in the longitudinal setting (Zhang-BLUP, PCPheno, and DunedinPACE 0.52%, 0.62%, and 1.56% cross-sectionally, 3.2%, 2.31%, and 2.08% longitudinally) (Figure 5; Supplementary Table 11, Table 2). Compared to established ageing risk factors, DNAmAges consistently ranked among the top three contributors explaining variability in all cognitive domains. They ranked first in baseline executive function and processing speed decline, second after education in memory decline, and third in executive function decline behind education and diabetes. These findings underscore the importance of DNAmAges in predicting cognitive decline, even when compared to well-established ageing risk factors (Figure 5; Supplementary Table 11).

Importantly, we found a significant and independent contribution of Zhang-BLUP and DunedinPACE to visual memory decline when including both clocks in the full model indicating that Zhang-BLUP and DunedinPACE probably capture distinct aspects of visual memory decline related to ageing (Supplementary Table 8).

When assessing the utility of PC-based DNAmAges, we noticed a more substantial impact of PC-training on the association with cognitive decline than on baseline cognition. PCPheno, in particular, displayed a significant improvement, with FDR significant associations in verbal and visual memory, executive function (TMT-B), and processing speed, while Pheno only in visual memory decline. This mirrors findings in the Health and Retirement Study^24^. Additionally, PCPheno’s effect size doubled compared to Pheno AA and explained more variability in cognitive decline than any other clock (Supplementary Table 4 & 5).

Contrary to previous studies^24,25^, we found that first-generation clocks, specifically Zhang-ENP and Zhang-BLUP, demonstrated comparable or even superior predictive performance compared to second-and third-generation clocks. Longitudinally, Zhang-ENP and PCPheno displayed the most associations (N=9), followed by Zhang-BLUP, PCGrim, and PCHannum (N=8) whereas cross-sectionally, DunedinPACE ranked first (N=6), followed by PCPheno, Zhang-BLUP, and EEAA (N=4) (Figure 5). The at-par performance of Zhang-ENP and Zhang-BLUP may be attributed to the extensive sample size used to train these measures^35^.

Although having the highest effect sizes, DunedinPACE didn’t consistently predict cognitive decline and didn’t explain more variability compared to other clocks. This might be attributed to the measurement scale ^24^, where one-unit change in DunedinPACE equals 10 standard deviations (SD), and in other DNAmAges 1/3rd to 1/6th of an SD (Table 1).

Higher Hannum, Zhang-ENP, and Zhang-BLUP AA were associated with FDR significance with better MMSE and MDRS scores cross-sectionally and longitudinally in our study. Previous reports were also inconsistent^17,24–26,29^. These paradoxical findings might be at least partly explained by the fact that these tests were not designed to assess cognitive function in the normal elderly, and a baseline MMSE<24 was an exclusion criterion in ASPS.

In the subgroup analysis, Zhang-BLUP and PCPheno behaved similarly, especially in memory decline, showing higher effect sizes in young, female, low education, normotensive, and non-smoking subgroups, pinpointing to a possible interaction with these factors. The most pronounced difference was observed by hypertension status, where a formally significant interaction between PCPheno and hypertension was also present. Among normotensives, PCPheno explained 11 times more variability in visual memory decline and eight times more in verbal memory decline (Supplementary Table 9). This suggests that DNAmAges’ effect may be masked in hypertensives and possibly in other high-risk groups like the elderly and smokers, where exposure begins in adulthood.

In *ApoE4*-carriers, the full model explained double the cognitive variability compared to non-carriers, except for baseline visual memory (Supplementary Table 9).

Longitudinally, larger effect sizes in *ApoE4-*carriers were even more pronounced with Zhang-BLUP, explaining in visual memory five times and in verbal memory 15 times more variability than in non-carriers. PCPheno explained eight times more variability in *ApoE4*-carriers for both visual and verbal memory. Zhang-BLUP consistently demonstrated higher effect sizes and explained more phenotypic variability in *ApoE4*-carriers across all cognitive domains. Importantly, the interaction was statistically significant when formally tested between *ApoE4* and Zhang-BLUP for verbal memory and processing speed decline as well as for DunedinPACE for processing speed decline. These findings underscore *ApoE4*-specific effects attributed to Zhang-BLUP and PCPheno in executive functions and memory domains, and to Zhang-BLUP in the processing speed domain. *ApoE* is known to play an essential role in human ageing with *ApoE4* recorded as the pro-ageing and *ApoE2* as the longevity allele^36^. The observed interaction indicates that the *ApoE2* allele is able to protect the brain even when biological ageing at the epigenetic level is speeded up. Further studies are needed to better understand this both biologically as well as prognostically relevant interactions with hypertension as well as *ApoE4* status.

Our study is a comprehensive and meticulous approach with notable strengths. Firstly, we ensured methodological robustness by assessing DNAmAge reproducibility and examining the effect of data pre-processing using technical replicates. Secondly, we extensively explored both cross-sectional and longitudinal associations between cognitive measures, MRI markers, and 13 AA measures from three epigenetic clock generations in a well-described population-based elderly cohort, using efficient linear mixed models to estimate individualised cognitive change trajectories. Thirdly, we highlighted the enhanced DNAmAge-cognition associations resulting from PC-training compared to original epigenetic clocks. Fourthly, we discerned variations in domain-specific predictive capabilities among first-, second-, and third-generation DNAmAges in the context of brain ageing. Moreover, our study emphasized the significance of DNAmAges by comparing them to established risk factors for brain ageing. Lastly, we conducted detailed subgroup analyses based on social demographics, vascular and genetic risk factors, and the presence of brain WMH, employing both subgroup and formal interaction analyses.

Our study has also certain weaknesses. Firstly, our exploratory analysis involved multiple tests, increasing the risk of false positives, even after FDR correction. Secondly, some associations were unexpected and mostly nominally significant, necessitating cautious interpretation. Thirdly, longitudinal WCST analysis revealed an uncorrectable learning effect that may introduce bias. Fourthly, the use of 1.5T MRI didn’t allow for to investigate microstructural changes in the brain. Lastly, the relatively short follow-up duration underscores the need for longer-term studies in the future.

In conclusion, we show that DNAmAges are significant predictors of cognitive performance and more so of cognitive decline even within a short follow-up period of three to six years. Their effect is independent of well-known risk factors, surprisingly explaining more variability than many of those including BMI, diabetes or *ApoE4*-carrier status. Importantly, we observed highly significant and clinically relevant interactions between DNAmAges and hypertension as well as *ApoE4*-carrier status that require further explorations of their underlying mechanisms. None of the available DNAmAges captured all aspects of brain ageing highlighting the need for the development of brain-specific methylation clocks.

## Methods

### ASPS cohort

The ASPS study is a single-centre prospective cohort investigating the impact of vascular risk factors on brain structure and function in the elderly population of Graz city^18,19^. Extensive clinical assessments were performed, including brain MRI, neuropsychological tests, blood tests, blood pressure measurements, ECG, and echocardiography. The present study analysed a subset of 795 participants with DNA methylation, MRI, and cognitive test data. Ethics approval and informed consent were obtained.

### DNA methylation

DNA was extracted from EDTA whole peripheral blood using the phenol-chloroform method and quality was checked using nanodrop and gel electrophoresis. DNAs meeting A260/280 of 1.7-2.0 and displaying high molecular weight bands on gel electrophoresis were chosen for methylation analysis. A total of 750 ng of DNA from each sample was bisulfite treated using the Zymo EZ-96 DNA-methylation kit followed by whole genome amplification, fragmentation, and hybridization to the Illumina Infinium Methylation EPIC Bead Chip array following the manufacturer’s protocol (Illumina Inc., San Diego, CA). DNA methylation analysis was performed at the Human Genomics Facility of the Genetic Laboratory at Erasmus MC, Rotterdam, the Netherlands (http://www.glimdna.org/).

### Data pre-processing pipelines

To assess the impact of pre-processing on DNAmAge estimates, we analysed six technical replicates using two pipelines: minfi^20^ and ewastools^21^. In the ewastools pipeline, default pre-processing steps were followed, including probe removal based on high detection p-values (>0.01) and dye-bias correction. Normalisation was skipped as recommended by the package authors ^21^. In the minfi pipeline, probes with detection p-values >0.05 and samples with >5% of CpG sites having detection p-values >0.05 were removed and data was normalized using the Noob method. Samples were excluded based on sex mismatch (N=8) and failed quality control checks (N=1). We measured the average absolute agreement between six replicate pairs for 11 DNAmAges using intra-class correlation (ICC). We employed a suitable two-way random effect model for ICC estimation from the psych R package.

### Epigenetic clocks estimations

Pre-processed data was uploaded to Horvath’s online calculator (https://dnamage.genetics.ucla.edu/) for estimating Horvath, Hannum, Pheno, and Grim DNAmAges, along with intrinsic (IEAA) and extrinsic (EEAA) epigenetic age acceleration. The Horvath is a multi-tissue clock based on 353 CpGs^3^ and Hannum is a blood-based clock derived from 71 CpGs^4^. These clocks were developed to estimate chronological age based on DNA methylation across the genome. IEAA is a residual from a regression of Horvath DNAmAge on chronological age, and blood cell count estimates. EEAA is a residual obtained by regressing a weighted average of Hannum DNAmAge and estimated measures of specific immune cell types against chronological age. EEAA captures both age-related changes in blood cell composition and intrinsic epigenetic alterations^37^. PhenoAge is an estimate of biological age developed using a two-step approach; first calculating a weighted average of 10 clinical traits and regressing them against blood DNA methylation, revealing 513 CpG biomarkers^38^. GrimAge predicts lifespan by regressing surrogate DNA methylation biomarkers of plasma proteins and smoking pack years against time-to-death due to all-cause mortality, derived from 1030 CpG sites^7^.

Zhang et al. introduced age predictors in 2019, using Elastic Net (Zhang-ENP) and Best Linear Unbiased Prediction (Zhang-BLUP) algorithms on >13,000 samples. Zhang-ENP is based on 514 CpG sites, while Zhang BLUP involves 319,607 CpG sites for age estimation^35^ (https://github.com/qzhang314/DNAm-based-age-predictor/tree/v1.0.0/).

We also used the newly proposed principal component-based (PC-based) PCHorvath, PCHannum, PCPheno and PCGrim DNAmAges^39^ that are the proxies of the original Horvath, Hannum, Pheno and Grim DNAmAges (https://github.com/MorganLevineLab/PC-Clocks). Instead of CpGs, these DNAmAges are calculated using principal components (PCs) of CpGs to reduce technical These proxies were based on varying numbers of PCs: PCHorvath (121 PCs), PCHannum (390 PCs), PCPheno (652 PCs), and PCGrim (1936 PCs).

DunedinPACE is one of the newest DNAmAge that measures the rate of ageing over a specific 20-year period. It was developed using longitudinal organ-system integrity data of 19 biomarkers across physiological systems and a DNA methylation algorithm with 173 selected CpG sites^8^ (https://github.com/danbelsky/DunedinPACE).

### Cognition

ASPS participants underwent a comprehensive battery of cognitive assessment tests. Global cognitive assessment was performed using the Mini-Mental State Examination (MMSE) and Mattis Dementia Rating Scale (MDRS).

We used trail making test part B (TMT-B), Wisconsin card sorting test (WCST), Purdue pegboard test (PPT), Bäumler’s Lern-und Gedächtnistest and computerised complex reaction time task to evaluate various cognitive domains^40^.

Executive function was evaluated using the Trail Making Test part B (TMT-B) and the Wisconsin Card Sorting Test (WCST). In TMT-B, individuals quickly connect alternating numbers and letters in alternating sequences (e.g., 1-A-2-B-3-C, and so on) swiftly. The time in seconds is measured to correctly connect all the numbers and letters in the specified sequence. The WCST involves sorting cards based on specific rules, which change during the test. We combined categories and perseverative errors, yielding a WCST score.

Manual dexterity was assessed using the Purdue Pegboard Test (PPT) components: two-handed performance (RLB) and assembly. The PPT consists of a board with multiple holes and pegs of different shapes and sizes. We used two-handed performance (RLB) and assembly measures from PPT.

Bäumler’s Lern-und Gedächtnistest was used to evaluate performances in visual and verbal memory. Two subsets (tail and design recall) screen for visual memory whereas verbal memory is evaluated by three subsets (word and digit association tasks, and story recall) (Schmidt et al. 1999). The processing speed was assessed using a computerised complex reaction time task (Wiener Reaktionsgerät) that tested subject’s ability to react selectively by pressing a button as quickly as possible when a specific combination of a visual and acoustic signal appears ^40^.

Performance on the time variable from TMT-B and the reaction time task, as well as perseverative errors from WCST, is considered better when lower. To ensure uniformity, the scores were transformed to higher-is-better by adding 1 to the maximum value within each dataset and then subtracting the respective variable’s value. This adjustment aligned the polarity of all performance measures in the same direction.

### MRI markers

MRI marker estimation details have been previously published^18,40^. In brief, all MRI scans were conducted using 1.5T scanners with proton density-and T2-weighted sequences. Brain parenchymal fraction (BPF) represents the ratio of brain parenchymal volume to total intracranial volume. White matter hyperintensity (WMH) load in cubic millimetres represents the summation of individual lesion volumes. The WMH score was determined using the Fazekas scale for deep white matter changes, wherein scores range from 0 (no white matter change) to 3 (large confluent areas)^18,40^. WMH volume was normal log-transformed before the analysis.

### Covariates

Hypertension was categorised as present when there was a history of hypertension, hypertension medication, or mean systolic blood pressure ≥ 140mm Hg or mean diastolic pressure ≥ 90mm Hg. Diabetes was defined as a history of diabetes, use of antidiabetic treatment, or fasting blood sugar level > 140 mg/dl. The presence of cardiovascular disease (CVD) was identified based on cardiac abnormalities, coronary heart disease evidence, appropriate ECG findings, or signs of left ventricular hypertrophy. Education in years was the number of years of schooling, including university and higher education programs. Smoking status was categorised as never, former, or current smoker based on self-reported information. *ApoE* genotyping was performed using PCR-RFLP to determine the presence of the E4 allele, which was graded as present, or absent.

### Subgroup analysis

We divided our cohort into subgroups based on age (Young: ≤65years/Old: >65years), sex (Male/Female), hypertension (Normotensive/Hypertensive), BMI (normal weight: <25 Kg/m2/overweight: ≥25 kg/m2), education (Low education: ≤10years/ High education: >10years), CVD (No/Yes), diabetes (No/Yes), Smoking (Never/Former/Current), WMH scores(WMHSC 0 & 1/ WMHSC 2 & 3) and ApoE4 allele (Absent/Present). The age subgroup was made by dividing the cohort based on median age. We chose Zhang-BLUP, PCPheno, Grim, and DunedinPACE AA due to their outstanding performance within their respective generations

### Interaction analysis

We conducted a formal assessment of the impact of risk factors on the relationship between DNAmAges and cognitive function, utilising interaction terms. In order to streamline our analysis and reduce the number of statistical tests due to the numerous subgroups involved, we identified the most effective predictor of cognitive performance and focused on the subgroup where the effect size exhibited more than a twofold increase compared to others. Nevertheless, we conducted *ApoE4-*specific interaction tests in all phenotypes, utilizing all four epigenetic clocks.

### Statistical analysis

Epigenetic age acceleration (AA) is the residual resulting from regressing DNAm Ages on chronological age. We used AA measures in regression models to test the marker of the ageing hypothesis. We calculated per-year cognitive change for longitudinal analysis using linear mixed models adjusted for baseline age allowing individual specific intercepts and slopes. We deployed two nested linear regression models to test the association of phenotype of interest with various AAs. The first model was adjusted for baseline chronological age, sex and education. In the second model, in addition, we adjusted for vascular risk factors, namely hypertension, diabetes, presence of CVD, BMI, total cholesterol levels, smoking status and *Apoe4* allele. The estimates of white blood cell type abundance included levels of CD8T, CD4T, natural killer, monocytes, neutrophils and B cells and were calculated from DNA methylation data using Salas method ^41^. All analyses were conducted in R version 4.0.4 using the stats, lme4, tidyverse and psych packages in RStudio^42^. Partial R square was calculated using “pmvd” and “lmg” methods from relaimpo R package^43^.

## Data Availability

All data produced in the present study are available upon reasonable request to the authors

## Notes

### Competing Interest Statement

The authors have declared no competing interest.

### Funding Statement

The research reported in this article was funded by the Austrian National Bank Anniversary Fund, P15435, City Graz, Graz, Austria and, the Austrian Ministry of Science under the aegis of the EU Joint Programme - Neurodegenerative Disease Research - www.jpnd.eu. The project is supported through the following funding organizations under the aegis of EU Joint Programme - Neurodegenerative Disease Research - www.jpnd.eu: Australia, National Health and Medical Research Council, Austria, Federal Ministry of Science, Research and Economy; Canada, Canadian Institutes of Health Research; France, French National Research Agency; Germany, Federal Ministry of Education and Research; Netherlands, The Netherlands Organisation for Health Research and Development; United Kingdom, Medical Research Council. This project has received funding from the European Unions Horizon 2020 research and innovation program under grant agreement no. 643417.

### Author Declarations

The Medical Ethics Committee of Karl- Franzens University of Graz gave ethical approval for The Austrian Stroke Prevention Study. Written informed consent was obtained from all study participants.

