## Supplementary tables for "Epigenetic age acceleration is related to cognition and cognitive decline in the Elderly: Results of the Austrian Stroke Prevention Study"

**Supplementary Table 1:Intraclass correlations among six technical replicates**

| | | | EWAStools | | | | Minfi Noob | | | |
|---|---|---|---|---|---|---|---|---|---|---|
| **Intraclass correlation** | **Type** | **Epigenetic Age** | **ICC** | **p value** | **lower** | **upper** | **ICC** | **p value** | **lower** | **upper** |
| Single_raters_absolute | ICC1 | Horvath | 0.95 | 0.0002 | 0.74 | 0.99 | 0.92 | 0.0006 | 0.61 | 0.99 |
| Single_random_raters | **ICC2** | **Horvath** | **0.95** | **0.0005** | **0.74** | **0.99** | **0.92** | **0.0016** | **0.61** | **0.99** |
| Single_fixed_raters | ICC3 | Horvath | 0.95 | 0.0005 | 0.7 | 0.99 | 0.92 | 0.0016 | 0.55 | 0.99 |
| Average_raters_absolute | ICC1k | Horvath | 0.98 | 0.0002 | 0.85 | 1 | 0.96 | 0.0006 | 0.76 | 0.99 |
| Average_random_raters | ICC2k | Horvath | 0.98 | 0.0005 | 0.85 | 1 | 0.96 | 0.0016 | 0.76 | 0.99 |
| Average_fixed_raters | ICC3k | Horvath | 0.98 | 0.0005 | 0.82 | 1 | 0.96 | 0.0016 | 0.71 | 0.99 |
| Single_raters_absolute | ICC1 | PCHorvath | 0.96 | 6E-05 | 0.81 | 0.99 | 0.97 | 5E-05 | 0.81 | 1 |
| Single_random_raters | **ICC2** | **PCHorvath** | **0.96** | **0.0002** | **0.81** | **0.99** | **0.97** | **0.0002** | **0.81** | **1** |
| Single_fixed_raters | ICC3 | PCHorvath | 0.96 | 0.0002 | 0.77 | 1 | 0.97 | 0.0002 | 0.79 | 1 |
| Average_raters_absolute | ICC1k | PCHorvath | 0.98 | 6E-05 | 0.89 | 1 | 0.98 | 5E-05 | 0.9 | 1 |
| Average_random_raters | ICC2k | PCHorvath | 0.98 | 0.0002 | 0.89 | 1 | 0.98 | 0.0002 | 0.9 | 1 |
| Average_fixed_raters | ICC3k | PCHorvath | 0.98 | 0.0002 | 0.87 | 1 | 0.98 | 0.0002 | 0.88 | 1 |
| Single_raters_absolute | ICC1 | Hannum | 0.62 | 0.052 | -0.163 | 0.94 | 0.62 | 0.051 | -0.16 | 0.94 |
| Single_random_raters | **ICC2** | **Hannum** | **0.64** | **0.043** | **-0.068** | **0.94** | **0.64** | **0.037** | **-0.061** | **0.94** |
| Single_fixed_raters | ICC3 | Hannum | 0.69 | 0.043 | -0.136 | 0.95 | 0.71 | 0.037 | -0.094 | 0.95 |
| Average_raters_absolute | ICC1k | Hannum | 0.77 | 0.052 | -0.39 | 0.97 | 0.77 | 0.051 | -0.382 | 0.97 |
| Average_random_raters | ICC2k | Hannum | 0.78 | 0.043 | -0.145 | 0.97 | 0.78 | 0.037 | -0.129 | 0.97 |
| Average_fixed_raters | ICC3k | Hannum | 0.82 | 0.043 | -0.314 | 0.97 | 0.83 | 0.037 | -0.207 | 0.98 |
| Single_raters_absolute | ICC1 | PCHannum | 0.92 | 0.0008 | 0.58 | 0.99 | 0.90 | 0.0012 | 0.53 | 0.99 |
| Single_random_raters | **ICC2** | **PCHannum** | **0.92** | **0.0019** | **0.58** | **0.99** | **0.90** | **0.0027** | **0.53** | **0.99** |
| Single_fixed_raters | ICC3 | PCHannum | 0.92 | 0.0019 | 0.52 | 0.99 | 0.90 | 0.0027 | 0.47 | 0.99 |
| Average_raters_absolute | ICC1k | PCHannum | 0.96 | 0.0008 | 0.74 | 0.99 | 0.95 | 0.0012 | 0.7 | 0.99 |
| Average_random_raters | ICC2k | PCHannum | 0.96 | 0.0019 | 0.74 | 0.99 | 0.95 | 0.0027 | 0.69 | 0.99 |
| Average_fixed_raters | ICC3k | PCHannum | 0.96 | 0.0019 | 0.69 | 0.99 | 0.95 | 0.0027 | 0.64 | 0.99 |
| Single_raters_absolute | ICC1 | Zhang ENP | 0.88 | 0.0024 | 0.44 | 0.98 | 0.87 | 0.0028 | 0.41 | 0.98 |
| Single_random_raters | **ICC2** | **Zhang ENP** | **0.88** | **0.0048** | **0.44** | **0.98** | **0.87** | **0.0056** | **0.41** | **0.98** |
| Single_fixed_raters | ICC3 | Zhang ENP | 0.88 | 0.0048 | 0.36 | 0.98 | 0.87 | 0.0056 | 0.33 | 0.98 |
| Average_raters_absolute | ICC1k | Zhang ENP | 0.93 | 0.0024 | 0.61 | 0.99 | 0.93 | 0.0028 | 0.58 | 0.99 |

| | | | | | | | | | | |
|---|---|---|---|---|---|---|---|---|---|---|
| Average_random_raters | ICC2k | Zhang ENP | 0.93 | 0.0048 | 0.61 | 0.99 | 0.93 | 0.0056 | 0.58 | 0.99 |
| Average_fixed_raters | ICC3k | Zhang ENP | 0.93 | 0.0048 | 0.53 | 0.99 | 0.93 | 0.0056 | 0.5 | 0.99 |
| Single_raters_absolute | ICC1 | Zhang BLUP | 0.94 | 0.0003 | 0.68 | 0.99 | 0.94 | 0.0002 | 0.7 | 0.99 |
| Single_random_raters | **ICC2** | **Zhang BLUP** | **0.94** | **0.0009** | **0.68** | **0.99** | **0.94** | **0.0007** | **0.7** | **0.99** |
| Single_fixed_raters | ICC3 | Zhang BLUP | 0.94 | 0.0009 | 0.63 | 0.99 | 0.94 | 0.0007 | 0.65 | 0.99 |
| Average_raters_absolute | ICC1k | Zhang BLUP | 0.97 | 0.0003 | 0.81 | 1 | 0.97 | 0.0002 | 0.83 | 1 |
| Average_random_raters | ICC2k | Zhang BLUP | 0.97 | 0.0009 | 0.81 | 1 | 0.97 | 0.0007 | 0.83 | 1 |
| Average_fixed_raters | ICC3k | Zhang BLUP | 0.97 | 0.0009 | 0.77 | 1 | 0.97 | 0.0007 | 0.79 | 1 |
| Single_raters_absolute | ICC1 | Pheno | 0.78 | 0.0119 | 0.15 | 0.97 | 0.78 | 0.0119 | 0.15 | 0.97 |
| Single_random_raters | **ICC2** | **Pheno** | **0.79** | **0.0079** | **0.12** | **0.97** | **0.79** | **0.0079** | **0.12** | **0.97** |
| Single_fixed_raters | ICC3 | Pheno | 0.85 | 0.0079 | 0.26 | 0.98 | 0.85 | 0.0079 | 0.26 | 0.98 |
| Average_raters_absolute | ICC1k | Pheno | 0.88 | 0.0119 | 0.27 | 0.98 | 0.88 | 0.0119 | 0.27 | 0.98 |
| Average_random_raters | ICC2k | Pheno | 0.88 | 0.0079 | 0.21 | 0.98 | 0.88 | 0.0079 | 0.21 | 0.98 |
| Average_fixed_raters | ICC3k | Pheno | 0.92 | 0.0079 | 0.41 | 0.99 | 0.92 | 0.0079 | 0.41 | 0.99 |
| Single_raters_absolute | ICC1 | PCPheno | 0.92 | 0.0006 | 0.62 | 0.99 | 0.91 | 0.0009 | 0.56 | 0.99 |
| Single_random_raters | **ICC2** | **PCPheno** | **0.92** | **0.0015** | **0.62** | **0.99** | **0.91** | **0.0022** | **0.56** | **0.99** |
| Single_fixed_raters | ICC3 | PCPheno | 0.92 | 0.0015 | 0.56 | 0.99 | 0.91 | 0.0022 | 0.5 | 0.99 |
| Average_raters_absolute | ICC1k | PCPheno | 0.96 | 0.0006 | 0.76 | 0.99 | 0.95 | 0.0009 | 0.72 | 0.99 |
| Average_random_raters | ICC2k | PCPheno | 0.96 | 0.0015 | 0.76 | 0.99 | 0.95 | 0.0022 | 0.72 | 0.99 |
| Average_fixed_raters | ICC3k | PCPheno | 0.96 | 0.0015 | 0.72 | 0.99 | 0.95 | 0.0022 | 0.66 | 0.99 |
| Single_raters_absolute | ICC1 | Grim | 0.90 | 0.0012 | 0.53 | 0.99 | 0.95 | 0.0001 | 0.75 | 0.99 |
| Single_random_raters | **ICC2** | **Grim** | **0.90** | **0.0027** | **0.53** | **0.99** | **0.95** | **0.0003** | **0.73** | **0.99** |
| Single_fixed_raters | ICC3 | Grim | 0.90 | 0.0027 | 0.46 | 0.99 | 0.96 | 0.0003 | 0.75 | 0.99 |
| Average_raters_absolute | ICC1k | Grim | 0.95 | 0.0012 | 0.69 | 0.99 | 0.98 | 0.0001 | 0.86 | 1 |
| Average_random_raters | ICC2k | Grim | 0.95 | 0.0027 | 0.69 | 0.99 | 0.98 | 0.0003 | 0.85 | 1 |
| Average_fixed_raters | ICC3k | Grim | 0.95 | 0.0027 | 0.63 | 0.99 | 0.98 | 0.0003 | 0.86 | 1 |
| Single_raters_absolute | ICC1 | PCGrim | 1 | 5E-08 | 0.98 | 1 | 1 | 2E-08 | 0.98 | 1 |
| Single_random_raters | **ICC2** | **PCGrim** | **1** | **8E-08** | **0.92** | **1** | **1** | **2E-07** | **0.98** | **1** |
| Single_fixed_raters | ICC3 | PCGrim | 1 | 8E-08 | 0.99 | 1 | 1 | 2E-07 | 0.98 | 1 |
| Average_raters_absolute | ICC1k | PCGrim | 1 | 5E-08 | 0.99 | 1 | 1 | 2E-08 | 0.99 | 1 |
| Average_random_raters | ICC2k | PCGrim | 1 | 8E-08 | 0.96 | 1 | 1 | 2E-07 | 0.99 | 1 |

| | | | | | | | | | | |
|---|---|---|---|---|---|---|---|---|---|---|
| Average_fixed_raters | ICC3k | PCGrim | 1 | 8E-08 | 0.99 | 1 | 1 | 2E-07 | 0.99 | 1 |
| Single_raters_absolute | ICC1 | DunedinPACE | 0.89 | 0.0018 | 0.47 | 0.98 | 0.84 | 0.0045 | 0.33 | 0.98 |
| Single_random_raters | **ICC2** | **DunedinPACE** | **0.89** | **0.0027** | **0.47** | **0.98** | **0.84** | **0.0084** | **0.33** | **0.98** |
| Single_fixed_raters | ICC3 | DunedinPACE | 0.9 | 0.0027 | 0.46 | 0.99 | 0.84 | 0.0084 | 0.25 | 0.98 |
| Average_raters_absolute | ICC1k | DunedinPACE | 0.94 | 0.0018 | 0.64 | 0.99 | 0.92 | 0.0045 | 0.5 | 0.99 |
| Average_random_raters | ICC2k | DunedinPACE | 0.94 | 0.0027 | 0.64 | 0.99 | 0.92 | 0.0084 | 0.5 | 0.99 |
| Average_fixed_raters | ICC3k | DunedinPACE | 0.95 | 0.0027 | 0.63 | 0.99 | 0.92 | 0.0084 | 0.4 | 0.99 |

**Supplementary Table 2: Correlations between epigenetic age and age acceleration**

| | age_BL | Horvath | PCHorvath | Hannum | PCHannum | Zhang-ENP | Zhang-BLUP | Pheno | PCPheno | Grim | PCGrim | DunedinPACE | Horvath_AA | PCHorvath_AA | Hannum_AA | EEAA | IEAA | Zhang-ENP_AA | Zhang-BLUP_AA | Pheno_AA | PCPheno_AA | Grim_AA | PCGrim_AA | DunedinPACE_AA |
|---|---|---|---|---|---|---|---|---|---|---|---|---|---|---|---|---|---|---|---|---|---|---|---|---|
| age_BL | 1 | 0.64 | 0.62 | 0.71 | 0.66 | 0.82 | 0.83 | 0.62 | 0.7 | 0.81 | 0.89 | 0.18 | 0 | 0 | 0 | -0.01 | -0.02 | 0 | 0 | 0 | 0 | 0 | 0 | 0 |
| Horvath | 0.64 | 1 | 0.84 | 0.81 | 0.85 | 0.81 | 0.81 | 0.76 | 0.8 | 0.62 | 0.67 | 0.29 | 0.76 | 0.56 | 0.51 | 0.48 | 0.72 | 0.49 | 0.5 | 0.46 | 0.48 | 0.16 | 0.2 | 0.18 |
| PCHorvath | 0.62 | 0.84 | 1 | 0.82 | 0.96 | 0.81 | 0.8 | 0.74 | 0.9 | 0.65 | 0.71 | 0.36 | 0.57 | 0.78 | 0.54 | 0.52 | 0.5 | 0.52 | 0.51 | 0.45 | 0.62 | 0.25 | 0.34 | 0.25 |
| Hannum | 0.71 | 0.81 | 0.82 | 1 | 0.89 | 0.9 | 0.87 | 0.74 | 0.8 | 0.68 | 0.74 | 0.31 | 0.46 | 0.48 | 0.7 | 0.67 | 0.39 | 0.54 | 0.5 | 0.39 | 0.48 | 0.18 | 0.23 | 0.19 |
| PCHannum | 0.66 | 0.85 | 0.96 | 0.89 | 1 | 0.85 | 0.85 | 0.75 | 0.9 | 0.67 | 0.73 | 0.36 | 0.55 | 0.7 | 0.59 | 0.58 | 0.47 | 0.53 | 0.55 | 0.43 | 0.61 | 0.23 | 0.31 | 0.24 |
| Zhang-ENP | 0.82 | 0.81 | 0.81 | 0.9 | 0.85 | 1 | 0.94 | 0.77 | 0.9 | 0.73 | 0.79 | 0.25 | 0.37 | 0.39 | 0.45 | 0.42 | 0.33 | 0.58 | 0.47 | 0.33 | 0.39 | 0.13 | 0.14 | 0.1 |
| Zhnag-BLUP | 0.83 | 0.81 | 0.8 | 0.87 | 0.85 | 0.94 | 1 | 0.75 | 0.8 | 0.74 | 0.8 | 0.25 | 0.36 | 0.36 | 0.39 | 0.36 | 0.33 | 0.45 | 0.55 | 0.31 | 0.36 | 0.11 | 0.12 | 0.1 |
| Pheno | 0.62 | 0.76 | 0.74 | 0.74 | 0.75 | 0.77 | 0.75 | 1 | 0.8 | 0.68 | 0.69 | 0.41 | 0.48 | 0.45 | 0.43 | 0.42 | 0.43 | 0.45 | 0.43 | 0.79 | 0.51 | 0.31 | 0.31 | 0.31 |
| PCPheno | 0.7 | 0.79 | 0.88 | 0.84 | 0.9 | 0.85 | 0.84 | 0.8 | 1 | 0.73 | 0.79 | 0.43 | 0.45 | 0.57 | 0.49 | 0.5 | 0.38 | 0.48 | 0.47 | 0.47 | 0.71 | 0.28 | 0.37 | 0.31 |
| Grim | 0.81 | 0.62 | 0.65 | 0.68 | 0.67 | 0.73 | 0.74 | 0.68 | 0.7 | 1 | 0.94 | 0.5 | 0.13 | 0.19 | 0.15 | 0.15 | 0.09 | 0.13 | 0.12 | 0.23 | 0.23 | 0.59 | 0.48 | 0.36 |
| PCGrim | 0.89 | 0.67 | 0.71 | 0.74 | 0.73 | 0.79 | 0.8 | 0.69 | 0.8 | 0.94 | 1 | 0.44 | 0.12 | 0.2 | 0.15 | 0.15 | 0.09 | 0.11 | 0.1 | 0.18 | 0.24 | 0.37 | 0.46 | 0.28 |
| DunedinPACE | 0.18 | 0.29 | 0.36 | 0.31 | 0.36 | 0.25 | 0.25 | 0.41 | 0.4 | 0.5 | 0.44 | 1 | 0.23 | 0.31 | 0.26 | 0.29 | 0.18 | 0.18 | 0.18 | 0.38 | 0.43 | 0.59 | 0.6 | 0.98 |
| Horvath_AA | 0 | 0.76 | 0.57 | 0.46 | 0.55 | 0.37 | 0.36 | 0.48 | 0.5 | 0.13 | 0.12 | 0.23 | 1 | 0.73 | 0.66 | 0.63 | 0.95 | 0.64 | 0.65 | 0.61 | 0.63 | 0.21 | 0.27 | 0.23 |
| PCHorvath_AA | 0 | 0.56 | 0.78 | 0.48 | 0.7 | 0.39 | 0.36 | 0.45 | 0.6 | 0.19 | 0.2 | 0.31 | 0.73 | 1 | 0.69 | 0.67 | 0.65 | 0.67 | 0.66 | 0.57 | 0.8 | 0.32 | 0.43 | 0.31 |
| Hannum_AA | 0 | 0.51 | 0.54 | 0.7 | 0.59 | 0.45 | 0.39 | 0.43 | 0.5 | 0.15 | 0.15 | 0.26 | 0.66 | 0.69 | 1 | 0.97 | 0.58 | 0.77 | 0.71 | 0.55 | 0.69 | 0.26 | 0.32 | 0.27 |
| EEAA | 0.0 | 0.48 | 0.52 | 0.67 | 0.58 | 0.42 | 0.36 | 0.42 | 0.5 | 0.15 | 0.15 | 0.29 | 0.63 | 0.67 | 0.97 | 1 | 0.52 | 0.74 | 0.67 | 0.55 | 0.71 | 0.27 | 0.35 | 0.3 |
| IEAA | 0.0 | 0.72 | 0.5 | 0.39 | 0.47 | 0.33 | 0.33 | 0.43 | 0.4 | 0.09 | 0.09 | 0.18 | 0.95 | 0.65 | 0.58 | 0.52 | 1 | 0.61 | 0.62 | 0.57 | 0.56 | 0.18 | 0.24 | 0.19 |
| Zhang-ENP_AA | 0 | 0.49 | 0.52 | 0.54 | 0.53 | 0.58 | 0.45 | 0.45 | 0.5 | 0.13 | 0.11 | 0.18 | 0.64 | 0.67 | 0.77 | 0.74 | 0.61 | 1 | 0.81 | 0.58 | 0.67 | 0.22 | 0.25 | 0.18 |
| Zhang-BLUP_AA | 0 | 0.5 | 0.51 | 0.5 | 0.55 | 0.47 | 0.55 | 0.43 | 0.5 | 0.12 | 0.1 | 0.18 | 0.65 | 0.66 | 0.71 | 0.67 | 0.62 | 0.81 | 1 | 0.55 | 0.65 | 0.2 | 0.23 | 0.18 |
| Pheno_AA | 0 | 0.46 | 0.45 | 0.39 | 0.43 | 0.33 | 0.31 | 0.79 | 0.5 | 0.23 | 0.18 | 0.38 | 0.61 | 0.57 | 0.55 | 0.55 | 0.57 | 0.58 | 0.55 | 1 | 0.65 | 0.4 | 0.4 | 0.39 |

| | | | | | | | | | | | | | | | | | | | | | | | | |
|---|---|---|---|---|---|---|---|---|---|---|---|---|---|---|---|---|---|---|---|---|---|---|---|---|
| PCPheno_AA | 0 | 0.48 | 0.62 | 0.48 | 0.61 | 0.39 | 0.36 | 0.51 | 0.7 | 0.23 | 0.24 | 0.43 | 0.63 | 0.8 | 0.69 | 0.71 | 0.56 | 0.67 | 0.65 | 0.65 | 1 | 0.39 | 0.52 | 0.43 |
| Grim_AA | 0 | 0.16 | 0.25 | 0.18 | 0.23 | 0.13 | 0.11 | 0.31 | 0.3 | 0.59 | 0.37 | 0.59 | 0.21 | 0.32 | 0.26 | 0.27 | 0.18 | 0.22 | 0.2 | 0.4 | 0.39 | 1 | 0.81 | 0.6 |
| PCGrim_AA | 0 | 0.2 | 0.34 | 0.23 | 0.31 | 0.14 | 0.12 | 0.31 | 0.4 | 0.48 | 0.46 | 0.6 | 0.27 | 0.43 | 0.32 | 0.35 | 0.24 | 0.25 | 0.23 | 0.4 | 0.52 | 0.81 | 1 | 0.61 |
| DunedinPACE_AA | 0 | 0.18 | 0.25 | 0.19 | 0.24 | 0.1 | 0.1 | 0.31 | 0.3 | 0.36 | 0.28 | 0.98 | 0.23 | 0.31 | 0.27 | 0.3 | 0.19 | 0.18 | 0.18 | 0.39 | 0.43 | 0.6 | 0.61 | 1 |

**Supplementary Table 3: Cross-sectional associations of epigenetic age acceleration with cognition**

| | | MODEL 1 | | | | | | MODEL 2 | | | | | |
|---|---|---|---|---|---|---|---|---|---|---|---|---|---|
| **PHENO** | **PREDICTOR** | **Beta** | **SE** | ***p value*** | ***p adj*** | ***part R2*** | ***R2:Model1*** | **Beta** | **SE** | ***p value*** | ***p adj*** | ***part R2*** | ***R2:Model2*** |
| MMSE | Horvath | 0.01 | 0.011 | 0.165 | 0.32 | 0.25% | 13.87% | 0.02 | 0.011 | 0.099 | 0.24 | 0.37% | 16.03% |
| MMSE | PCHorvath | 0.03 | 0.012 | 0.026 | 0.12 | 0.61% | 14.20% | 0.03 | 0.012 | 0.014 | 0.08 | 0.74% | 16.42% |
| MMSE | Hannum | 0.03 | 0.012 | 0.009 | 0.06 | 0.86% | 14.40% | 0.04 | 0.013 | 0.004 | 0.04 | 1.02% | 16.67% |
| MMSE | PCHannum | 0.02 | 0.012 | 0.116 | 0.27 | 0.33% | 13.93% | 0.02 | 0.013 | 0.074 | 0.21 | 0.41% | 16.09% |
| MMSE | EEAA | 0.02 | 0.010 | 0.032 | 0.13 | 0.63% | 14.16% | 0.03 | 0.011 | 0.016 | 0.09 | 0.77% | 16.39% |
| MMSE | IEAA | 0.02 | 0.011 | 0.150 | 0.30 | 0.26% | 13.89% | 0.02 | 0.011 | 0.102 | 0.24 | 0.35% | 16.02% |
| MMSE | ENP | 0.05 | 0.014 | 0.001 | 0.01 | 1.35% | 14.91% | 0.06 | 0.015 | 0.000 | 0.00 | 1.79% | 17.50% |
| MMSE | BLUP | 0.04 | 0.015 | 0.003 | 0.03 | 1.02% | 14.61% | 0.04 | 0.015 | 0.006 | 0.05 | 0.88% | 16.59% |
| MMSE | Pheno | 0.01 | 0.009 | 0.147 | 0.30 | 0.26% | 13.89% | 0.01 | 0.009 | 0.144 | 0.30 | 0.26% | 15.96% |
| MMSE | PCPheno | 0.03 | 0.011 | 0.019 | 0.10 | 0.67% | 14.26% | 0.03 | 0.012 | 0.020 | 0.10 | 0.67% | 16.34% |
| MMSE | Grim | 0.01 | 0.015 | 0.612 | 0.72 | 0.04% | 13.68% | 0.02 | 0.017 | 0.197 | 0.38 | 0.16% | 15.91% |
| MMSE | PCGrim | 0.01 | 0.020 | 0.692 | 0.76 | 0.02% | 13.67% | 0.03 | 0.024 | 0.229 | 0.39 | 0.16% | 15.88% |
| MMSE | DunedinPACE | 0.63 | 0.515 | 0.219 | 0.35 | 0.22% | 13.82% | 0.71 | 0.567 | 0.209 | 0.39 | 0.18% | 15.90% |
| MDRS | Horvath | -0.02 | 0.023 | 0.351 | 0.48 | 0.10% | 27.55% | -0.01 | 0.023 | 0.516 | 0.64 | 0.05% | 28.39% |
| MDRS | PCHorvath | 0.04 | 0.025 | 0.122 | 0.28 | 0.19% | 27.69% | 0.03 | 0.026 | 0.258 | 0.41 | 0.12% | 28.47% |
| MDRS | Hannum | 0.02 | 0.026 | 0.378 | 0.51 | 0.07% | 27.54% | 0.03 | 0.027 | 0.297 | 0.42 | 0.09% | 28.46% |
| MDRS | PCHannum | 0.04 | 0.026 | 0.105 | 0.26 | 0.20% | 27.71% | 0.04 | 0.027 | 0.170 | 0.34 | 0.15% | 28.53% |
| MDRS | EEAA | 0.02 | 0.022 | 0.328 | 0.46 | 0.08% | 27.56% | 0.02 | 0.023 | 0.335 | 0.46 | 0.07% | 28.44% |
| MDRS | IEAA | -0.03 | 0.024 | 0.168 | 0.32 | 0.20% | 27.65% | -0.03 | 0.024 | 0.280 | 0.41 | 0.13% | 28.46% |
| MDRS | ENP | 0.03 | 0.030 | 0.285 | 0.42 | 0.10% | 27.58% | 0.05 | 0.032 | 0.125 | 0.28 | 0.20% | 28.58% |
| MDRS | BLUP | 0.03 | 0.031 | 0.297 | 0.43 | 0.09% | 27.57% | 0.03 | 0.033 | 0.291 | 0.42 | 0.09% | 28.46% |
| MDRS | Pheno | 0.01 | 0.019 | 0.505 | 0.63 | 0.04% | 27.51% | 0.02 | 0.020 | 0.281 | 0.41 | 0.10% | 28.46% |
| MDRS | PCPheno | 0.02 | 0.024 | 0.384 | 0.51 | 0.06% | 27.54% | 0.01 | 0.024 | 0.719 | 0.80 | 0.01% | 28.36% |
| MDRS | Grim | 0.03 | 0.031 | 0.342 | 0.48 | 0.07% | 27.55% | 0.03 | 0.036 | 0.447 | 0.57 | 0.05% | 28.40% |

| | | | | | | | | | | | | | |
|---|---|---|---|---|---|---|---|---|---|---|---|---|---|
| MDRS | PCGrim | -0.01 | 0.043 | 0.878 | 0.91 | 0.00% | 27.47% | -0.02 | 0.050 | 0.630 | 0.72 | 0.03% | 28.37% |
| MDRS | DunedinPACE | 0.30 | 1.101 | 0.786 | 0.82 | 0.01% | 27.48% | -0.58 | 1.194 | 0.626 | 0.72 | 0.03% | 28.37% |
| TMT | Horvath | -0.10 | 0.363 | 0.773 | 0.82 | 0.01% | 24.67% | 0.04 | 0.362 | 0.920 | 0.94 | 0.00% | 25.6% |
| TMT | PCHorvath | 0.01 | 0.409 | 0.974 | 0.98 | 0.00% | 24.66% | 0.08 | 0.420 | 0.852 | 0.89 | 0.00% | 25.6% |
| TMT | Hannum | -0.27 | 0.422 | 0.516 | 0.63 | 0.05% | 24.71% | -0.12 | 0.433 | 0.786 | 0.84 | 0.01% | 25.6% |
| TMT | PCHannum | -0.21 | 0.416 | 0.610 | 0.72 | 0.03% | 24.69% | -0.13 | 0.425 | 0.752 | 0.82 | 0.01% | 25.6% |
| TMT | EEAA | -0.31 | 0.347 | 0.368 | 0.50 | 0.11% | 24.74% | -0.21 | 0.356 | 0.561 | 0.68 | 0.05% | 25.6% |
| TMT | IEAA | 0.00 | 0.381 | 0.993 | 0.99 | 0.00% | 24.66% | 0.11 | 0.381 | 0.778 | 0.84 | 0.01% | 25.6% |
| TMT | ENP | 0.34 | 0.491 | 0.489 | 0.62 | 0.04% | 24.71% | 0.55 | 0.503 | 0.273 | 0.41 | 0.11% | 25.7% |
| TMT | BLUP | 0.02 | 0.509 | 0.963 | 0.98 | 0.00% | 24.66% | 0.12 | 0.516 | 0.812 | 0.86 | 0.01% | 25.6% |
| TMT | Pheno | -0.11 | 0.308 | 0.722 | 0.78 | 0.01% | 24.68% | 0.04 | 0.315 | 0.901 | 0.92 | 0.00% | 25.6% |
| TMT | PCPheno | -0.10 | 0.380 | 0.787 | 0.82 | 0.01% | 24.67% | -0.03 | 0.386 | 0.946 | 0.95 | 0.00% | 25.6% |
| TMT | Grim | -0.61 | 0.491 | 0.213 | 0.35 | 0.21% | 24.82% | -0.31 | 0.570 | 0.581 | 0.69 | 0.05% | 25.6% |
| TMT | PCGrim | -1.22 | 0.684 | 0.074 | 0.22 | 0.44% | 24.98% | -0.87 | 0.787 | 0.267 | 0.41 | 0.22% | 25.7% |
| TMT | DunedinPACE | -40.54 | 17.247 | 0.019 | 0.10 | 0.70% | 25.20% | -32.50 | 18.912 | 0.086 | 0.23 | 0.51% | 25.9% |
| WCST | Horvath | -0.09 | 0.052 | 0.087 | 0.23 | 0.45% | 1.88% | -0.10 | 0.053 | 0.060 | 0.19 | 0.57% | 2.91% |
| WCST | PCHorvath | -0.20 | 0.060 | 0.001 | 0.01 | 1.59% | 2.87% | -0.24 | 0.062 | 0.000 | 0.00 | 2.11% | 4.36% |
| WCST | Hannum | -0.20 | 0.061 | 0.001 | 0.01 | 1.52% | 2.94% | -0.20 | 0.063 | 0.002 | 0.02 | 1.51% | 3.78% |
| WCST | PCHannum | -0.22 | 0.060 | 0.000 | 0.00 | 1.99% | 3.28% | -0.25 | 0.062 | 0.000 | 0.00 | 2.36% | 4.60% |
| WCST | EEAA | -0.18 | 0.050 | 0.000 | 0.00 | 1.80% | 3.23% | -0.17 | 0.051 | 0.001 | 0.01 | 1.78% | 4.00% |
| WCST | IEAA | -0.06 | 0.055 | 0.261 | 0.39 | 0.20% | 1.66% | -0.08 | 0.055 | 0.172 | 0.34 | 0.30% | 2.68% |
| WCST | ENP | -0.29 | 0.070 | 0.000 | 0.00 | 2.21% | 3.62% | -0.31 | 0.072 | 0.000 | 0.00 | 2.53% | 4.84% |
| WCST | BLUP | -0.27 | 0.072 | 0.000 | 0.00 | 1.89% | 3.32% | -0.31 | 0.074 | 0.000 | 0.00 | 2.32% | 4.75% |
| WCST | Pheno | -0.11 | 0.044 | 0.010 | 0.06 | 1.03% | 2.37% | -0.12 | 0.046 | 0.011 | 0.07 | 1.12% | 3.32% |
| WCST | PCPheno | -0.25 | 0.056 | 0.000 | 0.00 | 2.75% | 3.96% | -0.25 | 0.057 | 0.000 | 0.00 | 2.97% | 5.05% |
| WCST | Grim | -0.14 | 0.071 | 0.049 | 0.16 | 0.62% | 2.00% | -0.17 | 0.083 | 0.038 | 0.15 | 0.59% | 3.01% |
| WCST | PCGrim | -0.15 | 0.099 | 0.135 | 0.29 | 0.38% | 1.79% | -0.18 | 0.115 | 0.109 | 0.26 | 0.37% | 2.77% |

| | | | | | | | | | | | | | |
|---|---|---|---|---|---|---|---|---|---|---|---|---|---|
| WCST | DunedinPACE | -5.97 | 2.476 | 0.016 | 0.09 | 0.92% | 2.25% | -6.56 | 2.724 | 0.016 | 0.09 | 0.95% | 3.21% |
| PPT RLB | Horvath | 0.07 | 0.034 | 0.049 | 0.16 | 0.29% | 35.50% | 0.09 | 0.035 | 0.013 | 0.08 | 0.47% | 37.00% |
| PPT RLB | PCHorvath | 0.07 | 0.039 | 0.066 | 0.20 | 0.25% | 35.46% | 0.09 | 0.040 | 0.025 | 0.11 | 0.38% | 36.90% |
| PPT RLB | Hannum | 0.04 | 0.040 | 0.326 | 0.46 | 0.08% | 35.25% | 0.07 | 0.042 | 0.091 | 0.24 | 0.22% | 36.71% |
| PPT RLB | PCHannum | 0.06 | 0.040 | 0.164 | 0.32 | 0.14% | 35.33% | 0.08 | 0.041 | 0.063 | 0.19 | 0.26% | 36.76% |
| PPT RLB | EEAA | 0.04 | 0.033 | 0.268 | 0.40 | 0.10% | 35.27% | 0.06 | 0.034 | 0.099 | 0.24 | 0.21% | 36.70% |
| PPT RLB | IEAA | 0.05 | 0.036 | 0.139 | 0.30 | 0.17% | 35.36% | 0.07 | 0.037 | 0.043 | 0.15 | 0.33% | 36.82% |
| PPT RLB | ENP | 0.13 | 0.047 | 0.007 | 0.06 | 0.54% | 35.78% | 0.17 | 0.048 | 0.001 | 0.01 | 0.87% | 37.49% |
| PPT RLB | BLUP | 0.08 | 0.048 | 0.116 | 0.27 | 0.20% | 35.38% | 0.11 | 0.050 | 0.028 | 0.11 | 0.38% | 36.89% |
| PPT RLB | Pheno | 0.05 | 0.029 | 0.094 | 0.24 | 0.20% | 35.41% | 0.08 | 0.030 | 0.010 | 0.07 | 0.45% | 37.05% |
| PPT RLB | PCPheno | 0.06 | 0.036 | 0.083 | 0.23 | 0.22% | 35.42% | 0.07 | 0.037 | 0.045 | 0.15 | 0.29% | 36.81% |
| PPT RLB | Grim | -0.06 | 0.047 | 0.225 | 0.35 | 0.16% | 35.29% | 0.04 | 0.055 | 0.469 | 0.59 | 0.05% | 36.50% |
| PPT RLB | PCGrim | -0.08 | 0.065 | 0.193 | 0.35 | 0.21% | 35.31% | 0.03 | 0.076 | 0.684 | 0.77 | 0.02% | 36.47% |
| PPT RLB | DunedinPACE | -2.22 | 1.648 | 0.179 | 0.33 | 0.21% | 35.32% | -0.89 | 1.832 | 0.627 | 0.72 | 0.03% | 36.47% |
| PPT Assembly | Horvath | 0.02 | 0.038 | 0.514 | 0.63 | 0.03% | 34.85% | 0.04 | 0.038 | 0.310 | 0.44 | 0.08% | 36.32% |
| PPT Assembly | PCHorvath | 0.02 | 0.043 | 0.680 | 0.76 | 0.01% | 34.83% | 0.03 | 0.044 | 0.451 | 0.57 | 0.04% | 36.27% |
| PPT Assembly | Hannum | 0.02 | 0.044 | 0.696 | 0.76 | 0.01% | 34.83% | 0.05 | 0.046 | 0.266 | 0.41 | 0.09% | 36.33% |
| PPT Assembly | PCHannum | 0.02 | 0.043 | 0.618 | 0.72 | 0.02% | 34.84% | 0.04 | 0.045 | 0.333 | 0.46 | 0.07% | 36.31% |
| PPT Assembly | EEAA | 0.02 | 0.036 | 0.679 | 0.76 | 0.01% | 34.83% | 0.04 | 0.038 | 0.333 | 0.46 | 0.07% | 36.31% |
| PPT Assembly | IEAA | 0.01 | 0.040 | 0.772 | 0.82 | 0.01% | 34.82% | 0.02 | 0.040 | 0.583 | 0.69 | 0.03% | 36.25% |
| PPT Assembly | ENP | 0.09 | 0.051 | 0.097 | 0.25 | 0.21% | 35.05% | 0.12 | 0.053 | 0.025 | 0.11 | 0.36% | 36.68% |
| PPT Assembly | BLUP | 0.07 | 0.053 | 0.220 | 0.35 | 0.12% | 34.94% | 0.09 | 0.054 | 0.100 | 0.24 | 0.21% | 36.47% |
| PPT Assembly | Pheno | 0.02 | 0.032 | 0.499 | 0.63 | 0.04% | 34.85% | 0.05 | 0.033 | 0.115 | 0.26 | 0.17% | 36.45% |
| PPT Assembly | PCPheno | 0.02 | 0.040 | 0.684 | 0.76 | 0.01% | 34.83% | 0.03 | 0.041 | 0.449 | 0.57 | 0.04% | 36.27% |
| PPT Assembly | Grim | -0.06 | 0.051 | 0.222 | 0.35 | 0.18% | 34.94% | 0.06 | 0.060 | 0.357 | 0.48 | 0.07% | 36.30% |
| PPT Assembly | PCGrim | -0.11 | 0.071 | 0.120 | 0.28 | 0.33% | 35.02% | 0.01 | 0.083 | 0.882 | 0.91 | 0.00% | 36.22% |
| PPT Assembly | DunedinPACE | -4.73 | 1.803 | 0.009 | 0.06 | 0.88% | 35.40% | -2.95 | 2.004 | 0.141 | 0.30 | 0.52% | 36.42% |

| | | | | | | | | | | | | | |
|---|---|---|---|---|---|---|---|---|---|---|---|---|---|
| VSM | Horvath | -0.29 | 0.228 | 0.208 | 0.35 | 0.21% | 22.52% | -0.26 | 0.232 | 0.264 | 0.41 | 0.18% | 24.20% |
| VSM | PCHorvath | -0.37 | 0.255 | 0.144 | 0.30 | 0.29% | 22.58% | -0.31 | 0.267 | 0.248 | 0.40 | 0.21% | 24.21% |
| VSM | Hannum | -0.60 | 0.271 | 0.027 | 0.12 | 0.60% | 22.87% | -0.52 | 0.278 | 0.063 | 0.19 | 0.48% | 24.44% |
| VSM | PCHannum | -0.43 | 0.262 | 0.100 | 0.25 | 0.36% | 22.64% | -0.34 | 0.272 | 0.211 | 0.39 | 0.25% | 24.23% |
| VSM | EEAA | -0.50 | 0.223 | 0.024 | 0.11 | 0.63% | 22.89% | -0.44 | 0.228 | 0.057 | 0.19 | 0.51% | 24.46% |
| VSM | IEAA | -0.33 | 0.239 | 0.171 | 0.32 | 0.23% | 22.55% | -0.30 | 0.243 | 0.221 | 0.39 | 0.20% | 24.22% |
| VSM | ENP | -0.62 | 0.312 | 0.048 | 0.16 | 0.47% | 22.77% | -0.59 | 0.321 | 0.064 | 0.19 | 0.41% | 24.44% |
| VSM | BLUP | -0.69 | 0.323 | 0.033 | 0.13 | 0.53% | 22.83% | -0.68 | 0.329 | 0.040 | 0.15 | 0.52% | 24.52% |
| VSM | Pheno | -0.23 | 0.195 | 0.249 | 0.38 | 0.18% | 22.49% | -0.13 | 0.201 | 0.528 | 0.64 | 0.06% | 24.10% |
| VSM | PCPheno | -0.56 | 0.240 | 0.020 | 0.10 | 0.69% | 22.93% | -0.50 | 0.247 | 0.043 | 0.15 | 0.62% | 24.51% |
| VSM | Grim | -0.84 | 0.308 | 0.007 | 0.06 | 0.92% | 23.13% | -0.43 | 0.364 | 0.237 | 0.39 | 0.44% | 24.21% |
| VSM | PCGrim | -1.16 | 0.433 | 0.007 | 0.06 | 0.90% | 23.11% | -0.61 | 0.509 | 0.229 | 0.39 | 0.48% | 24.22% |
| VSM | DunedinPACE | -42.22 | 10.983 | 0.000 | 0.00 | 1.75% | 23.90% | -34.23 | 12.255 | 0.005 | 0.04 | 1.56% | 24.92% |
| VRM | Horvath | -0.14 | 0.174 | 0.428 | 0.55 | 0.07% | 29.32% | -0.12 | 0.178 | 0.483 | 0.60 | 0.06% | 31.14% |
| VRM | PCHorvath | -0.25 | 0.195 | 0.197 | 0.35 | 0.18% | 29.42% | -0.25 | 0.205 | 0.231 | 0.39 | 0.19% | 31.24% |
| VRM | Hannum | -0.36 | 0.207 | 0.085 | 0.23 | 0.31% | 29.54% | -0.27 | 0.214 | 0.203 | 0.39 | 0.22% | 31.26% |
| VRM | PCHannum | -0.25 | 0.200 | 0.206 | 0.35 | 0.18% | 29.41% | -0.23 | 0.209 | 0.279 | 0.41 | 0.16% | 31.21% |
| VRM | EEAA | -0.34 | 0.170 | 0.043 | 0.16 | 0.41% | 29.65% | -0.27 | 0.176 | 0.125 | 0.28 | 0.30% | 31.33% |
| VRM | IEAA | -0.16 | 0.182 | 0.394 | 0.51 | 0.08% | 29.32% | -0.15 | 0.187 | 0.417 | 0.55 | 0.08% | 31.16% |
| VRM | ENP | -0.42 | 0.237 | 0.080 | 0.23 | 0.32% | 29.55% | -0.29 | 0.246 | 0.233 | 0.39 | 0.21% | 31.24% |
| VRM | BLUP | -0.38 | 0.246 | 0.128 | 0.28 | 0.24% | 29.48% | -0.31 | 0.253 | 0.215 | 0.39 | 0.20% | 31.25% |
| VRM | Pheno | 0.01 | 0.149 | 0.943 | 0.97 | 0.00% | 29.25% | 0.04 | 0.155 | 0.778 | 0.84 | 0.01% | 31.10% |
| VRM | PCPheno | -0.34 | 0.183 | 0.061 | 0.19 | 0.37% | 29.60% | -0.33 | 0.189 | 0.080 | 0.22 | 0.37% | 31.40% |
| VRM | Grim | -0.14 | 0.235 | 0.557 | 0.66 | 0.04% | 29.29% | -0.10 | 0.280 | 0.732 | 0.81 | 0.01% | 31.11% |
| VRM | PCGrim | -0.47 | 0.330 | 0.159 | 0.31 | 0.22% | 29.45% | -0.56 | 0.391 | 0.153 | 0.31 | 0.23% | 31.30% |
| VRM | DunedinPACE | -5.33 | 8.442 | 0.528 | 0.64 | 0.05% | 29.29% | -4.08 | 9.459 | 0.666 | 0.76 | 0.02% | 31.11% |
| Speed | Horvath | 0.98 | 0.772 | 0.205 | 0.35 | 0.18% | 21.39% | 1.38 | 0.791 | 0.082 | 0.22 | 0.33% | 23.25% |

| | | | | | | | | | | | | | |
|---|---|---|---|---|---|---|---|---|---|---|---|---|---|
| Speed | PCHorvath | 2.10 | 0.864 | 0.015 | 0.09 | 0.70% | 21.86% | 2.44 | 0.901 | 0.007 | 0.05 | 0.82% | 23.73% |
| Speed | Hannum | 1.80 | 0.902 | 0.046 | 0.16 | 0.51% | 21.65% | 2.07 | 0.933 | 0.027 | 0.11 | 0.59% | 23.46% |
| Speed | PCHannum | 1.77 | 0.881 | 0.045 | 0.16 | 0.48% | 21.65% | 2.11 | 0.915 | 0.021 | 0.10 | 0.59% | 23.51% |
| Speed | EEAA | 1.43 | 0.740 | 0.053 | 0.17 | 0.50% | 21.62% | 1.69 | 0.768 | 0.028 | 0.11 | 0.60% | 23.45% |
| Speed | IEAA | 0.92 | 0.812 | 0.258 | 0.39 | 0.14% | 21.36% | 1.23 | 0.831 | 0.139 | 0.30 | 0.25% | 23.15% |
| Speed | ENP | 3.17 | 1.046 | 0.003 | 0.03 | 1.13% | 22.20% | 3.56 | 1.082 | 0.001 | 0.01 | 1.24% | 24.11% |
| Speed | BLUP | 3.10 | 1.077 | 0.004 | 0.04 | 0.98% | 22.11% | 3.55 | 1.110 | 0.001 | 0.02 | 1.13% | 24.05% |
| Speed | Pheno | 1.38 | 0.654 | 0.035 | 0.13 | 0.50% | 21.70% | 1.41 | 0.675 | 0.038 | 0.15 | 0.46% | 23.39% |
| Speed | PCPheno | 2.02 | 0.797 | 0.011 | 0.07 | 0.76% | 21.91% | 2.34 | 0.826 | 0.005 | 0.04 | 0.90% | 23.81% |
| Speed | Grim | 0.50 | 1.037 | 0.630 | 0.72 | 0.03% | 21.24% | 0.06 | 1.230 | 0.959 | 0.96 | 0.00% | 22.91% |
| Speed | PCGrim | 1.23 | 1.444 | 0.395 | 0.51 | 0.09% | 21.30% | 1.49 | 1.694 | 0.381 | 0.51 | 0.12% | 22.99% |
| Speed | DunedinPACE | -46.60 | 36.450 | 0.202 | 0.35 | 0.19% | 21.40% | -49.52 | 40.992 | 0.227 | 0.39 | 0.20% | 23.07% |

**Supplementary Table 4:Longitudinal associations of epigenetic age acceleration with cognition**

| | | MODEL 1 | | | | | | MODEL 2 | | | | | |
|---|---|---|---|---|---|---|---|---|---|---|---|---|---|
| **PHENO** | **PREDICTOR** | **Beta** | **SE** | ***p value*** | ***p adj*** | ***part R2*** | ***R2 Model1*** | **Beta** | **SE** | ***p value*** | ***p adj*** | ***part R2*** | ***R2 Model2*** |
| MMSE change | Horvath | 0.00 | 0.000 | 0.925 | 0.95 | 0.00% | 7.10% | 0.00 | 0.000 | 0.955 | 0.96 | 0.00% | 7.54% |
| MMSE change | PCHorvath | 0.00 | 0.000 | 0.780 | 0.87 | 0.01% | 7.11% | 0.00 | 0.000 | 0.769 | 0.91 | 0.02% | 7.56% |
| MMSE change | Hannum | 0.00 | 0.000 | 0.596 | 0.75 | 0.05% | 7.15% | 0.00 | 0.000 | 0.816 | 0.94 | 0.01% | 7.55% |
| MMSE change | PCHannum | 0.00 | 0.000 | 0.612 | 0.75 | 0.04% | 7.15% | 0.00 | 0.000 | 0.653 | 0.84 | 0.04% | 7.59% |
| MMSE change | EEAA | 0.00 | 0.000 | 0.616 | 0.75 | 0.04% | 7.15% | 0.00 | 0.000 | 0.823 | 0.94 | 0.01% | 7.55% |
| MMSE change | IEAA | 0.00 | 0.000 | 0.817 | 0.90 | 0.01% | 7.11% | 0.00 | 0.000 | 0.895 | 0.94 | 0.00% | 7.55% |
| MMSE change | ENP | 0.00 | 0.000 | 0.581 | 0.74 | 0.06% | 7.16% | 0.00 | 0.000 | 0.685 | 0.85 | 0.03% | 7.58% |
| MMSE change | BLUP | 0.00 | 0.000 | 0.737 | 0.83 | 0.02% | 7.12% | 0.00 | 0.001 | 0.848 | 0.94 | 0.01% | 7.55% |
| MMSE change | Pheno | 0.00 | 0.000 | 0.925 | 0.95 | 0.00% | 7.10% | 0.00 | 0.000 | 0.889 | 0.94 | 0.00% | 7.55% |
| MMSE change | PCPheno | 0.00 | 0.000 | 0.482 | 0.66 | 0.12% | 7.19% | 0.00 | 0.000 | 0.545 | 0.74 | 0.09% | 7.62% |
| MMSE change | Grim | 0.00 | 0.000 | 0.157 | 0.28 | 0.46% | 7.48% | 0.00 | 0.001 | 0.067 | 0.16 | 0.52% | 8.23% |
| MMSE change | PCGrim | 0.00 | 0.001 | 0.416 | 0.59 | 0.18% | 7.23% | 0.00 | 0.001 | 0.222 | 0.37 | 0.26% | 7.85% |
| MMSE change | DunedinPACE | -0.01 | 0.017 | 0.516 | 0.69 | 0.11% | 7.18% | -0.02 | 0.019 | 0.363 | 0.54 | 0.13% | 7.71% |
| MDRS change | Horvath | 0.00 | 0.002 | 0.579 | 0.74 | 0.06% | 5.16% | 0.00 | 0.002 | 0.338 | 0.51 | 0.14% | 6.87% |
| MDRS change | PCHorvath | 0.00 | 0.002 | 0.929 | 0.95 | 0.00% | 5.10% | 0.00 | 0.002 | 0.768 | 0.91 | 0.02% | 6.70% |
| MDRS change | Hannum | 0.00 | 0.002 | 0.903 | 0.95 | 0.00% | 5.11% | 0.00 | 0.002 | 0.633 | 0.82 | 0.04% | 6.73% |
| MDRS change | PCHannum | 0.00 | 0.002 | 0.927 | 0.95 | 0.00% | 5.10% | 0.00 | 0.002 | 0.742 | 0.91 | 0.02% | 6.70% |
| MDRS change | EEAA | 0.00 | 0.002 | 0.998 | 1.00 | 0.00% | 5.10% | 0.00 | 0.002 | 0.682 | 0.85 | 0.03% | 6.72% |
| MDRS change | IEAA | 0.00 | 0.002 | 0.511 | 0.69 | 0.08% | 5.19% | 0.00 | 0.002 | 0.344 | 0.52 | 0.15% | 6.87% |
| MDRS change | ENP | 0.00 | 0.002 | 0.485 | 0.66 | 0.10% | 5.20% | 0.00 | 0.002 | 0.766 | 0.91 | 0.03% | 6.70% |
| MDRS change | BLUP | 0.00 | 0.002 | 0.412 | 0.59 | 0.14% | 5.24% | 0.00 | 0.003 | 0.621 | 0.82 | 0.07% | 6.73% |
| MDRS change | Pheno | 0.00 | 0.001 | 0.630 | 0.75 | 0.05% | 5.15% | 0.00 | 0.002 | 0.877 | 0.94 | 0.01% | 6.69% |
| MDRS change | PCPheno | 0.00 | 0.002 | 0.211 | 0.36 | 0.38% | 5.41% | 0.00 | 0.002 | 0.391 | 0.57 | 0.24% | 6.83% |

| | | | | | | | | | | | | | |
|---|---|---|---|---|---|---|---|---|---|---|---|---|---|
| MDRS change | Grim | 0.00 | 0.002 | 0.413 | 0.59 | 0.17% | 5.24% | 0.00 | 0.003 | 0.280 | 0.46 | 0.25% | 6.92% |
| MDRS change | PCGrim | 0.00 | 0.003 | 0.535 | 0.70 | 0.10% | 5.18% | 0.00 | 0.004 | 0.428 | 0.61 | 0.15% | 6.81% |
| MDRS change | DunedinPACE | -0.09 | 0.084 | 0.295 | 0.45 | 0.29% | 5.32% | -0.08 | 0.095 | 0.420 | 0.61 | 0.18% | 6.81% |
| TMT change | Horvath | 0.00 | 0.011 | 0.820 | 0.90 | 0.01% | 4.81% | 0.00 | 0.011 | 0.848 | 0.94 | 0.01% | 12.2% |
| TMT change | PCHorvath | -0.02 | 0.012 | 0.056 | 0.13 | 0.81% | 5.52% | -0.02 | 0.012 | 0.078 | 0.18 | 0.65% | 12.8% |
| TMT change | Hannum | -0.03 | 0.012 | 0.032 | 0.09 | 0.94% | 5.71% | -0.02 | 0.012 | 0.111 | 0.22 | 0.60% | 12.7% |
| TMT change | PCHannum | -0.03 | 0.012 | 0.013 | 0.04 | 1.31% | 6.02% | -0.02 | 0.012 | 0.041 | 0.12 | 0.96% | 13.1% |
| TMT change | EEAA | -0.02 | 0.010 | 0.022 | 0.06 | 1.06% | 5.84% | -0.02 | 0.010 | 0.076 | 0.18 | 0.78% | 12.9% |
| TMT change | IEAA | 0.00 | 0.011 | 0.872 | 0.93 | 0.01% | 4.80% | 0.00 | 0.011 | 0.908 | 0.95 | 0.00% | 12.2% |
| TMT change | ENP | -0.03 | 0.014 | 0.043 | 0.10 | 0.83% | 5.61% | -0.02 | 0.014 | 0.114 | 0.22 | 0.52% | 12.7% |
| TMT change | BLUP | -0.04 | 0.014 | 0.009 | 0.03 | 1.34% | 6.13% | -0.03 | 0.014 | 0.032 | 0.10 | 0.96% | 13.1% |
| TMT change | Pheno | -0.01 | 0.009 | 0.268 | 0.43 | 0.28% | 5.04% | -0.01 | 0.009 | 0.523 | 0.73 | 0.09% | 12.3% |
| TMT change | PCPheno | -0.03 | 0.010 | 0.002 | 0.01 | 2.03% | 6.69% | -0.03 | 0.010 | 0.010 | 0.05 | 1.56% | 13.5% |
| TMT change | Grim | -0.04 | 0.014 | 0.003 | 0.02 | 1.83% | 6.49% | -0.04 | 0.016 | 0.019 | 0.06 | 1.11% | 13.3% |
| TMT change | PCGrim | -0.06 | 0.019 | 0.001 | 0.01 | 2.07% | 6.82% | -0.05 | 0.022 | 0.012 | 0.05 | 1.34% | 13.5% |
| TMT change | DunedinPACE | -1.37 | 0.494 | 0.006 | 0.03 | 1.69% | 6.29% | -1.06 | 0.541 | 0.051 | 0.13 | 0.97% | 13.0% |
| WCST change | Horvath | 0.01 | 0.007 | 0.141 | 0.26 | 0.52% | 2.55% | 0.01 | 0.007 | 0.100 | 0.21 | 0.65% | 5.00% |
| WCST change | PCHorvath | 0.02 | 0.007 | 0.003 | 0.02 | 1.97% | 3.89% | 0.03 | 0.007 | 0.001 | 0.01 | 2.58% | 6.85% |
| WCST change | Hannum | 0.02 | 0.008 | 0.002 | 0.01 | 2.03% | 4.10% | 0.02 | 0.008 | 0.004 | 0.03 | 2.02% | 6.17% |
| WCST change | PCHannum | 0.02 | 0.007 | 0.001 | 0.01 | 2.39% | 4.31% | 0.03 | 0.007 | 0.001 | 0.01 | 2.72% | 6.88% |
| WCST change | EEAA | 0.02 | 0.006 | 0.001 | 0.01 | 2.39% | 4.48% | 0.02 | 0.006 | 0.003 | 0.02 | 2.32% | 6.33% |
| WCST change | IEAA | 0.01 | 0.007 | 0.251 | 0.41 | 0.29% | 2.38% | 0.01 | 0.007 | 0.161 | 0.29 | 0.45% | 4.84% |
| WCST change | ENP | 0.03 | 0.009 | 0.001 | 0.01 | 2.34% | 4.44% | 0.03 | 0.009 | 0.000 | 0.01 | 2.75% | 7.23% |
| WCST change | BLUP | 0.03 | 0.009 | 0.001 | 0.01 | 2.26% | 4.37% | 0.03 | 0.009 | 0.000 | 0.01 | 2.77% | 7.28% |
| WCST change | Pheno | 0.01 | 0.006 | 0.016 | 0.05 | 1.36% | 3.30% | 0.01 | 0.006 | 0.017 | 0.06 | 1.51% | 5.64% |
| WCST change | PCPheno | 0.02 | 0.007 | 0.000 | 0.01 | 3.04% | 4.90% | 0.02 | 0.007 | 0.000 | 0.01 | 3.21% | 7.13% |
| WCST change | Grim | 0.02 | 0.009 | 0.021 | 0.06 | 1.27% | 3.18% | 0.02 | 0.010 | 0.022 | 0.07 | 1.29% | 5.55% |

| | | | | | | | | | | | | | |
|---|---|---|---|---|---|---|---|---|---|---|---|---|---|
| WCST change | PCGrim | 0.02 | 0.012 | 0.065 | 0.14 | 0.87% | 2.80% | 0.02 | 0.014 | 0.103 | 0.21 | 0.81% | 4.99% |
| WCST change | DunedinPACE | 0.83 | 0.314 | 0.008 | 0.03 | 1.69% | 3.52% | 0.86 | 0.345 | 0.013 | 0.05 | 1.80% | 5.73% |
| PPT RLB change | Horvath | 0.00 | 0.001 | 0.099 | 0.20 | 0.47% | 5.72% | 0.00 | 0.001 | 0.064 | 0.16 | 0.57% | 10.36% |
| PPT RLB change | PCHorvath | 0.00 | 0.001 | 0.078 | 0.16 | 0.53% | 5.80% | 0.00 | 0.001 | 0.073 | 0.17 | 0.59% | 10.32% |
| PPT RLB change | Hannum | 0.00 | 0.001 | 0.132 | 0.25 | 0.40% | 5.63% | 0.00 | 0.001 | 0.125 | 0.24 | 0.42% | 10.15% |
| PPT RLB change | PCHannum | 0.00 | 0.001 | 0.110 | 0.22 | 0.44% | 5.69% | 0.00 | 0.001 | 0.099 | 0.21 | 0.47% | 10.22% |
| PPT RLB change | EEAA | 0.00 | 0.001 | 0.077 | 0.16 | 0.54% | 5.80% | 0.00 | 0.001 | 0.093 | 0.20 | 0.49% | 10.24% |
| PPT RLB change | IEAA | 0.00 | 0.001 | 0.155 | 0.28 | 0.35% | 5.58% | 0.00 | 0.001 | 0.084 | 0.18 | 0.51% | 10.27% |
| PPT RLB change | ENP | 0.00 | 0.002 | 0.007 | 0.03 | 1.33% | 6.64% | 0.00 | 0.002 | 0.007 | 0.04 | 1.37% | 11.15% |
| PPT RLB change | BLUP | 0.00 | 0.002 | 0.306 | 0.46 | 0.21% | 5.39% | 0.00 | 0.002 | 0.126 | 0.24 | 0.41% | 10.14% |
| PPT RLB change | Pheno | 0.00 | 0.001 | 0.268 | 0.43 | 0.21% | 5.42% | 0.00 | 0.001 | 0.199 | 0.34 | 0.30% | 10.00% |
| PPT RLB change | PCPheno | 0.00 | 0.001 | 0.067 | 0.15 | 0.58% | 5.85% | 0.00 | 0.001 | 0.047 | 0.13 | 0.64% | 10.46% |
| PPT RLB change | Grim | 0.00 | 0.002 | 0.271 | 0.43 | 0.32% | 5.42% | 0.00 | 0.002 | 0.936 | 0.95 | 0.00% | 9.67% |
| PPT RLB change | PCGrim | 0.00 | 0.002 | 0.713 | 0.83 | 0.03% | 5.21% | 0.00 | 0.002 | 0.336 | 0.51 | 0.20% | 9.86% |
| PPT RLB change | DunedinPACE | 0.03 | 0.056 | 0.632 | 0.75 | 0.06% | 5.23% | 0.01 | 0.062 | 0.894 | 0.94 | 0.00% | 9.67% |
| PPT AS change | Horvath | 0.00 | 0.000 | 0.356 | 0.53 | 0.16% | 3.27% | 0.00 | 0.000 | 0.286 | 0.46 | 0.22% | 6.81% |
| PPT AS change | PCHorvath | 0.00 | 0.000 | 0.835 | 0.90 | 0.01% | 3.10% | 0.00 | 0.000 | 0.811 | 0.94 | 0.01% | 6.58% |
| PPT AS change | Hannum | 0.00 | 0.000 | 0.727 | 0.83 | 0.03% | 3.12% | 0.00 | 0.000 | 0.938 | 0.95 | 0.00% | 6.57% |
| PPT AS change | PCHannum | 0.00 | 0.000 | 0.828 | 0.90 | 0.01% | 3.10% | 0.00 | 0.000 | 0.684 | 0.85 | 0.03% | 6.61% |
| PPT AS change | EEAA | 0.00 | 0.000 | 0.723 | 0.83 | 0.03% | 3.12% | 0.00 | 0.000 | 0.890 | 0.94 | 0.00% | 6.58% |
| PPT AS change | IEAA | 0.00 | 0.000 | 0.381 | 0.56 | 0.14% | 3.25% | 0.00 | 0.000 | 0.310 | 0.49 | 0.20% | 6.79% |
| PPT AS change | ENP | 0.00 | 0.000 | 0.667 | 0.79 | 0.04% | 3.13% | 0.00 | 0.000 | 0.605 | 0.80 | 0.06% | 6.63% |
| PPT AS change | BLUP | 0.00 | 0.000 | 0.957 | 0.96 | 0.00% | 3.09% | 0.00 | 0.000 | 0.852 | 0.94 | 0.01% | 6.58% |
| PPT AS change | Pheno | 0.00 | 0.000 | 0.675 | 0.79 | 0.04% | 3.13% | 0.00 | 0.000 | 0.917 | 0.95 | 0.00% | 6.57% |
| PPT AS change | PCPheno | 0.00 | 0.000 | 0.599 | 0.75 | 0.07% | 3.15% | 0.00 | 0.000 | 0.810 | 0.94 | 0.02% | 6.58% |
| PPT AS change | Grim | 0.00 | 0.000 | 0.192 | 0.33 | 0.47% | 3.44% | 0.00 | 0.001 | 0.979 | 0.98 | 0.00% | 6.57% |
| PPT AS change | PCGrim | 0.00 | 0.001 | 0.121 | 0.24 | 0.77% | 3.58% | 0.00 | 0.001 | 0.756 | 0.91 | 0.05% | 6.59% |

| | | | | | | | | | | | | | |
|---|---|---|---|---|---|---|---|---|---|---|---|---|---|
| PPT AS change | DunedinPACE | 0.04 | 0.016 | 0.008 | 0.03 | 1.91% | 4.50% | 0.03 | 0.018 | 0.087 | 0.19 | 1.52% | 7.18% |
| VSM change | Horvath | 0.00 | 0.002 | 0.109 | 0.22 | 0.46% | 6.14% | 0.00 | 0.003 | 0.156 | 0.29 | 0.48% | 7.92% |
| VSM change | PCHorvath | -0.01 | 0.003 | 0.037 | 0.09 | 0.79% | 6.49% | -0.01 | 0.003 | 0.051 | 0.13 | 0.87% | 8.29% |
| VSM change | Hannum | -0.01 | 0.003 | 0.000 | 0.01 | 2.34% | 8.31% | -0.01 | 0.003 | 0.001 | 0.01 | 2.44% | 9.84% |
| VSM change | PCHannum | -0.01 | 0.003 | 0.014 | 0.04 | 1.08% | 6.81% | -0.01 | 0.003 | 0.026 | 0.08 | 1.10% | 8.53% |
| VSM change | EEAA | -0.01 | 0.002 | 0.000 | 0.00 | 2.54% | 8.66% | -0.01 | 0.002 | 0.000 | 0.01 | 2.60% | 10.11% |
| VSM change | IEAA | 0.00 | 0.003 | 0.149 | 0.27 | 0.40% | 6.04% | 0.00 | 0.003 | 0.198 | 0.34 | 0.43% | 7.85% |
| VSM change | ENP | -0.01 | 0.003 | 0.000 | 0.01 | 2.66% | 8.51% | -0.01 | 0.003 | 0.000 | 0.01 | 2.87% | 10.38% |
| VSM change | BLUP | -0.01 | 0.003 | 0.000 | 0.00 | 3.08% | 8.97% | -0.01 | 0.003 | 0.000 | 0.01 | 3.20% | 10.74% |
| VSM change | Pheno | -0.01 | 0.002 | 0.004 | 0.02 | 1.61% | 7.25% | -0.01 | 0.002 | 0.009 | 0.05 | 1.68% | 8.92% |
| VSM change | PCPheno | -0.01 | 0.002 | 0.001 | 0.01 | 2.23% | 8.01% | -0.01 | 0.003 | 0.001 | 0.01 | 2.31% | 9.65% |
| VSM change | Grim | -0.01 | 0.003 | 0.000 | 0.01 | 2.18% | 8.04% | -0.01 | 0.004 | 0.001 | 0.01 | 2.34% | 9.65% |
| VSM change | PCGrim | -0.01 | 0.005 | 0.003 | 0.01 | 1.48% | 7.41% | -0.01 | 0.005 | 0.012 | 0.05 | 1.48% | 8.82% |
| VSM change | DunedinPACE | -0.36 | 0.118 | 0.002 | 0.01 | 1.71% | 7.46% | -0.35 | 0.131 | 0.008 | 0.05 | 2.08% | 8.97% |
| VRM change | Horvath | -0.01 | 0.005 | 0.127 | 0.24 | 0.40% | 17.84% | -0.01 | 0.005 | 0.182 | 0.33 | 0.41% | 19.89% |
| VRM change | PCHorvath | -0.01 | 0.005 | 0.007 | 0.03 | 1.24% | 18.69% | -0.01 | 0.005 | 0.013 | 0.05 | 1.19% | 20.68% |
| VRM change | Hannum | -0.02 | 0.005 | 0.003 | 0.02 | 1.46% | 18.97% | -0.01 | 0.005 | 0.012 | 0.05 | 1.33% | 20.71% |
| VRM change | PCHannum | -0.01 | 0.005 | 0.006 | 0.03 | 1.27% | 18.74% | -0.01 | 0.005 | 0.014 | 0.06 | 1.19% | 20.65% |
| VRM change | EEAA | -0.01 | 0.004 | 0.002 | 0.01 | 1.59% | 19.16% | -0.01 | 0.004 | 0.007 | 0.04 | 1.47% | 20.88% |
| VRM change | IEAA | -0.01 | 0.005 | 0.174 | 0.30 | 0.34% | 17.75% | -0.01 | 0.005 | 0.223 | 0.37 | 0.36% | 19.83% |
| VRM change | ENP | -0.02 | 0.006 | 0.005 | 0.02 | 1.32% | 18.79% | -0.01 | 0.006 | 0.016 | 0.06 | 1.25% | 20.62% |
| VRM change | BLUP | -0.02 | 0.006 | 0.001 | 0.01 | 1.72% | 19.22% | -0.02 | 0.007 | 0.005 | 0.03 | 1.61% | 21.00% |
| VRM change | Pheno | 0.00 | 0.004 | 0.357 | 0.53 | 0.15% | 17.58% | 0.00 | 0.004 | 0.503 | 0.71 | 0.10% | 19.65% |
| VRM change | PCPheno | -0.02 | 0.005 | 0.001 | 0.01 | 2.01% | 19.49% | -0.02 | 0.005 | 0.001 | 0.01 | 2.02% | 21.50% |
| VRM change | Grim | -0.01 | 0.006 | 0.042 | 0.10 | 0.69% | 18.15% | -0.01 | 0.007 | 0.048 | 0.13 | 0.68% | 20.27% |
| VRM change | PCGrim | -0.02 | 0.009 | 0.036 | 0.09 | 0.70% | 18.20% | -0.02 | 0.010 | 0.032 | 0.10 | 0.78% | 20.39% |
| VRM change | DunedinPACE | -0.17 | 0.221 | 0.449 | 0.63 | 0.10% | 17.53% | -0.15 | 0.247 | 0.541 | 0.74 | 0.09% | 19.63% |

| | | | | | | | | | | | | | |
|---|---|---|---|---|---|---|---|---|---|---|---|---|---|
| Speed change | Horvath | -0.02 | 0.023 | 0.289 | 0.45 | 0.20% | 0.59% | -0.03 | 0.024 | 0.298 | 0.48 | 0.22% | 2.58% |
| Speed change | PCHorvath | -0.08 | 0.026 | 0.003 | 0.01 | 1.68% | 2.24% | -0.08 | 0.027 | 0.003 | 0.02 | 1.69% | 4.32% |
| Speed change | Hannum | -0.05 | 0.027 | 0.041 | 0.10 | 0.79% | 1.23% | -0.05 | 0.027 | 0.057 | 0.14 | 0.78% | 3.13% |
| Speed change | PCHannum | -0.06 | 0.026 | 0.014 | 0.04 | 1.09% | 1.61% | -0.07 | 0.027 | 0.015 | 0.06 | 1.14% | 3.64% |
| Speed change | EEAA | -0.05 | 0.021 | 0.029 | 0.08 | 0.87% | 1.35% | -0.05 | 0.022 | 0.035 | 0.10 | 0.93% | 3.31% |
| Speed change | IEAA | -0.02 | 0.025 | 0.530 | 0.70 | 0.07% | 0.44% | -0.01 | 0.026 | 0.604 | 0.80 | 0.06% | 2.40% |
| Speed change | ENP | -0.07 | 0.030 | 0.022 | 0.06 | 1.06% | 1.45% | -0.07 | 0.031 | 0.019 | 0.06 | 1.14% | 3.54% |
| Speed change | BLUP | -0.09 | 0.031 | 0.007 | 0.03 | 1.49% | 1.89% | -0.09 | 0.033 | 0.009 | 0.05 | 1.64% | 3.83% |
| Speed change | Pheno | -0.04 | 0.019 | 0.051 | 0.12 | 0.69% | 1.15% | -0.03 | 0.020 | 0.079 | 0.18 | 0.61% | 3.01% |
| Speed change | PCPheno | -0.06 | 0.023 | 0.008 | 0.03 | 1.28% | 1.82% | -0.06 | 0.024 | 0.008 | 0.05 | 1.39% | 3.85% |
| Speed change | Grim | -0.06 | 0.031 | 0.050 | 0.12 | 0.64% | 1.16% | -0.05 | 0.036 | 0.191 | 0.34 | 0.45% | 2.71% |
| Speed change | PCGrim | -0.15 | 0.042 | 0.000 | 0.01 | 2.12% | 2.97% | -0.15 | 0.049 | 0.002 | 0.02 | 2.13% | 4.47% |
| Speed change | DunedinPACE | -1.29 | 1.108 | 0.245 | 0.41 | 0.21% | 0.64% | -1.19 | 1.244 | 0.338 | 0.51 | 0.17% | 2.54% |

**Supplementary Table 5:Cross sectional association of epigenetic age acceleration with MRI markers**

| | | MODEL 1 | | | MODEL 2 | | | MODEL 3 | | |
|---|---|---|---|---|---|---|---|---|---|---|
| **PHENO** | **PREDICTOR** | **Beta** | ***p value*** | ***p adj*** | **Beta** | ***p value*** | ***p adj*** | **Beta** | ***p value*** | ***p adj*** |
| BPF | Horvath AA | 0.00 | 0.017 | 0.13 | 0.00 | 0.009 | 0.07 | 0.00 | 0.019 | 0.12 |
| BPF | PCHorvath AA | 0.00 | 0.226 | 0.88 | 0.00 | 0.123 | 0.53 | 0.00 | 0.228 | 0.87 |
| BPF | Hannum AA | 0.00 | 0.007 | 0.06 | 0.00 | 0.003 | 0.03 | 0.00 | 0.004 | 0.04 |
| BPF | PCHannum AA | 0.00 | 0.101 | 0.49 | 0.00 | 0.054 | 0.26 | 0.00 | 0.108 | 0.52 |
| BPF | EEAA | 0.00 | 0.005 | 0.06 | 0.00 | 0.003 | 0.03 | 0.00 | 0.003 | 0.04 |
| BPF | IEAA | 0.00 | 0.026 | 0.14 | 0.00 | 0.015 | 0.09 | 0.00 | 0.021 | 0.12 |
| BPF | ENP AA | 0.00 | 0.002 | 0.05 | 0.00 | 0.001 | 0.02 | 0.00 | 0.002 | 0.03 |
| BPF | BLUP AA | 0.00 | 0.001 | 0.04 | 0.00 | 0.000 | 0.01 | 0.00 | 0.001 | 0.02 |
| BPF | Pheno AA | 0.00 | 0.024 | 0.14 | 0.00 | 0.015 | 0.09 | 0.00 | 0.021 | 0.12 |
| BPF | PCPheno AA | 0.00 | 0.420 | 0.99 | 0.00 | 0.359 | 0.98 | 0.00 | 0.446 | 0.87 |
| BPF | Grim AA | 0.00 | 0.831 | 1.00 | 0.00 | 0.228 | 0.89 | 0.00 | 0.299 | 0.87 |
| BPF | PCGrim AA | 0.00 | 0.143 | 0.62 | 0.00 | 0.415 | 0.98 | 0.00 | 0.259 | 0.87 |
| BPF | DunedinPACE AA | 0.00 | 0.728 | 0.99 | 0.00 | 0.912 | 0.98 | 0.00 | 0.975 | 0.99 |
| WMHln | Horvath AA | 0.00 | 0.705 | 0.99 | 0.00 | 0.662 | 0.98 | 0.00 | 0.691 | 0.87 |
| WMHln | PCHorvath AA | 0.00 | 0.770 | 1.00 | 0.00 | 0.598 | 0.98 | 0.00 | 0.665 | 0.87 |
| WMHln | Hannum AA | 0.00 | 0.674 | 0.99 | 0.00 | 0.751 | 0.98 | 0.00 | 0.891 | 0.99 |
| WMHln | PCHannum AA | 0.00 | 0.898 | 1.00 | 0.00 | 0.834 | 0.98 | 0.00 | 0.970 | 0.99 |
| WMHln | EEAA | 0.00 | 0.600 | 0.99 | 0.00 | 0.728 | 0.98 | 0.00 | 0.994 | 0.99 |
| WMHln | IEAA | 0.00 | 0.731 | 0.99 | 0.00 | 0.656 | 0.98 | 0.00 | 0.569 | 0.87 |
| WMHln | ENP AA | 0.00 | 0.579 | 0.99 | 0.00 | 0.627 | 0.98 | 0.00 | 0.583 | 0.87 |
| WMHln | BLUP AA | 0.00 | 0.535 | 0.99 | 0.00 | 0.581 | 0.98 | 0.00 | 0.536 | 0.87 |
| WMHln | Pheno AA | 0.00 | 0.521 | 0.99 | 0.00 | 0.500 | 0.98 | 0.00 | 0.333 | 0.87 |
| WMHln | PCPheno AA | 0.00 | 0.923 | 1.00 | 0.00 | 0.914 | 0.98 | 0.00 | 0.603 | 0.87 |
| WMHln | Grim AA | 0.00 | 0.605 | 0.99 | 0.00 | 0.572 | 0.98 | -0.01 | 0.465 | 0.87 |
| WMHln | PCGrim AA | 0.00 | 0.804 | 1.00 | 0.00 | 0.853 | 0.98 | 0.00 | 0.902 | 0.99 |
| WMHln | DunedinPACE AA | -0.01 | 0.962 | 1.00 | -0.07 | 0.799 | 0.98 | -0.22 | 0.449 | 0.87 |
| WMHSc | Horvath AA | 0.00 | 0.713 | 0.99 | 0.00 | 0.768 | 0.98 | 0.00 | 0.611 | 0.87 |
| WMHSc | PCHorvath AA | 0.00 | 0.567 | 0.99 | 0.00 | 0.554 | 0.98 | 0.00 | 0.520 | 0.87 |

| | | | | | | | | | | |
|---|---|---|---|---|---|---|---|---|---|---|
| WMHSc | Hannum AA | 0.00 | 0.465 | 0.99 | 0.00 | 0.491 | 0.98 | 0.00 | 0.526 | 0.87 |
| WMHSc | PCHannum AA | 0.00 | 0.420 | 0.99 | 0.00 | 0.449 | 0.98 | 0.00 | 0.520 | 0.87 |
| WMHSc | EEAA | 0.00 | 0.392 | 0.99 | 0.00 | 0.437 | 0.98 | 0.00 | 0.564 | 0.87 |
| WMHSc | IEAA | 0.00 | 0.848 | 1.00 | 0.00 | 0.860 | 0.98 | 0.00 | 0.741 | 0.90 |
| WMHSc | ENP AA | 0.00 | 0.883 | 1.00 | 0.00 | 0.992 | 0.99 | 0.00 | 0.956 | 0.99 |
| WMHSc | BLUP AA | 0.00 | 0.995 | 1.00 | 0.00 | 0.854 | 0.98 | 0.00 | 0.838 | 0.99 |
| WMHSc | Pheno AA | 0.00 | 0.972 | 1.00 | 0.00 | 0.753 | 0.98 | 0.00 | 0.473 | 0.87 |
| WMHSc | PCPheno AA | 0.00 | 0.670 | 0.99 | 0.00 | 0.932 | 0.98 | 0.00 | 0.681 | 0.87 |
| WMHSc | Grim AA | 0.00 | 0.737 | 0.99 | 0.00 | 0.993 | 0.99 | 0.00 | 0.683 | 0.87 |
| WMHSc | PCGrim AA | 0.01 | 0.526 | 0.99 | 0.00 | 0.699 | 0.98 | -0.01 | 0.678 | 0.87 |
| WMHSc | DunedinPACE AA | 0.15 | 0.549 | 0.99 | 0.03 | 0.925 | 0.98 | -0.21 | 0.447 | 0.87 |

*Supplementary Table 6: Cross-sectional and longitudinal associations of epigenetic age acceleration with executive function and memory after adjusting for MRI markers*

| | | Cross-sectional: Executive Functions | | | | | | | | Longitudinal: Executive Functions | | | | | | | |
|---|---|---|---|---|---|---|---|---|---|---|---|---|---|---|---|---|---|
| | | MODEL 1 | | MODEL 2 | | MODEL WMHln | | MODEL WMHSc | | MODEL 1 | | MODEL 2 | | MODEL WMHln | | MODEL WMHSc | |
| **PHENO** | **PREDICTOR** | **Beta** | ***p value*** | **Beta** | ***p value*** | **Beta** | ***p value*** | **Beta** | ***p value*** | **Beta** | ***p value*** | **Beta** | ***p value*** | **Beta** | ***p value*** | **Beta** | ***p value*** |
| TMT | Horvath AA | -0.10 | 0.773 | 0.04 | 0.920 | -0.04 | 0.915 | -0.05 | 0.886 | 0.00 | 0.820 | 0.00 | 0.848 | 0.00 | 0.719 | 0.00 | 0.809 |
| TMT | PCHorvath AA | 0.01 | 0.974 | 0.08 | 0.852 | 0.21 | 0.623 | 0.12 | 0.778 | -0.02 | 0.056 | -0.02 | 0.078 | -0.02 | 0.169 | -0.02 | 0.138 |
| TMT | Hannum AA | -0.27 | 0.516 | -0.12 | 0.786 | 0.14 | 0.758 | 0.10 | 0.827 | -0.03 | 0.032 | -0.02 | 0.111 | -0.02 | 0.197 | -0.02 | 0.213 |
| TMT | PCHannum AA | -0.21 | 0.610 | -0.13 | 0.752 | 0.05 | 0.904 | -0.03 | 0.944 | -0.03 | 0.013 | -0.02 | 0.041 | -0.02 | 0.111 | -0.02 | 0.069 |
| TMT | EEAA | -0.31 | 0.368 | -0.21 | 0.561 | 0.01 | 0.973 | -0.02 | 0.953 | -0.02 | 0.022 | -0.02 | 0.076 | -0.01 | 0.172 | -0.01 | 0.148 |
| TMT | IEAA | 0.00 | 0.993 | 0.11 | 0.778 | 0.01 | 0.987 | 0.00 | 0.998 | 0.00 | 0.872 | 0.00 | 0.908 | 0.00 | 0.851 | 0.00 | 0.857 |
| TMT | Zhang ENP AA | 0.34 | 0.489 | 0.55 | 0.273 | 0.70 | 0.182 | 0.64 | 0.216 | -0.03 | 0.043 | -0.02 | 0.114 | -0.02 | 0.138 | -0.02 | 0.168 |
| TMT | Zhang BLUP AA | 0.02 | 0.963 | 0.12 | 0.812 | 0.28 | 0.602 | 0.24 | 0.649 | -0.04 | 0.009 | -0.03 | 0.032 | -0.03 | 0.039 | -0.03 | 0.042 |
| TMT | Pheno AA | -0.11 | 0.722 | 0.04 | 0.901 | 0.01 | 0.980 | 0.05 | 0.877 | -0.01 | 0.268 | -0.01 | 0.523 | -0.01 | 0.455 | 0.00 | 0.575 |
| TMT | PCPheno AA | -0.10 | 0.787 | -0.03 | 0.946 | 0.13 | 0.743 | 0.09 | 0.816 | -0.03 | 0.002 | -0.03 | 0.010 | -0.02 | 0.028 | -0.02 | 0.029 |
| TMT | Grim AA | -0.61 | 0.213 | -0.31 | 0.581 | -0.24 | 0.688 | -0.30 | 0.604 | -0.04 | 0.003 | -0.04 | 0.019 | -0.03 | 0.055 | -0.03 | 0.031 |
| TMT | PCGrim AA | -1.22 | 0.074 | -0.87 | 0.267 | -0.68 | 0.405 | -0.84 | 0.296 | -0.06 | 0.001 | -0.05 | 0.012 | -0.04 | 0.046 | -0.05 | 0.027 |
| TMT | DunedinPACE AA | -40.54 | 0.019 | -32.50 | 0.086 | -32.53 | 0.098 | -33.21 | 0.089 | -1.37 | 0.006 | -1.06 | 0.051 | -0.80 | 0.146 | -0.89 | 0.108 |
| WCST | Horvath AA | -0.09 | 0.087 | -0.10 | 0.060 | -0.08 | 0.111 | -0.10 | 0.060 | 0.01 | 0.141 | 0.01 | 0.100 | 0.01 | 0.151 | 0.01 | 0.095 |
| WCST | PCHorvath AA | -0.20 | 0.001 | -0.24 | 0.000 | -0.20 | 0.000 | -0.24 | 0.000 | 0.02 | 0.003 | 0.03 | 0.001 | 0.02 | 0.001 | 0.03 | 0.000 |
| WCST | Hannum AA | -0.20 | 0.001 | -0.20 | 0.002 | -0.17 | 0.003 | -0.20 | 0.002 | 0.02 | 0.002 | 0.02 | 0.004 | 0.02 | 0.006 | 0.02 | 0.004 |
| WCST | PCHannum AA | -0.22 | 0.000 | -0.25 | 0.000 | -0.21 | 0.000 | -0.27 | 0.000 | 0.02 | 0.001 | 0.03 | 0.001 | 0.02 | 0.001 | 0.03 | 0.000 |
| WCST | EEAA | -0.18 | 0.000 | -0.17 | 0.001 | -0.14 | 0.003 | -0.18 | 0.001 | 0.02 | 0.001 | 0.02 | 0.003 | 0.01 | 0.009 | 0.02 | 0.003 |
| WCST | IEAA | -0.06 | 0.261 | -0.08 | 0.172 | -0.06 | 0.214 | -0.08 | 0.176 | 0.01 | 0.251 | 0.01 | 0.161 | 0.01 | 0.184 | 0.01 | 0.163 |
| WCST | Zhang ENP AA | -0.29 | 0.000 | -0.31 | 0.000 | -0.29 | 0.000 | -0.31 | 0.000 | 0.03 | 0.001 | 0.03 | 0.000 | 0.03 | 0.000 | 0.03 | 0.000 |

| | | | | | | | | | | | | | | | | | |
|---|---|---|---|---|---|---|---|---|---|---|---|---|---|---|---|---|---|
| WCST | Zhang BLUP AA | -0.27 | 0.000 | -0.31 | 0.000 | -0.30 | 0.000 | -0.33 | 0.000 | 0.03 | 0.001 | 0.03 | 0.000 | 0.03 | 0.000 | 0.04 | 0.000 |
| WCST | Pheno AA | -0.11 | 0.010 | -0.12 | 0.011 | -0.12 | 0.005 | -0.11 | 0.014 | 0.01 | 0.016 | 0.01 | 0.017 | 0.01 | 0.006 | 0.01 | 0.022 |
| WCST | PCPheno AA | -0.25 | 0.000 | -0.25 | 0.000 | -0.25 | 0.000 | -0.27 | 0.000 | 0.02 | 0.000 | 0.02 | 0.000 | 0.02 | 0.000 | 0.02 | 0.000 |
| WCST | Grim AA | -0.14 | 0.049 | -0.17 | 0.038 | -0.17 | 0.025 | -0.18 | 0.032 | 0.02 | 0.021 | 0.02 | 0.022 | 0.02 | 0.007 | 0.02 | 0.014 |
| WCST | PCGrim AA | -0.15 | 0.135 | -0.18 | 0.109 | -0.16 | 0.117 | -0.20 | 0.096 | 0.02 | 0.065 | 0.02 | 0.103 | 0.02 | 0.109 | 0.02 | 0.107 |
| WCST | DunedinPACE AA | -5.97 | 0.016 | -6.56 | 0.016 | -6.08 | 0.015 | -6.61 | 0.018 | 0.83 | 0.008 | 0.86 | 0.013 | 0.76 | 0.014 | 0.81 | 0.018 |

| | | **Cross-Sectional: Memory** | | | | | | **Longitudinal: Memory** | | | | | |
|---|---|---|---|---|---|---|---|---|---|---|---|---|---|
| | | **MODEL 1** | | **MODEL 2** | | **MODEL BPF** | | **MODEL 1** | | **MODEL 2** | | **MODEL BPF** | |
| **PHENO** | **PREDICTOR** | **Beta** | ***p value*** | **Beta** | ***p value*** | **Beta** | ***p value*** | **Beta** | ***p value*** | **Beta** | ***p value*** | **Beta** | ***p value*** |
| VSM | Horvath AA | -0.29 | 0.208 | -0.26 | 0.264 | -0.16 | 0.532 | 0.00 | 0.109 | 0.00 | 0.156 | 0.00 | 0.256 |
| VSM | PCHorvath AA | -0.37 | 0.144 | -0.31 | 0.248 | -0.11 | 0.711 | -0.01 | 0.037 | -0.01 | 0.051 | 0.00 | 0.223 |
| VSM | Hannum AA | -0.60 | 0.027 | -0.52 | 0.063 | -0.44 | 0.154 | -0.01 | 0.000 | -0.01 | 0.001 | -0.01 | 0.008 |
| VSM | PCHannum AA | -0.43 | 0.100 | -0.34 | 0.211 | -0.21 | 0.474 | -0.01 | 0.014 | -0.01 | 0.026 | 0.00 | 0.135 |
| VSM | EEAA | -0.50 | 0.024 | -0.44 | 0.057 | -0.42 | 0.097 | -0.01 | 0.000 | -0.01 | 0.000 | -0.01 | 0.003 |
| VSM | IEAA | -0.33 | 0.171 | -0.30 | 0.221 | -0.16 | 0.571 | 0.00 | 0.149 | 0.00 | 0.198 | 0.00 | 0.360 |
| VSM | Zhang ENP AA | -0.62 | 0.048 | -0.59 | 0.064 | -0.47 | 0.185 | -0.01 | 0.000 | -0.01 | 0.000 | -0.01 | 0.002 |
| VSM | Zhang BLUP AA | -0.69 | 0.033 | -0.68 | 0.040 | -0.46 | 0.214 | -0.01 | 0.000 | -0.01 | 0.000 | -0.01 | 0.002 |
| VSM | Pheno AA | -0.23 | 0.249 | -0.13 | 0.528 | -0.01 | 0.958 | -0.01 | 0.004 | -0.01 | 0.009 | 0.00 | 0.044 |
| VSM | PCPheno AA | -0.56 | 0.020 | -0.50 | 0.043 | -0.35 | 0.189 | -0.01 | 0.001 | -0.01 | 0.001 | -0.01 | 0.016 |
| VSM | Grim AA | -0.84 | 0.007 | -0.43 | 0.237 | -0.48 | 0.220 | -0.01 | 0.000 | -0.01 | 0.001 | -0.01 | 0.007 |
| VSM | PCGrim AA | -1.16 | 0.007 | -0.61 | 0.229 | -0.55 | 0.311 | -0.01 | 0.003 | -0.01 | 0.012 | -0.01 | 0.054 |
| VSM | DunedinPACE AA | -42.22 | 0.000 | -34.23 | 0.005 | -33.28 | 0.013 | -0.36 | 0.002 | -0.35 | 0.008 | -0.33 | 0.021 |
| VRM | Horvath AA | -0.14 | 0.428 | -0.12 | 0.483 | -0.07 | 0.729 | -0.01 | 0.127 | -0.01 | 0.182 | 0.00 | 0.454 |

| | | | | | | | | | | | | | |
|---|---|---|---|---|---|---|---|---|---|---|---|---|---|
| VRM | PCHorvath AA | -0.25 | 0.197 | -0.25 | 0.231 | -0.12 | 0.595 | -0.01 | 0.007 | -0.01 | 0.013 | -0.01 | 0.102 |
| VRM | Hannum AA | -0.36 | 0.085 | -0.27 | 0.203 | -0.30 | 0.205 | -0.02 | 0.003 | -0.01 | 0.012 | -0.01 | 0.020 |
| VRM | PCHannum AA | -0.25 | 0.206 | -0.23 | 0.279 | -0.17 | 0.455 | -0.01 | 0.006 | -0.01 | 0.014 | -0.01 | 0.059 |
| VRM | EEAA | -0.34 | 0.043 | -0.27 | 0.125 | -0.32 | 0.096 | -0.01 | 0.002 | -0.01 | 0.007 | -0.01 | 0.009 |
| VRM | IEAA | -0.16 | 0.394 | -0.15 | 0.417 | -0.07 | 0.726 | -0.01 | 0.174 | -0.01 | 0.223 | 0.00 | 0.578 |
| VRM | Zhang ENP AA | -0.42 | 0.080 | -0.29 | 0.233 | -0.36 | 0.171 | -0.02 | 0.005 | -0.01 | 0.016 | -0.01 | 0.028 |
| VRM | Zhang BLUP AA | -0.38 | 0.128 | -0.31 | 0.215 | -0.21 | 0.450 | -0.02 | 0.001 | -0.02 | 0.005 | -0.02 | 0.014 |
| VRM | Pheno AA | 0.01 | 0.943 | 0.04 | 0.778 | 0.12 | 0.460 | 0.00 | 0.357 | 0.00 | 0.503 | 0.00 | 0.849 |
| VRM | PCPheno AA | -0.34 | 0.061 | -0.33 | 0.080 | -0.22 | 0.280 | -0.02 | 0.001 | | 0.001 | -0.01 | 0.010 |
| VRM | Grim AA | -0.14 | 0.557 | -0.10 | 0.732 | -0.09 | 0.765 | -0.01 | 0.042 | -0.01 | 0.048 | -0.01 | 0.075 |
| VRM | PCGrim AA | -0.47 | 0.159 | -0.56 | 0.153 | -0.34 | 0.409 | -0.02 | 0.036 | -0.02 | 0.032 | -0.02 | 0.040 |
| VRM | DunedinPACE AA | -5.33 | 0.528 | -4.08 | 0.666 | 2.71 | 0.787 | -0.17 | 0.449 | -0.15 | 0.541 | -0.09 | 0.716 |

*Supplementary Table 7: Cross-sectional and longitudinal associations of epigenetic age acceleration with cognition in presence of white blood cell counts*

| Cross-sectional Analysis | | | | | | | | | | |
|---|---|---|---|---|---|---|---|---|---|---|
| | | MODEL 1 | | | MODEL 2 | | | MODEL 3 | | |
| **PHENO** | **PREDICTOR** | **Beta** | ***p value*** | ***p adj*** | **Beta** | ***p value*** | ***p adj*** | **Beta** | ***p value*** | ***p adj*** |
| MMSE | Horvath AA | 0.01 | 0.165 | 0.32 | 0.02 | 0.099 | 0.24 | 0.02 | 0.047 | 0.19 |
| MMSE | PCHorvath AA | 0.03 | 0.026 | 0.12 | 0.03 | 0.014 | 0.08 | 0.04 | 0.003 | 0.04 |
| MMSE | Hannum AA | 0.03 | 0.009 | 0.06 | 0.04 | 0.004 | 0.04 | 0.04 | 0.001 | 0.02 |
| MMSE | PCHannum AA | 0.02 | 0.116 | 0.27 | 0.02 | 0.074 | 0.21 | 0.03 | 0.024 | 0.14 |
| MMSE | EEAA | 0.02 | 0.032 | 0.13 | 0.03 | 0.016 | 0.09 | 0.03 | 0.004 | 0.04 |
| MMSE | IEAA | 0.02 | 0.150 | 0.30 | 0.02 | 0.102 | 0.24 | 0.02 | 0.084 | 0.26 |
| MMSE | ENP AA | 0.05 | 0.001 | 0.01 | 0.06 | 0.000 | 0.00 | 0.06 | 0.000 | 0.01 |
| MMSE | BLUP AA | 0.04 | 0.003 | 0.03 | 0.04 | 0.006 | 0.05 | 0.05 | 0.004 | 0.04 |
| MMSE | Pheno AA | 0.01 | 0.147 | 0.30 | 0.01 | 0.144 | 0.30 | 0.02 | 0.096 | 0.26 |
| MMSE | PCPheno AA | 0.03 | 0.019 | 0.10 | 0.03 | 0.020 | 0.10 | 0.04 | 0.006 | 0.05 |
| MMSE | Grim AA | 0.01 | 0.612 | 0.72 | 0.02 | 0.197 | 0.38 | 0.03 | 0.147 | 0.34 |
| MMSE | PCGrim AA | 0.01 | 0.692 | 0.76 | 0.03 | 0.229 | 0.39 | 0.04 | 0.127 | 0.32 |
| MMSE | DunedinPACE AA | 0.63 | 0.219 | 0.35 | 0.71 | 0.209 | 0.39 | 0.93 | 0.115 | 0.30 |
| MDRS | Horvath AA | -0.02 | 0.351 | 0.48 | -0.01 | 0.516 | 0.64 | -0.02 | 0.395 | 0.55 |
| MDRS | PCHorvath AA | 0.04 | 0.122 | 0.28 | 0.03 | 0.258 | 0.41 | 0.03 | 0.320 | 0.50 |
| MDRS | Hannum AA | 0.02 | 0.378 | 0.51 | 0.03 | 0.297 | 0.42 | 0.03 | 0.313 | 0.50 |
| MDRS | PCHannum AA | 0.04 | 0.105 | 0.26 | 0.04 | 0.170 | 0.34 | 0.04 | 0.222 | 0.45 |
| MDRS | EEAA | 0.02 | 0.328 | 0.46 | 0.02 | 0.335 | 0.46 | 0.02 | 0.347 | 0.53 |
| MDRS | IEAA | -0.03 | 0.168 | 0.32 | -0.03 | 0.280 | 0.41 | -0.03 | 0.267 | 0.49 |
| MDRS | ENP AA | 0.03 | 0.285 | 0.42 | 0.05 | 0.125 | 0.28 | 0.05 | 0.143 | 0.34 |
| MDRS | BLUP AA | 0.03 | 0.297 | 0.43 | 0.03 | 0.291 | 0.42 | 0.03 | 0.311 | 0.50 |
| MDRS | Pheno AA | 0.01 | 0.505 | 0.63 | 0.02 | 0.281 | 0.41 | 0.02 | 0.292 | 0.50 |
| MDRS | PCPheno AA | 0.02 | 0.384 | 0.51 | 0.01 | 0.719 | 0.80 | 0.01 | 0.698 | 0.76 |
| MDRS | Grim AA | 0.03 | 0.342 | 0.48 | 0.03 | 0.447 | 0.57 | 0.03 | 0.496 | 0.65 |
| MDRS | PCGrim AA | -0.01 | 0.878 | 0.91 | -0.02 | 0.630 | 0.72 | -0.03 | 0.565 | 0.68 |
| MDRS | DunedinPACE AA | 0.30 | 0.786 | 0.82 | -0.58 | 0.626 | 0.72 | -0.87 | 0.483 | 0.64 |

| | | | | | | | | | | |
|---|---|---|---|---|---|---|---|---|---|---|
| TMT | Horvath AA | -0.10 | 0.773 | 0.82 | 0.04 | 0.920 | 0.94 | 0.06 | 0.877 | 0.90 |
| TMT | PCHorvath AA | 0.01 | 0.974 | 0.98 | 0.08 | 0.852 | 0.89 | 0.10 | 0.817 | 0.86 |
| TMT | Hannum AA | -0.27 | 0.516 | 0.63 | -0.12 | 0.786 | 0.84 | -0.04 | 0.926 | 0.93 |
| TMT | PCHannum AA | -0.21 | 0.610 | 0.72 | -0.13 | 0.752 | 0.82 | -0.12 | 0.791 | 0.85 |
| TMT | EEAA | -0.31 | 0.368 | 0.50 | -0.21 | 0.561 | 0.68 | -0.16 | 0.682 | 0.75 |
| TMT | IEAA | 0.00 | 0.993 | 0.99 | 0.11 | 0.778 | 0.84 | 0.18 | 0.642 | 0.73 |
| TMT | ENP AA | 0.34 | 0.489 | 0.62 | 0.55 | 0.273 | 0.41 | 0.64 | 0.217 | 0.45 |
| TMT | BLUP AA | 0.02 | 0.963 | 0.98 | 0.12 | 0.812 | 0.86 | 0.22 | 0.678 | 0.75 |
| TMT | Pheno AA | -0.11 | 0.722 | 0.78 | 0.04 | 0.901 | 0.92 | 0.15 | 0.660 | 0.74 |
| TMT | PCPheno AA | -0.10 | 0.787 | 0.82 | -0.03 | 0.946 | 0.95 | 0.14 | 0.743 | 0.80 |
| TMT | Grim AA | -0.61 | 0.213 | 0.35 | -0.31 | 0.581 | 0.69 | -0.29 | 0.624 | 0.72 |
| TMT | PCGrim AA | -1.22 | 0.074 | 0.22 | -0.87 | 0.267 | 0.41 | -0.78 | 0.377 | 0.54 |
| TMT | DunedinPACE AA | -40.54 | 0.019 | 0.10 | -32.50 | 0.086 | 0.23 | -33.23 | 0.092 | 0.26 |
| WCST | Horvath AA | -0.09 | 0.087 | 0.23 | -0.10 | 0.060 | 0.19 | -0.05 | 0.380 | 0.54 |
| WCST | PCHorvath AA | -0.20 | 0.001 | 0.01 | -0.24 | 0.000 | 0.00 | -0.18 | 0.007 | 0.05 |
| WCST | Hannum AA | -0.20 | 0.001 | 0.01 | -0.20 | 0.002 | 0.02 | -0.15 | 0.027 | 0.14 |
| WCST | PCHannum AA | -0.22 | 0.000 | 0.00 | -0.25 | 0.000 | 0.00 | -0.20 | 0.003 | 0.04 |
| WCST | EEAA | -0.18 | 0.000 | 0.00 | -0.17 | 0.001 | 0.01 | -0.14 | 0.014 | 0.10 |
| WCST | IEAA | -0.06 | 0.261 | 0.39 | -0.08 | 0.172 | 0.34 | -0.04 | 0.526 | 0.67 |
| WCST | ENP AA | -0.29 | 0.000 | 0.00 | -0.31 | 0.000 | 0.00 | -0.28 | 0.000 | 0.01 |
| WCST | BLUP AA | -0.27 | 0.000 | 0.00 | -0.31 | 0.000 | 0.00 | -0.27 | 0.000 | 0.01 |
| WCST | Pheno AA | -0.11 | 0.010 | 0.06 | -0.12 | 0.011 | 0.07 | -0.08 | 0.076 | 0.25 |
| WCST | PCPheno AA | -0.25 | 0.000 | 0.00 | -0.25 | 0.000 | 0.00 | -0.24 | 0.000 | 0.01 |
| WCST | Grim AA | -0.14 | 0.049 | 0.16 | -0.17 | 0.038 | 0.15 | -0.13 | 0.137 | 0.34 |
| WCST | PCGrim AA | -0.15 | 0.135 | 0.29 | -0.18 | 0.109 | 0.26 | -0.08 | 0.543 | 0.67 |
| WCST | DunedinPACE AA | -5.97 | 0.016 | 0.09 | -6.56 | 0.016 | 0.09 | -6.22 | 0.027 | 0.14 |
| PPT RLB | Horvath AA | 0.07 | 0.049 | 0.16 | 0.09 | 0.013 | 0.08 | 0.08 | 0.030 | 0.15 |
| PPT RLB | PCHorvath AA | 0.07 | 0.066 | 0.20 | 0.09 | 0.025 | 0.11 | 0.09 | 0.043 | 0.19 |
| PPT RLB | Hannum AA | 0.04 | 0.326 | 0.46 | 0.07 | 0.091 | 0.24 | 0.06 | 0.151 | 0.34 |
| PPT RLB | PCHannum AA | 0.06 | 0.164 | 0.32 | 0.08 | 0.063 | 0.19 | 0.07 | 0.095 | 0.26 |
| PPT RLB | EEAA | 0.04 | 0.268 | 0.40 | 0.06 | 0.099 | 0.24 | 0.05 | 0.166 | 0.36 |
| PPT RLB | IEAA | 0.05 | 0.139 | 0.30 | 0.07 | 0.043 | 0.15 | 0.07 | 0.052 | 0.19 |

| | | | | | | | | | | |
|---|---|---|---|---|---|---|---|---|---|---|
| PPT RLB | ENP AA | 0.13 | 0.007 | 0.06 | 0.17 | 0.001 | 0.01 | 0.16 | 0.001 | 0.02 |
| PPT RLB | BLUP AA | 0.08 | 0.116 | 0.27 | 0.11 | 0.028 | 0.11 | 0.10 | 0.041 | 0.19 |
| PPT RLB | Pheno AA | 0.05 | 0.094 | 0.24 | 0.08 | 0.010 | 0.07 | 0.08 | 0.010 | 0.07 |
| PPT RLB | PCPheno AA | 0.06 | 0.083 | 0.23 | 0.07 | 0.045 | 0.15 | 0.08 | 0.052 | 0.19 |
| PPT RLB | Grim AA | -0.06 | 0.225 | 0.35 | 0.04 | 0.469 | 0.59 | 0.05 | 0.374 | 0.54 |
| PPT RLB | PCGrim AA | -0.08 | 0.193 | 0.35 | 0.03 | 0.684 | 0.77 | 0.05 | 0.524 | 0.67 |
| PPT RLB | DunedinPACE AA | -2.22 | 0.179 | 0.33 | -0.89 | 0.627 | 0.72 | -0.47 | 0.803 | 0.85 |
| PPT Assembly | Horvath AA | 0.02 | 0.514 | 0.63 | 0.04 | 0.310 | 0.44 | 0.03 | 0.519 | 0.67 |
| PPT Assembly | PCHorvath AA | 0.02 | 0.680 | 0.76 | 0.03 | 0.451 | 0.57 | 0.03 | 0.548 | 0.67 |
| PPT Assembly | Hannum AA | 0.02 | 0.696 | 0.76 | 0.05 | 0.266 | 0.41 | 0.05 | 0.302 | 0.50 |
| PPT Assembly | PCHannum AA | 0.02 | 0.618 | 0.72 | 0.04 | 0.333 | 0.46 | 0.04 | 0.390 | 0.55 |
| PPT Assembly | EEAA | 0.02 | 0.679 | 0.76 | 0.04 | 0.333 | 0.46 | 0.04 | 0.317 | 0.50 |
| PPT Assembly | IEAA | 0.01 | 0.772 | 0.82 | 0.02 | 0.583 | 0.69 | 0.02 | 0.641 | 0.73 |
| PPT Assembly | ENP AA | 0.09 | 0.097 | 0.25 | 0.12 | 0.025 | 0.11 | 0.12 | 0.026 | 0.14 |
| PPT Assembly | BLUP AA | 0.07 | 0.220 | 0.35 | 0.09 | 0.100 | 0.24 | 0.08 | 0.140 | 0.34 |
| PPT Assembly | Pheno AA | 0.02 | 0.499 | 0.63 | 0.05 | 0.115 | 0.26 | 0.06 | 0.085 | 0.26 |
| PPT Assembly | PCPheno AA | 0.02 | 0.684 | 0.76 | 0.03 | 0.449 | 0.57 | 0.05 | 0.285 | 0.50 |
| PPT Assembly | Grim AA | -0.06 | 0.222 | 0.35 | 0.06 | 0.357 | 0.48 | 0.09 | 0.147 | 0.34 |
| PPT Assembly | PCGrim AA | -0.11 | 0.120 | 0.28 | 0.01 | 0.882 | 0.91 | 0.09 | 0.321 | 0.50 |
| PPT Assembly | DunedinPACE AA | -4.73 | 0.009 | 0.06 | -2.95 | 0.141 | 0.30 | -2.33 | 0.263 | 0.49 |
| VSM | Horvath AA | -0.29 | 0.208 | 0.35 | -0.26 | 0.264 | 0.41 | -0.30 | 0.231 | 0.47 |
| VSM | PCHorvath AA | -0.37 | 0.144 | 0.30 | -0.31 | 0.248 | 0.40 | -0.32 | 0.266 | 0.49 |
| VSM | Hannum AA | -0.60 | 0.027 | 0.12 | -0.52 | 0.063 | 0.19 | -0.51 | 0.089 | 0.26 |
| VSM | PCHannum AA | -0.43 | 0.100 | 0.25 | -0.34 | 0.211 | 0.39 | -0.33 | 0.262 | 0.49 |
| VSM | EEAA | -0.50 | 0.024 | 0.11 | -0.44 | 0.057 | 0.19 | -0.45 | 0.081 | 0.26 |
| VSM | IEAA | -0.33 | 0.171 | 0.32 | -0.30 | 0.221 | 0.39 | -0.28 | 0.270 | 0.49 |
| VSM | ENP AA | -0.62 | 0.048 | 0.16 | -0.59 | 0.064 | 0.19 | -0.59 | 0.073 | 0.24 |
| VSM | BLUP AA | -0.69 | 0.033 | 0.13 | -0.68 | 0.040 | 0.15 | -0.66 | 0.051 | 0.19 |
| VSM | Pheno AA | -0.23 | 0.249 | 0.38 | -0.13 | 0.528 | 0.64 | -0.05 | 0.828 | 0.87 |
| VSM | PCPheno AA | -0.56 | 0.020 | 0.10 | -0.50 | 0.043 | 0.15 | -0.47 | 0.094 | 0.26 |
| VSM | Grim AA | -0.84 | 0.007 | 0.06 | -0.43 | 0.237 | 0.39 | -0.34 | 0.374 | 0.54 |
| VSM | PCGrim AA | -1.16 | 0.007 | 0.06 | -0.61 | 0.229 | 0.39 | -0.30 | 0.598 | 0.71 |

| | | | | | | | | | | |
|---|---|---|---|---|---|---|---|---|---|---|
| VSM | DunedinPACE AA | -42.22 | 0.000 | 0.00 | -34.23 | 0.005 | 0.04 | -30.56 | 0.017 | 0.10 |
| VRM | Horvath AA | -0.14 | 0.428 | 0.55 | -0.12 | 0.483 | 0.60 | -0.10 | 0.613 | 0.72 |
| VRM | PCHorvath AA | -0.25 | 0.197 | 0.35 | -0.25 | 0.231 | 0.39 | -0.20 | 0.362 | 0.54 |
| VRM | Hannum AA | -0.36 | 0.085 | 0.23 | -0.27 | 0.203 | 0.39 | -0.19 | 0.406 | 0.56 |
| VRM | PCHannum AA | -0.25 | 0.206 | 0.35 | -0.23 | 0.279 | 0.41 | -0.17 | 0.446 | 0.60 |
| VRM | EEAA | -0.34 | 0.043 | 0.16 | -0.27 | 0.125 | 0.28 | -0.20 | 0.309 | 0.50 |
| VRM | IEAA | -0.16 | 0.394 | 0.51 | -0.15 | 0.417 | 0.55 | -0.10 | 0.593 | 0.71 |
| VRM | ENP AA | -0.42 | 0.080 | 0.23 | -0.29 | 0.233 | 0.39 | -0.25 | 0.323 | 0.50 |
| VRM | BLUP AA | -0.38 | 0.128 | 0.28 | -0.31 | 0.215 | 0.39 | -0.27 | 0.304 | 0.50 |
| VRM | Pheno AA | 0.01 | 0.943 | 0.97 | 0.04 | 0.778 | 0.84 | 0.13 | 0.409 | 0.56 |
| VRM | PCPheno AA | -0.34 | 0.061 | 0.19 | -0.33 | 0.080 | 0.22 | -0.22 | 0.303 | 0.50 |
| VRM | Grim AA | -0.14 | 0.557 | 0.66 | -0.10 | 0.732 | 0.81 | 0.02 | 0.946 | 0.95 |
| VRM | PCGrim AA | -0.47 | 0.159 | 0.31 | -0.56 | 0.153 | 0.31 | -0.27 | 0.535 | 0.67 |
| VRM | DunedinPACE AA | -5.33 | 0.528 | 0.64 | -4.08 | 0.666 | 0.76 | -1.67 | 0.865 | 0.90 |
| Speed | Horvath AA | 0.98 | 0.205 | 0.35 | 1.38 | 0.082 | 0.22 | 1.19 | 0.159 | 0.35 |
| Speed | PCHorvath AA | 2.10 | 0.015 | 0.09 | 2.44 | 0.007 | 0.05 | 2.36 | 0.016 | 0.10 |
| Speed | Hannum AA | 1.80 | 0.046 | 0.16 | 2.07 | 0.027 | 0.11 | 1.94 | 0.053 | 0.19 |
| Speed | PCHannum AA | 1.77 | 0.045 | 0.16 | 2.11 | 0.021 | 0.10 | 2.02 | 0.043 | 0.19 |
| Speed | EEAA | 1.43 | 0.053 | 0.17 | 1.69 | 0.028 | 0.11 | 1.73 | 0.045 | 0.19 |
| Speed | IEAA | 0.92 | 0.258 | 0.39 | 1.23 | 0.139 | 0.30 | 1.01 | 0.238 | 0.47 |
| Speed | ENP AA | 3.17 | 0.003 | 0.03 | 3.56 | 0.001 | 0.01 | 3.47 | 0.002 | 0.03 |
| Speed | BLUP AA | 3.10 | 0.004 | 0.04 | 3.55 | 0.001 | 0.02 | 3.40 | 0.003 | 0.04 |
| Speed | Pheno AA | 1.38 | 0.035 | 0.13 | 1.41 | 0.038 | 0.15 | 1.31 | 0.065 | 0.22 |
| Speed | PCPheno AA | 2.02 | 0.011 | 0.07 | 2.34 | 0.005 | 0.04 | 2.62 | 0.005 | 0.05 |
| Speed | Grim AA | 0.50 | 0.630 | 0.72 | 0.06 | 0.959 | 0.96 | -0.17 | 0.893 | 0.91 |
| Speed | PCGrim AA | 1.23 | 0.395 | 0.51 | 1.49 | 0.381 | 0.51 | 1.20 | 0.530 | 0.67 |
| Speed | DunedinPACE AA | -46.60 | 0.202 | 0.35 | -49.52 | 0.227 | 0.39 | -54.99 | 0.197 | 0.42 |
| **Longitudinal analysis** | | | | | | | | | | |
| | | **MODEL 1** | | | **MODEL 2** | | | **MODEL 3** | | |
| **PHENO** | **PREDICTOR** | **Beta** | ***p value*** | ***p adj*** | **Beta** | ***p value*** | ***p adj*** | **Beta** | ***p value*** | ***p adj*** |
| MMSE change | Horvath AA | 0.00 | 0.925 | 0.95 | 0.00 | 0.955 | 0.96 | 0.00 | 0.993 | 0.99 |

| | | | | | | | | | | |
|---|---|---|---|---|---|---|---|---|---|---|
| MMSE change | PCHorvath AA | 0.00 | 0.780 | 0.87 | 0.00 | 0.769 | 0.91 | 0.00 | 0.756 | 0.93 |
| MMSE change | Hannum AA | 0.00 | 0.596 | 0.75 | 0.00 | 0.816 | 0.94 | 0.00 | 0.777 | 0.93 |
| MMSE change | PCHannum AA | 0.00 | 0.612 | 0.75 | 0.00 | 0.653 | 0.84 | 0.00 | 0.656 | 0.87 |
| MMSE change | EEAA | 0.00 | 0.616 | 0.75 | 0.00 | 0.823 | 0.94 | 0.00 | 0.720 | 0.92 |
| MMSE change | IEAA | 0.00 | 0.817 | 0.90 | 0.00 | 0.895 | 0.94 | 0.00 | 0.823 | 0.93 |
| MMSE change | ENP AA | 0.00 | 0.581 | 0.74 | 0.00 | 0.685 | 0.85 | 0.00 | 0.679 | 0.88 |
| MMSE change | BLUP AA | 0.00 | 0.737 | 0.83 | 0.00 | 0.848 | 0.94 | 0.00 | 0.908 | 0.97 |
| MMSE change | Pheno AA | 0.00 | 0.925 | 0.95 | 0.00 | 0.889 | 0.94 | 0.00 | 0.776 | 0.93 |
| MMSE change | PCPheno AA | 0.00 | 0.482 | 0.66 | 0.00 | 0.545 | 0.74 | 0.00 | 0.756 | 0.93 |
| MMSE change | Grim AA | 0.00 | 0.157 | 0.28 | 0.00 | 0.067 | 0.16 | 0.00 | 0.076 | 0.23 |
| MMSE change | PCGrim AA | 0.00 | 0.416 | 0.59 | 0.00 | 0.222 | 0.37 | 0.00 | 0.330 | 0.55 |
| MMSE change | DunedinPACE AA | -0.01 | 0.516 | 0.69 | -0.02 | 0.363 | 0.54 | -0.02 | 0.441 | 0.68 |
| MDRS change | Horvath AA | 0.00 | 0.579 | 0.74 | 0.00 | 0.338 | 0.51 | 0.00 | 0.228 | 0.45 |
| MDRS change | PCHorvath AA | 0.00 | 0.929 | 0.95 | 0.00 | 0.768 | 0.91 | 0.00 | 0.511 | 0.74 |
| MDRS change | Hannum AA | 0.00 | 0.903 | 0.95 | 0.00 | 0.633 | 0.82 | 0.00 | 0.351 | 0.56 |
| MDRS change | PCHannum AA | 0.00 | 0.927 | 0.95 | 0.00 | 0.742 | 0.91 | 0.00 | 0.470 | 0.70 |
| MDRS change | EEAA | 0.00 | 0.998 | 1.00 | 0.00 | 0.682 | 0.85 | 0.00 | 0.279 | 0.49 |
| MDRS change | IEAA | 0.00 | 0.511 | 0.69 | 0.00 | 0.344 | 0.52 | 0.00 | 0.261 | 0.48 |
| MDRS change | ENP AA | 0.00 | 0.485 | 0.66 | 0.00 | 0.766 | 0.91 | 0.00 | 0.873 | 0.95 |
| MDRS change | BLUP AA | 0.00 | 0.412 | 0.59 | 0.00 | 0.621 | 0.82 | 0.00 | 0.637 | 0.87 |
| MDRS change | Pheno AA | 0.00 | 0.630 | 0.75 | 0.00 | 0.877 | 0.94 | 0.00 | 0.827 | 0.93 |
| MDRS change | PCPheno AA | 0.00 | 0.211 | 0.36 | 0.00 | 0.391 | 0.57 | 0.00 | 0.841 | 0.94 |
| MDRS change | Grim AA | 0.00 | 0.413 | 0.59 | 0.00 | 0.280 | 0.46 | 0.00 | 0.474 | 0.70 |
| MDRS change | PCGrim AA | 0.00 | 0.535 | 0.70 | 0.00 | 0.428 | 0.61 | 0.00 | 0.994 | 0.99 |
| MDRS change | DunedinPACE AA | -0.09 | 0.295 | 0.45 | -0.08 | 0.420 | 0.61 | -0.03 | 0.783 | 0.93 |
| TMT change | Horvath AA | 0.00 | 0.820 | 0.90 | 0.00 | 0.848 | 0.94 | 0.00 | 0.864 | 0.95 |
| TMT change | PCHorvath AA | -0.02 | 0.056 | 0.13 | -0.02 | 0.078 | 0.18 | -0.02 | 0.095 | 0.25 |
| TMT change | Hannum AA | -0.03 | 0.032 | 0.09 | -0.02 | 0.111 | 0.22 | -0.01 | 0.254 | 0.47 |
| TMT change | PCHannum AA | -0.03 | 0.013 | 0.04 | -0.02 | 0.041 | 0.12 | -0.02 | 0.068 | 0.23 |
| TMT change | EEAA | -0.02 | 0.022 | 0.06 | -0.02 | 0.076 | 0.18 | -0.01 | 0.229 | 0.45 |
| TMT change | IEAA | 0.00 | 0.872 | 0.93 | 0.00 | 0.908 | 0.95 | 0.00 | 0.714 | 0.92 |
| TMT change | ENP AA | -0.03 | 0.043 | 0.10 | -0.02 | 0.114 | 0.22 | -0.02 | 0.161 | 0.35 |

| | | | | | | | | | | |
|---|---|---|---|---|---|---|---|---|---|---|
| TMT change | BLUP AA | -0.04 | 0.009 | 0.03 | -0.03 | 0.032 | 0.10 | -0.03 | 0.066 | 0.23 |
| TMT change | Pheno AA | -0.01 | 0.268 | 0.43 | -0.01 | 0.523 | 0.73 | 0.00 | 0.903 | 0.97 |
| TMT change | PCPheno AA | -0.03 | 0.002 | 0.01 | -0.03 | 0.010 | 0.05 | -0.02 | 0.081 | 0.23 |
| TMT change | Grim AA | -0.04 | 0.003 | 0.02 | -0.04 | 0.019 | 0.06 | -0.03 | 0.063 | 0.23 |
| TMT change | PCGrim AA | -0.06 | 0.001 | 0.01 | -0.05 | 0.012 | 0.05 | -0.03 | 0.178 | 0.38 |
| TMT change | DunedinPACE AA | -1.37 | 0.006 | 0.03 | -1.06 | 0.051 | 0.13 | -0.76 | 0.185 | 0.39 |
| WCST change | Horvath AA | 0.01 | 0.141 | 0.26 | 0.01 | 0.100 | 0.21 | 0.01 | 0.364 | 0.57 |
| WCST change | PCHorvath AA | 0.02 | 0.003 | 0.02 | 0.03 | 0.001 | 0.01 | 0.02 | 0.014 | 0.09 |
| WCST change | Hannum AA | 0.02 | 0.002 | 0.01 | 0.02 | 0.004 | 0.03 | 0.01 | 0.080 | 0.23 |
| WCST change | PCHannum AA | 0.02 | 0.001 | 0.01 | 0.03 | 0.001 | 0.01 | 0.02 | 0.013 | 0.09 |
| WCST change | EEAA | 0.02 | 0.001 | 0.01 | 0.02 | 0.003 | 0.02 | 0.01 | 0.076 | 0.23 |
| WCST change | IEAA | 0.01 | 0.251 | 0.41 | 0.01 | 0.161 | 0.29 | 0.01 | 0.349 | 0.56 |
| WCST change | ENP AA | 0.03 | 0.001 | 0.01 | 0.03 | 0.000 | 0.01 | 0.03 | 0.002 | 0.04 |
| WCST change | BLUP AA | 0.03 | 0.001 | 0.01 | 0.03 | 0.000 | 0.01 | 0.03 | 0.001 | 0.03 |
| WCST change | Pheno AA | 0.01 | 0.016 | 0.05 | 0.01 | 0.017 | 0.06 | 0.01 | 0.071 | 0.23 |
| WCST change | PCPheno AA | 0.02 | 0.000 | 0.01 | 0.02 | 0.000 | 0.01 | 0.02 | 0.008 | 0.08 |
| WCST change | Grim AA | 0.02 | 0.021 | 0.06 | 0.02 | 0.022 | 0.07 | 0.02 | 0.065 | 0.23 |
| WCST change | PCGrim AA | 0.02 | 0.065 | 0.14 | 0.02 | 0.103 | 0.21 | 0.01 | 0.551 | 0.79 |
| WCST change | DunedinPACE AA | 0.83 | 0.008 | 0.03 | 0.86 | 0.013 | 0.05 | 0.94 | 0.010 | 0.08 |
| PPT RLB change | Horvath AA | 0.00 | 0.099 | 0.20 | 0.00 | 0.064 | 0.16 | 0.00 | 0.059 | 0.23 |
| PPT RLB change | PCHorvath AA | 0.00 | 0.078 | 0.16 | 0.00 | 0.073 | 0.17 | 0.00 | 0.081 | 0.23 |
| PPT RLB change | Hannum AA | 0.00 | 0.132 | 0.25 | 0.00 | 0.125 | 0.24 | 0.00 | 0.199 | 0.40 |
| PPT RLB change | PCHannum AA | 0.00 | 0.110 | 0.22 | 0.00 | 0.099 | 0.21 | 0.00 | 0.100 | 0.25 |
| PPT RLB change | EEAA | 0.00 | 0.077 | 0.16 | 0.00 | 0.093 | 0.20 | 0.00 | 0.138 | 0.33 |
| PPT RLB change | IEAA | 0.00 | 0.155 | 0.28 | 0.00 | 0.084 | 0.18 | 0.00 | 0.113 | 0.28 |
| PPT RLB change | ENP AA | 0.00 | 0.007 | 0.03 | 0.00 | 0.007 | 0.04 | 0.00 | 0.009 | 0.08 |
| PPT RLB change | BLUP AA | 0.00 | 0.306 | 0.46 | 0.00 | 0.126 | 0.24 | 0.00 | 0.153 | 0.35 |
| PPT RLB change | Pheno AA | 0.00 | 0.268 | 0.43 | 0.00 | 0.199 | 0.34 | 0.00 | 0.253 | 0.47 |
| PPT RLB change | PCPheno AA | 0.00 | 0.067 | 0.15 | 0.00 | 0.047 | 0.13 | 0.00 | 0.087 | 0.24 |
| PPT RLB change | Grim AA | 0.00 | 0.271 | 0.43 | 0.00 | 0.936 | 0.95 | 0.00 | 0.924 | 0.97 |
| PPT RLB change | PCGrim AA | 0.00 | 0.713 | 0.83 | 0.00 | 0.336 | 0.51 | 0.00 | 0.477 | 0.70 |
| PPT RLB change | DunedinPACE AA | 0.03 | 0.632 | 0.75 | 0.01 | 0.894 | 0.94 | -0.01 | 0.938 | 0.98 |

| | | | | | | | | | | |
|---|---|---|---|---|---|---|---|---|---|---|
| PPT AS change | Horvath AA | 0.00 | 0.356 | 0.53 | 0.00 | 0.286 | 0.46 | 0.00 | 0.315 | 0.53 |
| PPT AS change | PCHorvath AA | 0.00 | 0.835 | 0.90 | 0.00 | 0.811 | 0.94 | 0.00 | 0.822 | 0.93 |
| PPT AS change | Hannum AA | 0.00 | 0.727 | 0.83 | 0.00 | 0.938 | 0.95 | 0.00 | 0.791 | 0.93 |
| PPT AS change | PCHannum AA | 0.00 | 0.828 | 0.90 | 0.00 | 0.684 | 0.85 | 0.00 | 0.657 | 0.87 |
| PPT AS change | EEAA | 0.00 | 0.723 | 0.83 | 0.00 | 0.890 | 0.94 | 0.00 | 0.804 | 0.93 |
| PPT AS change | IEAA | 0.00 | 0.381 | 0.56 | 0.00 | 0.310 | 0.49 | 0.00 | 0.279 | 0.49 |
| PPT AS change | ENP AA | 0.00 | 0.667 | 0.79 | 0.00 | 0.605 | 0.80 | 0.00 | 0.666 | 0.88 |
| PPT AS change | BLUP AA | 0.00 | 0.957 | 0.96 | 0.00 | 0.852 | 0.94 | 0.00 | 0.953 | 0.98 |
| PPT AS change | Pheno AA | 0.00 | 0.675 | 0.79 | 0.00 | 0.917 | 0.95 | 0.00 | 0.961 | 0.98 |
| PPT AS change | PCPheno AA | 0.00 | 0.599 | 0.75 | 0.00 | 0.810 | 0.94 | 0.00 | 0.918 | 0.97 |
| PPT AS change | Grim AA | 0.00 | 0.192 | 0.33 | 0.00 | 0.979 | 0.98 | 0.00 | 0.610 | 0.86 |
| PPT AS change | PCGrim AA | 0.00 | 0.121 | 0.24 | 0.00 | 0.756 | 0.91 | 0.00 | 0.755 | 0.93 |
| PPT AS change | DunedinPACE AA | 0.04 | 0.008 | 0.03 | 0.03 | 0.087 | 0.19 | 0.02 | 0.306 | 0.53 |
| VSM change | Horvath AA | 0.00 | 0.109 | 0.22 | 0.00 | 0.156 | 0.29 | 0.00 | 0.156 | 0.35 |
| VSM change | PCHorvath AA | -0.01 | 0.037 | 0.09 | -0.01 | 0.051 | 0.13 | -0.01 | 0.064 | 0.23 |
| VSM change | Hannum AA | -0.01 | 0.000 | 0.01 | -0.01 | 0.001 | 0.01 | -0.01 | 0.001 | 0.03 |
| VSM change | PCHannum AA | -0.01 | 0.014 | 0.04 | -0.01 | 0.026 | 0.08 | -0.01 | 0.041 | 0.18 |
| VSM change | EEAA | -0.01 | 0.000 | 0.00 | -0.01 | 0.000 | 0.01 | -0.01 | 0.001 | 0.03 |
| VSM change | IEAA | 0.00 | 0.149 | 0.27 | 0.00 | 0.198 | 0.34 | 0.00 | 0.249 | 0.47 |
| VSM change | ENP AA | -0.01 | 0.000 | 0.01 | -0.01 | 0.000 | 0.01 | -0.01 | 0.000 | 0.01 |
| VSM change | BLUP AA | -0.01 | 0.000 | 0.00 | -0.01 | 0.000 | 0.01 | -0.01 | 0.000 | 0.01 |
| VSM change | Pheno AA | -0.01 | 0.004 | 0.02 | -0.01 | 0.009 | 0.05 | 0.00 | 0.029 | 0.16 |
| VSM change | PCPheno AA | -0.01 | 0.001 | 0.01 | -0.01 | 0.001 | 0.01 | -0.01 | 0.006 | 0.07 |
| VSM change | Grim AA | -0.01 | 0.000 | 0.01 | -0.01 | 0.001 | 0.01 | -0.01 | 0.004 | 0.06 |
| VSM change | PCGrim AA | -0.01 | 0.003 | 0.01 | -0.01 | 0.012 | 0.05 | -0.01 | 0.082 | 0.23 |
| VSM change | DunedinPACE AA | -0.36 | 0.002 | 0.01 | -0.35 | 0.008 | 0.05 | -0.30 | 0.035 | 0.18 |
| VRM change | Horvath AA | -0.01 | 0.127 | 0.24 | -0.01 | 0.182 | 0.33 | 0.00 | 0.331 | 0.55 |
| VRM change | PCHorvath AA | -0.01 | 0.007 | 0.03 | -0.01 | 0.013 | 0.05 | -0.01 | 0.044 | 0.18 |
| VRM change | Hannum AA | -0.02 | 0.003 | 0.02 | -0.01 | 0.012 | 0.05 | -0.01 | 0.040 | 0.18 |
| VRM change | PCHannum AA | -0.01 | 0.006 | 0.03 | -0.01 | 0.014 | 0.06 | -0.01 | 0.049 | 0.20 |
| VRM change | EEAA | -0.01 | 0.002 | 0.01 | -0.01 | 0.007 | 0.04 | -0.01 | 0.037 | 0.18 |
| VRM change | IEAA | -0.01 | 0.174 | 0.30 | -0.01 | 0.223 | 0.37 | 0.00 | 0.364 | 0.57 |

| | | | | | | | | | | |
|---|---|---|---|---|---|---|---|---|---|---|
| VRM change | ENP AA | -0.02 | 0.005 | 0.02 | -0.01 | 0.016 | 0.06 | -0.01 | 0.038 | 0.18 |
| VRM change | BLUP AA | -0.02 | 0.001 | 0.01 | -0.02 | 0.005 | 0.03 | -0.02 | 0.010 | 0.08 |
| VRM change | Pheno AA | 0.00 | 0.357 | 0.53 | 0.00 | 0.503 | 0.71 | 0.00 | 0.955 | 0.98 |
| VRM change | PCPheno AA | -0.02 | 0.001 | 0.01 | -0.02 | 0.001 | 0.01 | -0.01 | 0.012 | 0.09 |
| VRM change | Grim AA | -0.01 | 0.042 | 0.10 | -0.01 | 0.048 | 0.13 | -0.01 | 0.140 | 0.33 |
| VRM change | PCGrim AA | -0.02 | 0.036 | 0.09 | -0.02 | 0.032 | 0.10 | -0.01 | 0.197 | 0.40 |
| VRM change | DunedinPACE AA | -0.17 | 0.449 | 0.63 | -0.15 | 0.541 | 0.74 | -0.07 | 0.790 | 0.93 |
| Speed change | Horvath AA | -0.02 | 0.289 | 0.45 | -0.03 | 0.298 | 0.48 | -0.02 | 0.472 | 0.70 |
| Speed change | PCHorvath AA | -0.08 | 0.003 | 0.01 | -0.08 | 0.003 | 0.02 | -0.08 | 0.005 | 0.06 |
| Speed change | Hannum AA | -0.05 | 0.041 | 0.10 | -0.05 | 0.057 | 0.14 | -0.05 | 0.116 | 0.28 |
| Speed change | PCHannum AA | -0.06 | 0.014 | 0.04 | -0.07 | 0.015 | 0.06 | -0.06 | 0.038 | 0.18 |
| Speed change | EEAA | -0.05 | 0.029 | 0.08 | -0.05 | 0.035 | 0.10 | -0.04 | 0.100 | 0.25 |
| Speed change | IEAA | -0.02 | 0.530 | 0.70 | -0.01 | 0.604 | 0.80 | -0.01 | 0.616 | 0.86 |
| Speed change | ENP AA | -0.07 | 0.022 | 0.06 | -0.07 | 0.019 | 0.06 | -0.07 | 0.027 | 0.16 |
| Speed change | BLUP AA | -0.09 | 0.007 | 0.03 | -0.09 | 0.009 | 0.05 | -0.08 | 0.014 | 0.09 |
| Speed change | Pheno AA | -0.04 | 0.051 | 0.12 | -0.03 | 0.079 | 0.18 | -0.03 | 0.162 | 0.35 |
| Speed change | PCPheno AA | -0.06 | 0.008 | 0.03 | -0.06 | 0.008 | 0.05 | -0.06 | 0.020 | 0.12 |
| Speed change | Grim AA | -0.06 | 0.050 | 0.12 | -0.05 | 0.191 | 0.34 | -0.04 | 0.291 | 0.51 |
| Speed change | PCGrim AA | -0.15 | 0.000 | 0.01 | -0.15 | 0.002 | 0.02 | -0.17 | 0.003 | 0.04 |
| Speed change | DunedinPACE AA | -1.29 | 0.245 | 0.41 | -1.19 | 0.338 | 0.51 | -0.64 | 0.631 | 0.87 |

Model 1(age, sex, education), Model 2 (Model 1, HT, DM, BMI, CVD, Apoe4, cholesterol, smoking), Model 3 ( Model2, CD8T,CD4T,N K,Bcell, Mono,Neu)

*Supplementary Table 8: Cross-sectional and longitudinal variability explained in visual memory by first and third generation clocks together*

| | **Cross-sectional** | **Longitudinal** |
|---|---|---|
| BLUP AA | 0.3% | 2.6% |
| DunedinPACE AA | 0.7% | 0.8% |
| age_BL | 10.4% | 0.1% |
| sex | 0.0% | 0.1% |
| Education | 5.0% | 4.5% |
| Hypertension | 0.1% | 0.0% |
| Diabetes | 0.0% | 0.0% |
| CVD | 0.6% | 0.5% |
| BMI | 0.0% | 0.2% |
| Cholesterol | 0.0% | 0.0% |
| Smoking | 0.5% | 0.1% |
| apoe4_bi | 0.0% | 0.0% |
| **R-squared model** | 0.25 | 12% |

*Supplementary Table 9: Subgroup specific associations*

| | WCST | | | | | | | VSM | | | | | |
|---|---|---|---|---|---|---|---|---|---|---|---|---|---|
| **PREDICTOR** | **N** | **Subgroup** | **Beta** | **SE** | ***p* *value*** | ***part R2*** | ***R 2 Model*** | **N** | **Beta** | **SE** | ***p* *value*** | ***part R2*** | ***R 2 Model*** |
| BLUP AA | 364 | Young | -0.33 | 0.100 | 0.001 | 3.10% | 9.07% | 357 | -0.86 | 0.478 | 0.073 | 0.71% | 17.00% |
| BLUP AA | 350 | Old | -0.28 | 0.110 | 0.010 | 1.53% | 4.05% | 337 | -0.37 | 0.455 | 0.413 | 0.24% | 16.26% |
| BLUP AA | 301 | Male | -0.32 | 0.112 | 0.004 | 2.49% | 6.41% | 297 | -0.45 | 0.465 | 0.337 | 0.22% | 29.95% |
| BLUP AA | 413 | Female | -0.32 | 0.100 | 0.002 | 2.20% | 5.14% | 397 | -0.95 | 0.474 | 0.045 | 0.84% | 21.42% |
| BLUP AA | 486 | Low_Ed | -0.40 | 0.095 | 0.000 | 2.97% | 7.87% | 466 | -0.62 | 0.379 | 0.103 | 0.47% | 21.19% |
| BLUP AA | 228 | High_Ed | -0.15 | 0.109 | 0.168 | 0.99% | 7.06% | 228 | -1.12 | 0.661 | 0.091 | 1.05% | 23.96% |
| BLUP AA | 217 | Normotensive | -0.28 | 0.111 | 0.012 | 3.06% | 8.86% | 213 | -1.22 | 0.572 | 0.034 | 1.86% | 23.45% |
| BLUP AA | 497 | Hypertensive | -0.30 | 0.096 | 0.002 | 2.07% | 4.92% | 481 | -0.40 | 0.408 | 0.322 | 0.21% | 22.14% |
| BLUP AA | 253 | Normal weight | -0.33 | 0.116 | 0.005 | 3.55% | 11.93% | 247 | -2.10 | 0.585 | 0.000 | 4.50% | 24.44% |
| BLUP AA | 461 | Overweight | -0.29 | 0.095 | 0.002 | 1.83% | 4.51% | 447 | 0.02 | 0.400 | 0.962 | 0.00% | 27.35% |
| BLUP AA | 424 | No_CVD | -0.28 | 0.074 | 0.000 | 2.99% | 6.02% | 415 | -0.61 | 0.400 | 0.129 | 0.39% | 26.32% |
| BLUP AA | 290 | CVD | -0.37 | 0.155 | 0.018 | 2.12% | 5.26% | 279 | -0.88 | 0.586 | 0.133 | 0.88% | 19.80% |
| BLUP AA | 435 | Never | -0.31 | 0.104 | 0.003 | 2.09% | 6.60% | 421 | -1.19 | 0.441 | 0.007 | 1.34% | 27.97% |
| BLUP AA | 201 | Former | -0.32 | 0.116 | 0.006 | 3.67% | 7.14% | 196 | -0.24 | 0.622 | 0.702 | 0.05% | 21.75% |
| BLUP AA | 78 | Current | -0.28 | 0.219 | 0.197 | 2.15% | 10.88% | 77 | 0.09 | 0.862 | 0.915 | 0.02% | 31.47% |
| BLUP AA | 636 | Nondiabetic | -0.37 | 0.082 | 0.000 | 3.19% | 5.64% | 622 | -0.97 | 0.358 | 0.007 | 0.91% | 24.50% |
| BLUP AA | 78 | Diabetic | 0.00 | 0.152 | 0.984 | 0.00% | 8.76% | 72 | 0.32 | 0.918 | 0.732 | 0.27% | 28.25% |
| BLUP AA | 507 | WMHSC_0_1 | -0.38 | 0.088 | 0.000 | 3.30% | 7.34% | 495 | -0.53 | 0.382 | 0.166 | 0.31% | 21.93% |
| BLUP AA | 162 | WMHSC_2_3 | -0.07 | 0.143 | 0.642 | 0.22% | 5.32% | 154 | -1.30 | 0.780 | 0.098 | 1.60% | 27.79% |
| PCPheno AA | 364 | Young | -0.35 | 0.083 | 0.000 | 5.67% | 10.64% | 357 | -0.89 | 0.401 | 0.027 | 1.22% | 17.40% |
| PCPheno AA | 350 | Old | -0.19 | 0.080 | 0.019 | 1.32% | 3.75% | 337 | -0.16 | 0.302 | 0.596 | 0.09% | 16.16% |
| PCPheno AA | 301 | Male | -0.31 | 0.087 | 0.000 | 3.97% | 7.78% | 297 | -0.33 | 0.334 | 0.325 | 0.31% | 29.96% |
| PCPheno AA | 413 | Female | -0.24 | 0.079 | 0.003 | 2.17% | 4.91% | 397 | -0.69 | 0.375 | 0.065 | 0.81% | 21.29% |

| | | | | | | | | | | | | | |
|---|---|---|---|---|---|---|---|---|---|---|---|---|---|
| PCPheno AA | 486 | Low_Ed | -0.29 | 0.073 | 0.000 | 2.83% | 7.49% | 466 | -0.53 | 0.295 | 0.071 | 0.74% | 21.30% |
| PCPheno AA | 228 | High_Ed | -0.21 | 0.088 | 0.016 | 3.51% | 8.72% | 228 | -0.55 | 0.465 | 0.236 | 0.51% | 23.45% |
| PCPheno AA | 217 | Normotensive | -0.21 | 0.087 | 0.016 | 3.74% | 8.62% | 213 | -1.17 | 0.450 | 0.010 | 2.98% | 24.28% |
| PCPheno AA | 497 | Hypertensive | -0.26 | 0.075 | 0.000 | 2.68% | 5.37% | 481 | -0.21 | 0.299 | 0.491 | 0.12% | 22.05% |
| PCPheno AA | 253 | Normal weight | -0.29 | 0.097 | 0.003 | 5.16% | 12.33% | 247 | -1.01 | 0.504 | 0.046 | 1.74% | 21.62% |
| PCPheno AA | 461 | Overweight | -0.23 | 0.072 | 0.001 | 2.21% | 4.72% | 447 | -0.29 | 0.285 | 0.308 | 0.26% | 27.53% |
| PCPheno AA | 424 | No_CVD | -0.23 | 0.059 | 0.000 | 3.14% | 6.07% | 415 | -0.71 | 0.299 | 0.017 | 1.24% | 26.93% |
| PCPheno AA | 290 | CVD | -0.31 | 0.114 | 0.008 | 2.95% | 5.77% | 279 | -0.09 | 0.439 | 0.832 | 0.02% | 19.13% |
| PCPheno AA | 435 | Never | -0.29 | 0.081 | 0.000 | 3.64% | 7.43% | 421 | -0.92 | 0.350 | 0.009 | 1.38% | 27.92% |
| PCPheno AA | 201 | Former | -0.17 | 0.087 | 0.047 | 2.04% | 5.31% | 196 | -0.08 | 0.408 | 0.838 | 0.02% | 21.70% |
| PCPheno AA | 78 | Current | -0.28 | 0.169 | 0.099 | 2.48% | 12.29% | 77 | -0.05 | 0.668 | 0.937 | 0.01% | 31.47% |
| PCPheno AA | 636 | Nondiabetic | -0.27 | 0.065 | 0.000 | 3.19% | 5.13% | 622 | -0.60 | 0.269 | 0.027 | 0.73% | 24.21% |
| PCPheno AA | 78 | Diabetic | -0.23 | 0.106 | 0.031 | 4.40% | 15.01% | 72 | -0.41 | 0.687 | 0.553 | 0.27% | 28.54% |
| PCPheno AA | 507 | WMHSC_0_1 | -0.27 | 0.073 | 0.000 | 3.08% | 6.56% | 495 | -0.38 | 0.314 | 0.232 | 0.35% | 21.85% |
| PCPheno AA | 162 | WMHSC_2_3 | -0.26 | 0.094 | 0.006 | 4.78% | 9.90% | 154 | -0.51 | 0.436 | 0.243 | 0.81% | 27.08% |
| PCGrim AA | 364 | Young | -0.32 | 0.162 | 0.050 | 1.03% | 7.23% | 357 | -1.09 | 0.787 | 0.167 | 0.79% | 16.68% |
| PCGrim AA | 350 | Old | -0.11 | 0.167 | 0.501 | 0.11% | 2.29% | 337 | -0.27 | 0.664 | 0.687 | 0.07% | 16.13% |
| PCGrim AA | 301 | Male | -0.15 | 0.172 | 0.368 | 0.22% | 4.01% | 297 | -0.37 | 0.700 | 0.599 | 0.12% | 29.79% |
| PCGrim AA | 413 | Female | -0.21 | 0.158 | 0.175 | 0.42% | 3.24% | 397 | -0.88 | 0.751 | 0.242 | 0.51% | 20.87% |
| PCGrim AA | 486 | Low_Ed | -0.17 | 0.147 | 0.246 | 0.22% | 4.76% | 466 | -0.38 | 0.598 | 0.525 | 0.14% | 20.80% |
| PCGrim AA | 228 | High_Ed | -0.34 | 0.173 | 0.054 | 4.28% | 7.84% | 228 | -1.27 | 0.991 | 0.202 | 1.12% | 23.53% |
| PCGrim AA | 217 | Normotensive | -0.35 | 0.169 | 0.041 | 3.46% | 7.91% | 213 | -0.46 | 0.903 | 0.610 | 0.16% | 21.81% |
| PCGrim AA | 497 | Hypertensive | -0.07 | 0.151 | 0.629 | 0.04% | 2.99% | 481 | -0.72 | 0.632 | 0.257 | 0.68% | 22.19% |
| PCGrim AA | 253 | Normal weight | -0.42 | 0.190 | 0.028 | 2.43% | 10.82% | 247 | -0.74 | 1.010 | 0.465 | 0.25% | 20.46% |
| PCGrim AA | 461 | Overweight | -0.08 | 0.144 | 0.595 | 0.04% | 2.59% | 447 | -0.46 | 0.594 | 0.442 | 0.31% | 27.45% |
| PCGrim AA | 424 | No_CVD | -0.20 | 0.119 | 0.093 | 0.74% | 3.30% | 415 | -1.28 | 0.631 | 0.043 | 1.35% | 26.64% |
| PCGrim AA | 290 | CVD | -0.21 | 0.232 | 0.362 | 0.20% | 3.61% | 279 | 0.34 | 0.882 | 0.699 | 0.04% | 19.17% |

| | | | | | | | | | | | | |
|---|---|---|---|---|---|---|---|---|---|---|---|---|
| PCGrim AA | 435 | Never | -0.27 | 0.182 | 0.138 | 0.49% | 5.17% | 421 | -0.92 | 0.791 | 0.246 | 0.32% | 26.93% |
| PCGrim AA | 201 | Former | -0.17 | 0.161 | 0.300 | 0.64% | 3.86% | 196 | -0.47 | 0.828 | 0.569 | 0.17% | 21.82% |
| PCGrim AA | 78 | Current | -0.28 | 0.280 | 0.329 | 0.66% | 9.93% | 77 | -0.76 | 1.093 | 0.491 | 0.52% | 31.96% |
| PCGrim AA | 636 | Nondiabetic | -0.19 | 0.129 | 0.136 | 0.44% | 2.88% | 622 | -0.72 | 0.555 | 0.198 | 0.48% | 23.80% |
| PCGrim AA | 78 | Diabetic | -0.19 | 0.232 | 0.419 | 0.65% | 9.66% | 72 | -0.67 | 1.459 | 0.649 | 0.17% | 28.36% |
| PCGrim AA | 507 | WMHSC_0_1 | -0.21 | 0.144 | 0.144 | 0.32% | 4.28% | 495 | -0.16 | 0.626 | 0.801 | 0.02% | 21.63% |
| PCGrim AA | 162 | WMHSC_2_3 | -0.21 | 0.186 | 0.255 | 1.54% | 6.01% | 154 | -1.32 | 0.956 | 0.171 | 1.59% | 27.35% |
| DunedinPACE AA | 364 | Young | -10.41 | 4.129 | 0.012 | 2.18% | 7.87% | 357 | -47.35 | 19.863 | 0.018 | 2.16% | 17.58% |
| DunedinPACE AA | 350 | Old | -3.46 | 3.681 | 0.348 | 0.27% | 2.42% | 337 | -21.51 | 15.238 | 0.159 | 0.88% | 16.60% |
| DunedinPACE AA | 301 | Male | -9.79 | 4.152 | 0.019 | 1.82% | 5.56% | 297 | -22.64 | 17.578 | 0.199 | 0.77% | 30.13% |
| DunedinPACE AA | 413 | Female | -4.02 | 3.655 | 0.272 | 0.34% | 3.08% | 397 | -45.24 | 17.241 | 0.009 | 1.80% | 21.99% |
| DunedinPACE AA | 217 | Normotensive | -7.56 | 4.213 | 0.074 | 2.74% | 7.46% | 213 | -48.13 | 22.307 | 0.032 | 2.24% | 23.48% |
| DunedinPACE AA | 497 | Hypertensive | -5.47 | 3.487 | 0.117 | 0.48% | 3.43% | 481 | -30.04 | 14.920 | 0.045 | 1.28% | 22.64% |
| DunedinPACE AA | 253 | Normal weight | -10.16 | 4.354 | 0.020 | 3.18% | 11.01% | 247 | -24.10 | 22.410 | 0.283 | 0.53% | 20.67% |
| DunedinPACE AA | 461 | Overweight | -5.04 | 3.521 | 0.153 | 0.36% | 2.98% | 447 | -38.14 | 14.960 | 0.011 | 1.78% | 28.43% |
| DunedinPACE AA | 486 | Low_Ed | -3.95 | 3.555 | 0.267 | 0.23% | 4.74% | 466 | -32.18 | 14.465 | 0.027 | 1.27% | 21.58% |
| DunedinPACE AA | 228 | High_Ed | -13.44 | 3.873 | 0.001 | 7.97% | 11.21% | 228 | -40.54 | 23.593 | 0.087 | 2.21% | 23.99% |
| DunedinPACE AA | 424 | No_CVD | -9.29 | 2.929 | 0.002 | 2.34% | 4.96% | 415 | -52.24 | 15.808 | 0.001 | 2.75% | 27.85% |
| DunedinPACE AA | 290 | CVD | -4.85 | 5.160 | 0.348 | 0.23% | 3.63% | 279 | -12.12 | 19.869 | 0.542 | 0.21% | 19.23% |
| DunedinPACE AA | 435 | Never | -4.74 | 4.019 | 0.239 | 0.39% | 4.99% | 421 | -36.48 | 17.433 | 0.037 | 0.90% | 27.47% |
| DunedinPACE AA | 201 | Former | -10.21 | 3.814 | 0.008 | 3.43% | 6.83% | 196 | -24.97 | 20.777 | 0.231 | 0.88% | 22.29% |
| DunedinPACE AA | 78 | Current | -7.37 | 8.162 | 0.369 | 0.77% | 9.73% | 77 | -63.64 | 30.784 | 0.043 | 4.78% | 35.63% |
| DunedinPACE AA | 636 | Nondiabetic | -5.68 | 2.993 | 0.058 | 0.85% | 3.09% | 622 | -36.65 | 13.024 | 0.005 | 1.63% | 24.57% |
| DunedinPACE AA | 78 | Diabetic | -12.27 | 5.960 | 0.043 | 4.43% | 14.27% | 72 | -20.69 | 40.093 | 0.608 | 0.18% | 28.43% |
| DunedinPACE AA | 507 | WMHSC_0_1 | -7.29 | 3.537 | 0.040 | 0.81% | 4.69% | 495 | -29.99 | 15.301 | 0.051 | 1.23% | 22.24% |
| DunedinPACE AA | 162 | WMHSC_2_3 | -6.90 | 4.015 | 0.088 | 2.13% | 7.02% | 154 | -47.29 | 22.074 | 0.034 | 3.40% | 28.69% |
| | **TMT Change** | | | | | | | **VSM Change** | | | | | |

| | N | Subgroup | Beta | SE | *p value* | *part R2* | *R 2 Model* | N | Beta | SE | *p value* | *part R2* | *R 2 Model* |
|---|---|---|---|---|---|---|---|---|---|---|---|---|---|
| BLUP AA | 276 | Young | -0.04 | 0.016 | 0.012 | 1.97% | 18.13% | 275 | -0.02 | 0.005 | 0.002 | 3.14% | 13.50% |
| BLUP AA | 187 | Old | -0.03 | 0.027 | 0.283 | 0.69% | 13.30% | 182 | -0.01 | 0.005 | 0.028 | 2.63% | 9.69% |
| BLUP AA | 209 | Male | -0.02 | 0.019 | 0.251 | 0.70% | 10.55% | 209 | -0.01 | 0.005 | 0.046 | 1.91% | 14.78% |
| BLUP AA | 254 | Female | -0.04 | 0.022 | 0.083 | 1.18% | 18.03% | 248 | -0.02 | 0.005 | 0.000 | 5.39% | 10.09% |
| BLUP AA | 302 | Low_Ed | -0.05 | 0.018 | 0.006 | 2.98% | 13.07% | 296 | -0.02 | 0.004 | 0.000 | 5.08% | 11.74% |
| BLUP AA | 161 | High_Ed | -0.02 | 0.023 | 0.472 | 0.26% | 15.46% | 161 | -0.01 | 0.007 | 0.091 | 1.60% | 12.73% |
| BLUP AA | 162 | Normotensive | -0.03 | 0.024 | 0.289 | 0.58% | 15.01% | 162 | -0.02 | 0.006 | 0.012 | 3.92% | 10.10% |
| BLUP AA | 301 | Hypertensive | -0.04 | 0.018 | 0.057 | 1.31% | 13.48% | 295 | -0.01 | 0.004 | 0.003 | 2.80% | 12.23% |
| BLUP AA | 163 | Normal weight | -0.04 | 0.025 | 0.109 | 1.59% | 18.78% | 160 | -0.02 | 0.006 | 0.000 | 7.14% | 21.49% |
| BLUP AA | 300 | Overweight | -0.03 | 0.018 | 0.128 | 0.70% | 12.07% | 297 | -0.01 | 0.004 | 0.042 | 1.41% | 9.89% |
| BLUP AA | 300 | No_CVD | -0.01 | 0.017 | 0.402 | 0.21% | 10.35% | 298 | -0.01 | 0.004 | 0.020 | 1.81% | 9.92% |
| BLUP AA | 163 | CVD | -0.07 | 0.027 | 0.013 | 3.74% | 16.83% | 159 | -0.02 | 0.006 | 0.000 | 7.10% | 17.01% |
| BLUP AA | 282 | Never | -0.03 | 0.018 | 0.111 | 0.96% | 13.74% | 277 | -0.02 | 0.005 | 0.000 | 5.81% | 15.79% |
| BLUP AA | 133 | Former | -0.06 | 0.030 | 0.033 | 3.38% | 15.20% | 132 | -0.01 | 0.007 | 0.307 | 0.62% | 5.45% |
| BLUP AA | 48 | Current | 0.05 | 0.051 | 0.321 | 1.44% | 36.98% | 48 | -0.01 | 0.012 | 0.365 | 1.99% | 22.00% |
| BLUP AA | 421 | Nondiabetic | -0.05 | 0.015 | 0.002 | 2.15% | 9.68% | 418 | -0.02 | 0.004 | 0.000 | 3.41% | 12.59% |
| BLUP AA | 42 | Diabetic | 0.08 | 0.063 | 0.239 | 4.15% | 17.34% | 39 | -0.02 | 0.013 | 0.192 | 2.92% | 10.17% |
| BLUP AA | 352 | WMHSC_0_1 | -0.03 | 0.016 | 0.044 | 1.24% | 12.20% | 347 | -0.01 | 0.004 | 0.001 | 2.94% | 9.83% |
| BLUP AA | 97 | WMHSC_2_3 | -0.03 | 0.038 | 0.363 | 0.64% | 23.74% | 96 | -0.02 | 0.008 | 0.035 | 4.18% | 23.01% |
| PCPheno AA | 276 | Young | -0.03 | 0.013 | 0.034 | 1.46% | 17.57% | 275 | -0.01 | 0.004 | 0.000 | 4.59% | 14.44% |
| PCPheno AA | 187 | Old | -0.03 | 0.017 | 0.072 | 2.30% | 14.33% | 182 | 0.00 | 0.003 | 0.433 | 0.34% | 7.40% |
| PCPheno AA | 209 | Male | -0.02 | 0.014 | 0.202 | 0.91% | 10.69% | 209 | 0.00 | 0.003 | 0.256 | 0.86% | 13.61% |
| PCPheno AA | 254 | Female | -0.04 | 0.016 | 0.021 | 2.38% | 18.82% | 248 | -0.01 | 0.004 | 0.001 | 4.96% | 9.10% |
| PCPheno AA | 302 | Low_Ed | -0.04 | 0.014 | 0.007 | 3.04% | 12.96% | 296 | -0.01 | 0.003 | 0.004 | 3.64% | 9.59% |
| PCPheno AA | 161 | High_Ed | -0.02 | 0.015 | 0.302 | 0.80% | 15.78% | 161 | -0.01 | 0.004 | 0.223 | 0.99% | 11.91% |

| | | | | | | | | | | | | | |
|---|---|---|---|---|---|---|---|---|---|---|---|---|---|
| PCPheno AA | 162 | Normotensive | -0.04 | 0.019 | 0.057 | 1.94% | 16.43% | 162 | -0.02 | 0.005 | 0.000 | 8.30% | 14.10% |
| PCPheno AA | 301 | Hypertensive | -0.02 | 0.013 | 0.068 | 1.39% | 13.39% | 295 | 0.00 | 0.003 | 0.178 | 0.74% | 10.05% |
| PCPheno AA | 163 | Normal weight | -0.04 | 0.022 | 0.108 | 1.81% | 18.79% | 160 | -0.01 | 0.005 | 0.030 | 3.15% | 16.66% |
| PCPheno AA | 300 | Overweight | -0.02 | 0.012 | 0.039 | 1.49% | 12.66% | 297 | -0.01 | 0.003 | 0.027 | 1.73% | 10.13% |
| PCPheno AA | 300 | No_CVD | -0.02 | 0.012 | 0.211 | 0.61% | 10.62% | 298 | -0.01 | 0.003 | 0.004 | 2.85% | 10.85% |
| PCPheno AA | 163 | CVD | -0.05 | 0.020 | 0.010 | 4.18% | 17.10% | 159 | -0.01 | 0.004 | 0.137 | 1.36% | 10.89% |
| PCPheno AA | 282 | Never | -0.03 | 0.013 | 0.018 | 2.55% | 14.72% | 277 | -0.01 | 0.003 | 0.000 | 4.66% | 14.33% |
| PCPheno AA | 133 | Former | -0.03 | 0.019 | 0.135 | 1.62% | 13.58% | 132 | 0.00 | 0.004 | 0.938 | 0.01% | 4.63% |
| PCPheno AA | 48 | Current | 0.03 | 0.038 | 0.406 | 1.02% | 36.46% | 48 | -0.01 | 0.009 | 0.225 | 5.04% | 23.38% |
| PCPheno AA | 421 | Nondiabetic | -0.03 | 0.010 | 0.001 | 2.65% | 9.74% | 418 | -0.01 | 0.003 | 0.001 | 2.61% | 11.52% |
| PCPheno AA | 42 | Diabetic | 0.02 | 0.051 | 0.658 | 1.34% | 13.93% | 39 | -0.01 | 0.011 | 0.562 | 0.55% | 5.42% |
| PCPheno AA | 352 | WMHSC_0_1 | -0.03 | 0.013 | 0.047 | 1.39% | 12.18% | 347 | -0.01 | 0.003 | 0.002 | 3.17% | 9.52% |
| PCPheno AA | 97 | WMHSC_2_3 | -0.03 | 0.020 | 0.110 | 2.08% | 25.30% | 96 | 0.00 | 0.004 | 0.369 | 0.82% | 19.53% |
| PCGrim AA | 276 | Young | -0.04 | 0.025 | 0.115 | 0.79% | 16.94% | 275 | -0.02 | 0.007 | 0.011 | 2.24% | 12.34% |
| PCGrim AA | 187 | Old | -0.08 | 0.040 | 0.037 | 3.00% | 14.88% | 182 | -0.01 | 0.008 | 0.288 | 0.72% | 7.68% |
| PCGrim AA | 209 | Male | -0.03 | 0.030 | 0.403 | 0.29% | 10.27% | 209 | -0.01 | 0.008 | 0.381 | 0.81% | 13.38% |
| PCGrim AA | 254 | Female | -0.09 | 0.032 | 0.004 | 3.17% | 19.75% | 248 | -0.02 | 0.007 | 0.005 | 2.50% | 7.54% |
| PCGrim AA | 302 | Low_Ed | -0.06 | 0.028 | 0.040 | 1.66% | 12.04% | 296 | -0.01 | 0.006 | 0.039 | 1.56% | 8.25% |
| PCGrim AA | 161 | High_Ed | -0.05 | 0.032 | 0.114 | 2.18% | 16.59% | 161 | -0.01 | 0.009 | 0.182 | 1.44% | 12.09% |
| PCGrim AA | 162 | Normotensive | -0.08 | 0.037 | 0.027 | 2.42% | 17.14% | 162 | -0.02 | 0.009 | 0.055 | 2.25% | 8.54% |
| PCGrim AA | 301 | Hypertensive | -0.04 | 0.027 | 0.174 | 0.72% | 12.95% | 295 | -0.01 | 0.006 | 0.122 | 1.09% | 10.23% |
| PCGrim AA | 163 | Normal weight | -0.08 | 0.045 | 0.073 | 2.69% | 19.13% | 160 | -0.02 | 0.010 | 0.024 | 4.15% | 16.89% |
| PCGrim AA | 300 | Overweight | -0.04 | 0.025 | 0.090 | 0.90% | 12.24% | 297 | -0.01 | 0.006 | 0.174 | 0.53% | 9.16% |
| PCGrim AA | 300 | No_CVD | -0.05 | 0.025 | 0.051 | 1.40% | 11.31% | 298 | -0.02 | 0.006 | 0.008 | 2.50% | 10.41% |
| PCGrim AA | 163 | CVD | -0.07 | 0.043 | 0.117 | 1.21% | 14.78% | 159 | -0.01 | 0.009 | 0.433 | 0.44% | 9.92% |
| PCGrim AA | 282 | Never | -0.06 | 0.030 | 0.038 | 1.83% | 14.31% | 277 | -0.01 | 0.008 | 0.071 | 1.12% | 10.94% |
| PCGrim AA | 133 | Former | -0.04 | 0.042 | 0.384 | 0.76% | 12.52% | 132 | -0.01 | 0.009 | 0.236 | 1.41% | 5.73% |

| | | | | | | | | | | | | | |
|---|---|---|---|---|---|---|---|---|---|---|---|---|---|
| PCGrim AA | 48 | Current | -0.09 | 0.055 | 0.120 | 4.31% | 39.40% | 48 | -0.02 | 0.013 | 0.147 | 5.48% | 24.70% |
| PCGrim AA | 421 | Nondiabetic | -0.06 | 0.022 | 0.003 | 1.90% | 9.36% | 418 | -0.02 | 0.006 | 0.006 | 1.92% | 10.70% |
| PCGrim AA | 42 | Diabetic | -0.01 | 0.104 | 0.955 | 0.01% | 13.37% | 39 | -0.01 | 0.020 | 0.741 | 0.23% | 4.61% |
| PCGrim AA | 352 | WMHSC_0_1 | -0.04 | 0.025 | 0.077 | 0.97% | 11.96% | 347 | -0.01 | 0.006 | 0.046 | 1.56% | 7.99% |
| PCGrim AA | 97 | WMHSC_2_3 | -0.09 | 0.049 | 0.073 | 2.71% | 25.90% | 96 | -0.01 | 0.010 | 0.387 | 0.88% | 19.47% |
| DunedinPACE AA | 276 | Young | -1.17 | 0.653 | 0.075 | 1.03% | 17.16% | 275 | -0.60 | 0.192 | 0.002 | 4.09% | 13.35% |
| DunedinPACE AA | 187 | Old | -1.03 | 0.942 | 0.277 | 1.34% | 13.32% | 182 | -0.04 | 0.177 | 0.833 | 0.04% | 7.08% |
| DunedinPACE AA | 209 | Male | -0.85 | 0.768 | 0.271 | 0.53% | 10.50% | 209 | -0.11 | 0.192 | 0.561 | 0.27% | 13.19% |
| DunedinPACE AA | 254 | Female | -1.35 | 0.776 | 0.083 | 1.55% | 18.03% | 248 | -0.64 | 0.183 | 0.001 | 4.78% | 9.06% |
| DunedinPACE AA | 162 | Normotensive | -1.22 | 0.940 | 0.195 | 0.87% | 15.33% | 162 | -0.57 | 0.235 | 0.016 | 4.27% | 9.84% |
| DunedinPACE AA | 301 | Hypertensive | -0.97 | 0.680 | 0.155 | 0.96% | 13.00% | 295 | -0.23 | 0.163 | 0.165 | 1.16% | 10.09% |
| DunedinPACE AA | 163 | Normal weight | -0.52 | 1.058 | 0.622 | 0.26% | 17.51% | 160 | -0.30 | 0.242 | 0.217 | 1.61% | 14.83% |
| DunedinPACE AA | 300 | Overweight | -1.20 | 0.645 | 0.065 | 1.42% | 12.41% | 297 | -0.35 | 0.161 | 0.030 | 1.78% | 10.07% |
| DunedinPACE AA | 302 | Low_Ed | -0.95 | 0.721 | 0.187 | 0.71% | 11.28% | 296 | -0.31 | 0.160 | 0.058 | 1.58% | 8.02% |
| DunedinPACE AA | 161 | High_Ed | -1.61 | 0.821 | 0.052 | 3.57% | 17.31% | 161 | -0.39 | 0.242 | 0.108 | 2.77% | 12.57% |
| DunedinPACE AA | 300 | No_CVD | -1.41 | 0.678 | 0.038 | 1.94% | 11.47% | 298 | -0.59 | 0.172 | 0.001 | 4.33% | 11.84% |
| DunedinPACE AA | 163 | CVD | -0.62 | 0.935 | 0.509 | 0.18% | 13.63% | 159 | -0.03 | 0.204 | 0.867 | 0.02% | 9.56% |
| DunedinPACE AA | 282 | Never | -0.99 | 0.713 | 0.167 | 1.38% | 13.54% | 277 | -0.40 | 0.187 | 0.031 | 2.01% | 11.40% |
| DunedinPACE AA | 133 | Former | -0.76 | 1.058 | 0.475 | 0.52% | 12.34% | 132 | -0.22 | 0.228 | 0.345 | 0.94% | 5.33% |
| DunedinPACE AA | 48 | Current | -2.04 | 1.617 | 0.215 | 2.17% | 37.92% | 48 | -0.59 | 0.382 | 0.133 | 5.83% | 25.00% |
| DunedinPACE AA | 421 | Nondiabetic | -1.33 | 0.543 | 0.015 | 1.71% | 8.76% | 418 | -0.40 | 0.137 | 0.003 | 2.62% | 10.91% |
| DunedinPACE AA | 42 | Diabetic | 2.39 | 2.609 | 0.368 | 2.20% | 15.71% | 39 | 0.15 | 0.525 | 0.779 | 0.48% | 4.50% |
| DunedinPACE AA | 352 | WMHSC_0_1 | -0.81 | 0.655 | 0.217 | 0.45% | 11.54% | 347 | -0.42 | 0.165 | 0.011 | 2.90% | 8.66% |
| DunedinPACE AA | 97 | WMHSC_2_3 | -1.59 | 1.105 | 0.153 | 2.61% | 24.84% | 96 | -0.21 | 0.230 | 0.360 | 1.65% | 19.56% |
| | **VRM Change** | | | | | | | **Speed Change** | | | | | |
| | **N** | **Subgroup** | **Beta** | **SE** | ***p value*** | ***part R2*** | ***R 2 Model*** | **N** | **Beta** | **SE** | ***p value*** | ***part R2*** | ***R 2 Model*** |

| | | | | | | | | | | | | | |
|---|---|---|---|---|---|---|---|---|---|---|---|---|---|
| BLUP AA | 275 | Young | -0.02 | 0.009 | 0.022 | 1.51% | 21.06% | 276 | -0.09 | 0.047 | 0.045 | 1.17% | 5.00% |
| BLUP AA | 182 | Old | -0.02 | 0.010 | 0.062 | 1.71% | 25.07% | 183 | -0.09 | 0.042 | 0.031 | 3.75% | 14.19% |
| BLUP AA | 209 | Male | -0.02 | 0.009 | 0.067 | 1.56% | 24.98% | 206 | -0.08 | 0.043 | 0.072 | 2.17% | 8.34% |
| BLUP AA | 248 | Female | -0.02 | 0.010 | 0.016 | 1.85% | 19.96% | 253 | -0.08 | 0.051 | 0.119 | 1.09% | 3.98% |
| BLUP AA | 296 | Low_Ed | -0.02 | 0.007 | 0.003 | 3.06% | 10.77% | 300 | -0.05 | 0.040 | 0.236 | 0.54% | 4.68% |
| BLUP AA | 161 | High_Ed | -0.02 | 0.013 | 0.147 | 1.01% | 12.55% | 159 | -0.18 | 0.057 | 0.002 | 5.37% | 18.75% |
| BLUP AA | 162 | Normotensive | -0.02 | 0.011 | 0.033 | 2.12% | 22.94% | 162 | -0.08 | 0.053 | 0.132 | 1.38% | 9.45% |
| BLUP AA | 295 | Hypertensive | -0.02 | 0.008 | 0.044 | 1.20% | 21.61% | 297 | -0.10 | 0.042 | 0.013 | 1.98% | 6.91% |
| BLUP AA | 160 | Normal weight | -0.03 | 0.011 | 0.023 | 2.37% | 28.54% | 162 | -0.15 | 0.058 | 0.012 | 5.26% | 14.37% |
| BLUP AA | 297 | Overweight | -0.02 | 0.008 | 0.056 | 1.26% | 19.44% | 297 | -0.04 | 0.040 | 0.323 | 0.32% | 2.31% |
| BLUP AA | 298 | No_CVD | -0.01 | 0.008 | 0.120 | 0.84% | 20.81% | 297 | -0.08 | 0.040 | 0.051 | 1.55% | 3.95% |
| BLUP AA | 159 | CVD | -0.03 | 0.011 | 0.005 | 3.77% | 23.42% | 162 | -0.11 | 0.057 | 0.063 | 1.84% | 9.77% |
| BLUP AA | 277 | Never | -0.02 | 0.008 | 0.007 | 1.90% | 23.70% | 280 | -0.08 | 0.041 | 0.059 | 1.27% | 3.68% |
| BLUP AA | 132 | Former | -0.02 | 0.013 | 0.100 | 1.65% | 17.14% | 131 | -0.16 | 0.067 | 0.017 | 4.22% | 11.98% |
| BLUP AA | 48 | Current | -0.03 | 0.024 | 0.228 | 2.37% | 41.32% | 48 | -0.03 | 0.102 | 0.784 | 0.18% | 27.13% |
| BLUP AA | 418 | Nondiabetic | -0.02 | 0.007 | 0.001 | 2.08% | 23.18% | 419 | -0.09 | 0.036 | 0.009 | 1.50% | 4.34% |
| BLUP AA | 39 | Diabetic | -0.01 | 0.019 | 0.691 | 0.40% | 14.49% | 40 | -0.12 | 0.087 | 0.188 | 5.47% | 35.80% |
| BLUP AA | 347 | WMHSC_0_1 | -0.02 | 0.008 | 0.028 | 1.26% | 21.23% | 351 | -0.06 | 0.038 | 0.100 | 0.71% | 3.74% |
| BLUP AA | 96 | WMHSC_2_3 | -0.02 | 0.013 | 0.082 | 2.66% | 34.14% | 94 | -0.15 | 0.058 | 0.013 | 6.55% | 19.31% |
| PCPheno AA | 275 | Young | -0.02 | 0.007 | 0.003 | 2.52% | 22.12% | 276 | -0.07 | 0.038 | 0.074 | 0.88% | 4.69% |
| PCPheno AA | 182 | Old | -0.01 | 0.006 | 0.078 | 1.48% | 24.90% | 183 | -0.06 | 0.026 | 0.015 | 4.03% | 14.85% |
| PCPheno AA | 209 | Male | -0.01 | 0.006 | 0.132 | 1.14% | 24.56% | 206 | -0.05 | 0.031 | 0.087 | 1.37% | 8.19% |
| PCPheno AA | 248 | Female | -0.02 | 0.007 | 0.003 | 3.39% | 20.96% | 253 | -0.07 | 0.037 | 0.046 | 1.80% | 4.59% |
| PCPheno AA | 296 | Low_Ed | -0.02 | 0.006 | 0.002 | 3.71% | 11.13% | 300 | -0.03 | 0.031 | 0.261 | 0.35% | 4.64% |
| PCPheno AA | 161 | High_Ed | -0.01 | 0.009 | 0.129 | 1.35% | 12.67% | 159 | -0.11 | 0.038 | 0.003 | 4.87% | 18.10% |
| PCPheno AA | 162 | Normotensive | -0.03 | 0.009 | 0.000 | 6.40% | 27.37% | 162 | -0.06 | 0.042 | 0.187 | 0.94% | 9.13% |
| PCPheno AA | 295 | Hypertensive | -0.01 | 0.006 | 0.135 | 0.75% | 21.10% | 297 | -0.08 | 0.029 | 0.009 | 1.92% | 7.09% |

| | | | | | | | | | | | | | |
|---|---|---|---|---|---|---|---|---|---|---|---|---|---|
| PCPheno AA | 160 | Normal weight | -0.02 | 0.010 | 0.055 | 1.78% | 27.81% | 162 | -0.11 | 0.052 | 0.029 | 3.38% | 13.45% |
| PCPheno AA | 297 | Overweight | -0.01 | 0.005 | 0.007 | 2.30% | 20.43% | 297 | -0.05 | 0.026 | 0.079 | 0.84% | 3.03% |
| PCPheno AA | 298 | No_CVD | -0.02 | 0.006 | 0.008 | 2.21% | 22.06% | 297 | -0.05 | 0.029 | 0.065 | 1.39% | 3.81% |
| PCPheno AA | 159 | CVD | -0.02 | 0.008 | 0.055 | 1.75% | 21.24% | 162 | -0.08 | 0.042 | 0.051 | 1.76% | 9.98% |
| PCPheno AA | 277 | Never | -0.02 | 0.006 | 0.000 | 3.48% | 25.27% | 280 | -0.05 | 0.031 | 0.131 | 0.86% | 3.22% |
| PCPheno AA | 132 | Former | 0.00 | 0.008 | 0.593 | 0.26% | 15.45% | 131 | -0.11 | 0.042 | 0.008 | 4.33% | 12.97% |
| PCPheno AA | 48 | Current | -0.02 | 0.018 | 0.325 | 2.42% | 40.54% | 48 | 0.06 | 0.076 | 0.464 | 0.85% | 28.05% |
| PCPheno AA | 418 | Nondiabetic | -0.02 | 0.005 | 0.001 | 2.15% | 23.32% | 419 | -0.07 | 0.025 | 0.008 | 1.44% | 4.41% |
| PCPheno AA | 39 | Diabetic | -0.02 | 0.016 | 0.280 | 1.79% | 17.68% | 40 | -0.07 | 0.069 | 0.298 | 2.44% | 34.27% |
| PCPheno AA | 347 | WMHSC_0_1 | -0.02 | 0.006 | 0.002 | 2.31% | 22.29% | 351 | -0.06 | 0.031 | 0.041 | 0.95% | 4.15% |
| PCPheno AA | 96 | WMHSC_2_3 | -0.01 | 0.007 | 0.254 | 1.35% | 32.76% | 94 | -0.07 | 0.030 | 0.022 | 5.85% | 18.41% |
| PCGrim AA | 275 | Young | -0.02 | 0.014 | 0.189 | 0.37% | 19.98% | 276 | -0.17 | 0.071 | 0.020 | 1.67% | 5.49% |
| PCGrim AA | 182 | Old | -0.03 | 0.014 | 0.035 | 2.26% | 25.49% | 183 | -0.13 | 0.063 | 0.038 | 4.64% | 14.02% |
| PCGrim AA | 209 | Male | -0.02 | 0.013 | 0.221 | 0.71% | 24.27% | 206 | -0.17 | 0.067 | 0.014 | 2.70% | 9.64% |
| PCGrim AA | 248 | Female | -0.03 | 0.015 | 0.057 | 1.12% | 19.24% | 253 | -0.17 | 0.073 | 0.026 | 2.65% | 4.99% |
| PCGrim AA | 296 | Low_Ed | -0.01 | 0.012 | 0.203 | 0.53% | 8.54% | 300 | -0.12 | 0.062 | 0.056 | 1.02% | 5.43% |
| PCGrim AA | 161 | High_Ed | -0.03 | 0.018 | 0.056 | 1.77% | 13.46% | 159 | -0.19 | 0.081 | 0.023 | 4.95% | 16.03% |
| PCGrim AA | 162 | Normotensive | -0.03 | 0.018 | 0.139 | 0.97% | 21.71% | 162 | -0.14 | 0.081 | 0.087 | 1.53% | 9.85% |
| PCGrim AA | 295 | Hypertensive | -0.02 | 0.012 | 0.143 | 0.76% | 21.08% | 297 | -0.17 | 0.062 | 0.006 | 2.64% | 7.38% |
| PCGrim AA | 160 | Normal weight | -0.03 | 0.020 | 0.086 | 1.73% | 27.46% | 162 | -0.20 | 0.106 | 0.065 | 4.11% | 12.65% |
| PCGrim AA | 297 | Overweight | -0.02 | 0.011 | 0.156 | 0.60% | 18.97% | 297 | -0.13 | 0.054 | 0.015 | 1.63% | 3.99% |
| PCGrim AA | 298 | No_CVD | -0.02 | 0.012 | 0.058 | 1.00% | 21.14% | 297 | -0.12 | 0.060 | 0.042 | 1.78% | 4.05% |
| PCGrim AA | 159 | CVD | -0.02 | 0.018 | 0.267 | 0.57% | 19.91% | 162 | -0.25 | 0.089 | 0.005 | 3.82% | 12.41% |
| PCGrim AA | 277 | Never | -0.03 | 0.014 | 0.021 | 1.15% | 23.14% | 280 | -0.18 | 0.069 | 0.012 | 1.97% | 4.67% |
| PCGrim AA | 132 | Former | -0.01 | 0.018 | 0.466 | 0.52% | 15.63% | 131 | -0.20 | 0.091 | 0.027 | 2.97% | 11.36% |
| PCGrim AA | 48 | Current | -0.01 | 0.027 | 0.796 | 0.13% | 39.05% | 48 | 0.00 | 0.113 | 0.988 | 0.00% | 26.98% |
| PCGrim AA | 418 | Nondiabetic | -0.02 | 0.011 | 0.039 | 0.70% | 21.96% | 419 | -0.15 | 0.053 | 0.005 | 1.79% | 4.58% |

| | | | | | | | | | | | | | |
|---|---|---|---|---|---|---|---|---|---|---|---|---|---|
| PCGrim AA | 39 | Diabetic | -0.03 | 0.029 | 0.306 | 2.16% | 17.32% | 40 | -0.22 | 0.137 | 0.118 | 3.96% | 37.45% |
| PCGrim AA | 347 | WMHSC_0_1 | -0.02 | 0.012 | 0.108 | 0.59% | 20.70% | 351 | -0.19 | 0.060 | 0.002 | 2.53% | 5.62% |
| PCGrim AA | 96 | WMHSC_2_3 | -0.03 | 0.018 | 0.088 | 2.95% | 34.06% | 94 | -0.14 | 0.076 | 0.068 | 4.01% | 16.46% |
| DunedinPACE AA | 275 | Young | -0.42 | 0.361 | 0.249 | 0.37% | 19.85% | 276 | -2.08 | 1.879 | 0.269 | 0.34% | 3.98% |
| DunedinPACE AA | 182 | Old | 0.15 | 0.337 | 0.649 | 0.08% | 23.59% | 183 | -0.47 | 1.479 | 0.751 | 0.06% | 11.86% |
| DunedinPACE AA | 209 | Male | 0.28 | 0.343 | 0.422 | 0.19% | 23.94% | 206 | -0.44 | 1.742 | 0.801 | 0.03% | 6.82% |
| DunedinPACE AA | 248 | Female | -0.68 | 0.359 | 0.059 | 1.19% | 19.21% | 253 | -1.97 | 1.797 | 0.273 | 0.55% | 3.48% |
| DunedinPACE AA | 162 | Normotensive | -0.08 | 0.450 | 0.863 | 0.02% | 20.57% | 162 | -1.57 | 2.079 | 0.451 | 0.40% | 8.41% |
| DunedinPACE AA | 295 | Hypertensive | -0.18 | 0.303 | 0.552 | 0.15% | 20.58% | 297 | -0.96 | 1.563 | 0.539 | 0.13% | 4.99% |
| DunedinPACE AA | 160 | Normal weight | 0.33 | 0.470 | 0.490 | 0.18% | 26.22% | 162 | -1.73 | 2.480 | 0.487 | 0.32% | 10.91% |
| DunedinPACE AA | 297 | Overweight | -0.27 | 0.299 | 0.358 | 0.32% | 18.63% | 297 | -1.77 | 1.437 | 0.220 | 0.42% | 2.49% |
| DunedinPACE AA | 296 | Low_Ed | -0.06 | 0.300 | 0.848 | 0.01% | 8.02% | 300 | 0.47 | 1.569 | 0.766 | 0.04% | 4.24% |
| DunedinPACE AA | 161 | High_Ed | -0.42 | 0.465 | 0.372 | 0.55% | 11.77% | 159 | -1.91 | 2.123 | 0.370 | 0.79% | 13.49% |
| DunedinPACE AA | 298 | No_CVD | -0.34 | 0.327 | 0.302 | 0.38% | 20.44% | 297 | -1.90 | 1.628 | 0.243 | 0.56% | 3.11% |
| DunedinPACE AA | 159 | CVD | 0.12 | 0.384 | 0.754 | 0.08% | 19.29% | 162 | 0.48 | 1.974 | 0.808 | 0.05% | 7.69% |
| DunedinPACE AA | 277 | Never | -0.21 | 0.333 | 0.525 | 0.07% | 21.71% | 280 | -1.82 | 1.667 | 0.275 | 0.35% | 2.83% |
| DunedinPACE AA | 132 | Former | -0.06 | 0.455 | 0.889 | 0.02% | 15.27% | 131 | -1.50 | 2.400 | 0.533 | 0.31% | 7.98% |
| DunedinPACE AA | 48 | Current | -0.58 | 0.777 | 0.457 | 1.08% | 39.86% | 48 | -1.13 | 3.273 | 0.733 | 0.31% | 27.22% |
| DunedinPACE AA | 418 | Nondiabetic | -0.13 | 0.262 | 0.617 | 0.05% | 21.18% | 419 | -1.31 | 1.312 | 0.319 | 0.19% | 2.96% |
| DunedinPACE AA | 39 | Diabetic | -0.45 | 0.766 | 0.560 | 1.03% | 15.08% | 40 | 0.20 | 3.875 | 0.960 | 0.01% | 31.64% |
| DunedinPACE AA | 347 | WMHSC_0_1 | -0.24 | 0.309 | 0.434 | 0.21% | 20.23% | 351 | -2.66 | 1.564 | 0.090 | 0.72% | 3.78% |
| DunedinPACE AA | 96 | WMHSC_2_3 | -0.24 | 0.402 | 0.553 | 0.33% | 31.98% | 94 | 0.38 | 1.773 | 0.832 | 0.05% | 12.99% |

### *Supplementary Table 10: ApoE4 specific subgroup associations*

| | ApoE4 non-carriers | | | | | | | ApoE4 carriers | | | | | |
|---|---|---|---|---|---|---|---|---|---|---|---|---|---|
| | **PREDICTOR** | **N** | **Beta** | **SE** | ***p value*** | ***part R2*** | ***R 2 Model*** | **N** | **Beta** | **SE** | ***p value*** | ***part R2*** | ***R 2 Model*** |
| **WCST** | BLUP AA | 580 | -0.28 | 0.084 | 0.001 | 1.87% | 3.94% | 134 | -0.38 | 0.151 | 0.012 | 5.28% | 18.20% |
| | PCPheno AA | 580 | -0.25 | 0.065 | 0.000 | 2.57% | 4.56% | 134 | -0.26 | 0.124 | 0.041 | 4.75% | 16.74% |
| | PCGrim AA | 580 | -0.22 | 0.129 | 0.088 | 0.49% | 2.53% | 134 | 0.03 | 0.250 | 0.913 | 0.02% | 13.84% |
| | DunedinPACE AA | 580 | -6.70 | 3.079 | 0.030 | 0.87% | 2.83% | 134 | -5.05 | 6.059 | 0.406 | 0.43% | 14.32% |
| **VSM** | BLUP AA | 563 | -0.45 | 0.361 | 0.209 | 0.27% | 24.20% | 131 | -1.60 | 0.826 | 0.056 | 2.32% | 30.72% |
| | PCPheno AA | 563 | -0.32 | 0.267 | 0.228 | 0.30% | 24.19% | 131 | -1.14 | 0.677 | 0.095 | 2.53% | 30.20% |
| | PCGrim AA | 563 | -0.72 | 0.553 | 0.195 | 0.66% | 24.22% | 131 | -0.07 | 1.374 | 0.961 | 0.00% | 28.54% |
| | DunedinPACE AA | 563 | -33.18 | 13.353 | 0.013 | 1.63% | 24.83% | 131 | -40.26 | 33.584 | 0.233 | 1.00% | 29.39% |
| **TMT change** | BLUP AA | 372 | -0.03 | 0.016 | 0.085 | 0.81% | 12.73% | 91 | -0.04 | 0.032 | 0.171 | 1.73% | 25.26% |
| | PCPheno AA | 372 | -0.03 | 0.012 | 0.030 | 1.41% | 13.15% | 91 | -0.03 | 0.025 | 0.306 | 1.64% | 24.47% |
| | PCGrim AA | 372 | -0.05 | 0.024 | 0.026 | 1.54% | 13.22% | 91 | -0.08 | 0.057 | 0.167 | 1.71% | 25.29% |
| | DunedinPACE AA | 372 | -0.84 | 0.602 | 0.163 | 0.71% | 12.49% | 91 | -2.43 | 1.345 | 0.075 | 3.96% | 26.49% |
| **VSM change** | BLUP AA | 369 | -0.01 | 0.004 | 0.004 | 2.14% | 8.90% | 88 | -0.03 | 0.010 | 0.005 | 9.97% | 23.63% |
| | PCPheno AA | 369 | -0.01 | 0.003 | 0.027 | 1.29% | 7.97% | 88 | -0.02 | 0.007 | 0.021 | 11.46% | 21.04% |
| | PCGrim AA | 369 | -0.01 | 0.005 | 0.012 | 1.84% | 8.35% | 88 | 0.00 | 0.017 | 0.781 | 0.09% | 15.39% |
| | DunedinPACE AA | 369 | -0.39 | 0.140 | 0.005 | 2.56% | 8.71% | 88 | -0.05 | 0.417 | 0.904 | 0.03% | 15.31% |
| **VRM change** | BLUP AA | 369 | -0.01 | 0.007 | 0.094 | 0.74% | 19.59% | 88 | -0.05 | 0.015 | 0.001 | 11.10% | 39.76% |
| | PCPheno AA | 369 | -0.01 | 0.005 | 0.022 | 1.28% | 20.14% | 88 | -0.03 | 0.012 | 0.011 | 10.29% | 36.58% |
| | PCGrim AA | 369 | -0.02 | 0.011 | 0.099 | 0.69% | 19.57% | 88 | -0.04 | 0.027 | 0.148 | 1.42% | 32.73% |
| | DunedinPACE AA | 369 | -0.19 | 0.272 | 0.483 | 0.13% | 19.07% | 88 | 0.12 | 0.665 | 0.857 | 0.04% | 30.87% |
| **Speed change** | BLUP AA | 369 | -0.06 | 0.035 | 0.118 | 0.75% | 4.40% | 90 | -0.23 | 0.089 | 0.010 | 7.72% | 16.48% |
| | PCPheno AA | 369 | -0.069 | 0.025 | 0.006 | 2.00% | 5.76% | 90 | -0.05 | 0.072 | 0.490 | 0.48% | 9.62% |
| | PCGrim AA | 369 | -0.163 | 0.051 | 0.001 | 3.22% | 6.46% | 90 | -0.139 | 0.163 | 0.398 | 0.44% | 9.89% |
| | DunedinPACE AA | 369 | -2.33 | 1.307 | 0.076 | 0.88% | 4.59% | 90 | 4.03 | 3.865 | 0.301 | 2.41% | 10.31% |

*Supplementary Table 11: Variability explained by age acceleration measures compared to other risk factors in cognition*

| WCST_BL | | | | |
|---|---|---|---|---|
| Pred | BLUP | **PCPheno** | PCGrim | DunedinPACE |
| **AA** | **2.3%** | **3.0%** | **0.4%** | **1.0%** |
| Education | 0.87% | 0.70% | 0.85% | 0.80% |
| age_BL | 0.55% | 0.52% | 0.54% | 0.52% |
| CVD | 0.55% | 0.47% | 0.51% | 0.44% |
| Diabetes | 0.21% | 0.28% | 0.16% | 0.20% |
| Cholesterol | 0.14% | 0.04% | 0.13% | 0.06% |
| Smoking | 0.03% | 0.03% | 0.09% | 0.07% |
| BMI | 0.01% | 0.03% | 0.03% | 0.09% |
| sex | 0.05% | 0.01% | 0.09% | 0.07% |
| apoe4_bi | 0.00% | 0.00% | 0.01% | 0.01% |
| Hypertension | 0.02% | 0.00% | 0.00% | 0.00% |

| TMT Change | | | | |
|---|---|---|---|---|
| Pred | BLUP | PCPheno | PCGrim | **DunedinPACE** |
| Diabetes | 5.94% | 5.91% | 5.85% | 5.93% |
| Education | 4.25% | 4.20% | 4.24% | 4.17% |
| **AA** | **1.0%** | **1.56%** | **1.34%** | **0.97%** |
| CVD | 1.46% | 1.42% | 1.43% | 1.39% |
| apoe4_bi | 0.15% | 0.16% | 0.09% | 0.09% |
| Smoking | 0.11% | 0.13% | 0.32% | 0.14% |
| sex | 0.12% | 0.07% | 0.04% | 0.10% |
| BMI | 0.05% | 0.06% | 0.13% | 0.17% |
| Hypertension | 0.05% | 0.02% | 0.02% | 0.02% |
| Cholesterol | 0.04% | 0.00% | 0.00% | 0.00% |
| age_BL | 0.01% | 0.00% | 0.00% | 0.00% |

| VRM Change | | | | |
|---|---|---|---|---|
| Pred | BLUP | PCPheno | PCGrim | **DunedinPACE** |

| Visual Memory | | | | |
|---|---|---|---|---|
| Pred | BLUP | PCPheno | PCGrim | **DunedinPACE** |
| age_BL | 14.84% | 14.85% | 14.84% | 14.87% |
| Education | 7.09% | 7.07% | 7.09% | 7.01% |
| **AA** | **0.5%** | **0.6%** | **0.5%** | **1.6%** |
| CVD | 0.83% | 0.83% | 0.81% | 0.74% |
| Smoking | 0.90% | 0.88% | 0.71% | 0.57% |
| Hypertension | 0.21% | 0.16% | 0.18% | 0.14% |
| Diabetes | 0.07% | 0.06% | 0.07% | 0.03% |
| Cholesterol | 0.01% | 0.00% | 0.01% | 0.00% |
| BMI | 0.03% | 0.03% | 0.02% | 0.00% |
| apoe4_bi | 0.00% | 0.00% | 0.00% | 0.00% |
| sex | 0.01% | 0.00% | 0.01% | 0.00% |

| VSM Change | | | | |
|---|---|---|---|---|
| Pred | BLUP | PCPheno | PCGrim | **DunedinPACE** |
| Education | 5.96% | 5.79% | 5.83% | 5.74% |
| **AA** | **3.20%** | **2.31%** | **1.48%** | **2.08%** |
| CVD | 0.75% | 0.74% | 0.76% | 0.64% |
| BMI | 0.36% | 0.30% | 0.17% | 0.09% |
| Smoking | 0.14% | 0.20% | 0.18% | 0.12% |
| Cholesterol | 0.13% | 0.04% | 0.05% | 0.01% |
| age_BL | 0.12% | 0.06% | 0.07% | 0.05% |
| sex | 0.05% | 0.07% | 0.10% | 0.05% |
| apoe4_bi | 0.03% | 0.04% | 0.10% | 0.11% |
| Hypertension | 0.00% | 0.09% | 0.07% | 0.08% |
| Diabetes | 0.00% | 0.01% | 0.00% | 0.01% |

| Speed Change | | | | |
|---|---|---|---|---|
| Pred | BLUP | PCPheno | PCGrim | **DunedinPACE** |

| | | | | | | | | | | |
|---|---|---|---|---|---|---|---|---|---|---|
| Education | 17.90% | 17.85% | 17.86% | 17.85% | | **AA** | **1.64%** | **1.39%** | **2.13%** | **0.17%** |
| **AA** | **1.61%** | **2.02%** | **0.8%** | **0.09%** | | Cholesterol | 0.33% | 0.50% | 0.51% | 0.41% |
| CVD | 0.65% | 0.63% | 0.67% | 0.69% | | sex | 0.21% | 0.26% | 0.48% | 0.15% |
| Smoking | 0.41% | 0.50% | 0.51% | 0.38% | | Education | 0.35% | 0.42% | 0.42% | 0.39% |
| Hypertension | 0.23% | 0.40% | 0.38% | 0.35% | | BMI | 0.14% | 0.16% | 0.31% | 0.19% |
| Diabetes | 0.02% | 0.03% | 0.02% | 0.06% | | Smoking | 0.65% | 0.68% | 0.18% | 0.68% |
| apoe4_bi | 0.03% | 0.03% | 0.07% | 0.06% | | Hypertension | 0.26% | 0.16% | 0.17% | 0.18% |
| age_BL | 0.08% | 0.02% | 0.03% | 0.04% | | CVD | 0.19% | 0.18% | 0.17% | 0.21% |
| BMI | 0.01% | 0.02% | 0.06% | 0.04% | | apoe4_bi | 0.02% | 0.02% | 0.06% | 0.05% |
| Cholesterol | 0.06% | 0.00% | 0.01% | 0.02% | | Diabetes | 0.04% | 0.07% | 0.03% | 0.11% |
| sex | 0.02% | 0.00% | 0.00% | 0.06% | | age_BL | 0.00% | 0.01% | 0.01% | 0.01% |
